# Development and Validation of an Interpretable 3-day Intensive Care Unit Readmission Prediction Model Using Explainable Boosting Machines

**DOI:** 10.1101/2021.11.01.21265700

**Authors:** Stefan Hegselmann, Christian Ertmer, Thomas Volkert, Antje Gottschalk, Martin Dugas, Julian Varghese

## Abstract

Intensive care unit readmissions are associated with mortality and bad outcomes. Machine learning could help to identify patients at risk to improve discharge decisions. However, many models are black boxes, so that dangerous properties might remain unnoticed. In this study, an inherently interpretable model for 3-day ICU readmission prediction was developed. We used a retrospective cohort of 15,589 ICU stays and 169 variables collected between 2006 and 2019. A team of doctors inspected the model, checked the plausibility of each component, and removed problematic parts. Qualitative feedback revealed several challenges for interpretable machine learning in healthcare. The resulting model used 67 features and showed an area under the precision-recall curve of 0.119±0.020 and an area under the receiver operating characteristic curve of 0.680±0.025. This is on par with state-of-the-art gradient boosting machines and outperforms the Simplified Acute Physiology Score II. External validation with the Medical Information Mart for Intensive Care database version IV confirmed our findings. Hence, a machine learning model for readmission prediction with a high level of human control is feasible without sacrificing performance.

---

Intensive care units (ICU) treat patients with severe or life-threatening conditions by providing specialized care, modalities for life support, and extensive monitoring capabilities^1^. Large quantities of data are collected continuously for each patient in a Patient Data Management System (PDMS)^2^. Decisions to end intensive treatment and transfer a patient to a lower level of care are complex and include consideration of several aspects^3^. Discharging a patient too early can lead to deterioration of the patient’s health status with subsequent readmission, which is associated with mortality and bad outcomes such as an increased length of stay^4–6^. A study from 2013 including 105 ICUs in the United States found a median ICU readmission rate of 5.9%^7^. Several risk factors have been identified for ICU readmissions such as admission origin, comorbidities, physiological abnormalities, and age^6^. However, it can be challenging for clinicians to incorporate all available information appropriately and interpret them in an individual patient case^8^.

Machine learning (ML) can automatically detect relevant patterns in large quantities of data and has already shown its potential to transform healthcare^9^. However, many ML models are black boxes because they are complex and humans are unable to understand them^10^. Studies revealed that ML for healthcare contained an unnoticed racial bias^11^ or relies on spurious correlations^12^. This is especially dangerous in a sensitive domain such as healthcare^13^. Interpretable ML can be one approach to alleviate these issues by adding the ability to explain in terms understandable to humans. This allows to put the responsibility to ensure additional desiderata such as robustness, causality, and fairness on human practitioners^14^. Many studies use so-called post hoc explanations to interpret black box models^15–17^. However, popular explanation methods such as local interpretable model-agnostic explanations (LIME)^18^ or Shapley additive explanations (SHAP)^19^ showed shortcomings regarding robustness and adversarial attacks^20–22^. Hence in this work, we consider inherently interpretable or transparent models^10,23^, which are no black boxes and allow humans to understand the model.

There is a research gap of studies evaluating transparent ML models for healthcare including human evaluation. A recent review on explainable artificial intelligence using electronic health records showed that nine out of 42 papers used inherently interpretable models^24^. Applications included mortality prediction, disease classification, risk stratification, and biomedical knowledge discovery. However, only three studies reported human expert confirmation of their results, which is considered essential for meaningful evaluation of interpretable ML^14^. Another review including 42 studies for hospital readmission prediction found that none applied interpretable ML^25^. For ICU readmission prediction, we identified two papers^26,27^ that explicitly developed interpretable models based on rule sets and logistic regression. However, no human validation of the results was performed.

In this work, we developed an inherently interpretable ML model for 3-day ICU readmission prediction trained on routinely collected data. The main goal is twofold. First, we suggest a procedure to develop a transparent model that includes clinicians to inspect and verify the entire model. This process is evaluated to determine its consequences and reveal possible problems. We used Explainable Boosting Machines (EBM) for our experiments which learn modular risk functions that can be inspected and removed by humans^28^. They showed good performance across several tasks^29,30^ and already proved useful for the healthcare domain^12,31^. Second, the resulting EBM model will be compared to different baseline and state-of-the-art ML models to assess the effect of transparency on performance. This comparison included the validated Simplified Acute Physiology Score (SAPS) II, logistic regression (LR) with feature selection, gradient boosting machines (GBMs), and recurrent neural networks (RNNs) with long short-term memory units. We extracted a local cohort to conduct our experiment based on data collected for ICU stays between 2006 and 2019 managed by the Department of Anesthesiology, Intensive Care and Pain Medicine (ANIT-UKM) at the University Hospital Münster (UKM), Germany. For external validation, the Medical Information Mart for Intensive Care database version IV (MIMIC-IV) containing ICU data collected from 2008 to 2019 was used^32,33^.

## Results

### Cohort selection and label generation

We considered all ICU and intermediate care (IMC) transfers of adult patients between 2006 and 2019 managed by the Department of Anesthesiology and Intensive Care Medicine at the University Hospital Münster (ANIT-UKM) for this study. Initially, 199,764 relevant transfers were obtained from the hospital information system (HIS) of the UKM. After data cleaning and cohort selection (Figure 4), 15,589 ICU stays of 14,188 patients were included. Most transfers were removed because another department was responsible or it was an IMC transfer. Stays of patients that were discharged to a general ward and were readmitted to any ICU (n=822) or IMC unit (n=31) or died within three days in the UKM (n=38) were labeled positively (n=891, 5.7%). Table 1 summarizes key characteristics of the resulting UKM cohort. Patients of included ICU stays were on average 63.33±14.73 years old and more than two-thirds had reported gender male (n=10,670). ICU stays with 3-day readmission or death after discharge showed several differences: patients were on average around three years older, the ratio of male patients further increased from 68.3% to 70.8%, and the mean length of the previous ICU stay was about 13.5 hours longer.

**Table 1.** Overview of UKM cohort. Key characteristics of all included ICU stays and divided by their labels. Note that this information is based on ICU stays so that a single patient can be considered more than once. A description and additional information of the ICUs are provided in the supplement.

| Characteristic | All ICU stays | No 3-day readmission or death after ICU discharge | 3-day readmissions or death after ICU discharge |
| --- | --- | --- | --- |
| Number of stays | 15,589 (100.0%) | 14,698 (94.3%) | 891 (5.7%) |
| Number of patients | 14,188 (100.0%) | 13,349 (94.1%) | 839 (5.9%) |
| Mean age (SD) | 63.33 (14.73) | 63.16 (14.77) | 66.08 (13.85) |
| Gender female | 4,919 (100.0%) | 4,659 (94.7%) | 260 (5.3%) |
| Gender male | 10,670 (100.0%) | 10,039 (94.1%) | 631 (5.9%) |
| Mean length of ICU stay (SD) | 3.70 days (8.08 days) | 3.67 days (8.11 days) | 4.23 days (7.53 days) |
| ICU at discharge | ICU 1 (n=4,063)<br>ICU 2 (n=6,402)<br>ICU 3 (n=1,034)<br>ICU 4 (n=4,090) | ICU 1 (n=3,820)<br>ICU 2 (n=6,035)<br>ICU 3 (n=960)<br>ICU 4 (n=3,883) | ICU 1 (n=243)<br>ICU 2 (n=367)<br>ICU 3 (n=74)<br>ICU 4 (n=207) |

We used data that was routinely collected in the ICU. To this end, 6,496 item definitions with 651,258,647 time-stamped recordings were extracted from the PDMS of the ANIT-UKM. Extensive data cleaning and preprocessing were applied to the raw recordings (see methods). We included 120 non-medication and 49 medication variables with an acceptable data quality that were considered relevant for model development (see supplement). EBMs cannot incorporate time-series data. Hence, we assigned variables to nine different feature classes based on their data and generated descriptive statistics for different time intervals (see supplement). This resulted in 1,423 features for each ICU stay.

### Creation and inspection of interpretable EBM model with limited size

An EBM trained on all features would contain 1,423 1D risk functions hampering interpretability due to its size. Hence, we wanted to create a model with at most 80 1D functions, which was determined a priori as a suitable maximum. To this end, we first performed parameter tuning on the validation split to obtain useful hyperparameters for the model (see supplement). We used the area under the precision-recall curve (PR-AUC) as the primary performance indicator in our experiments due to the label imbalance. We also report the area under the receiver operating characteristic curve (ROC-AUC) since it is commonly used in the medical literature. The best EBM with 1,423 1D risk functions achieved a PR-AUC of 0.151±0.028 and ROC-AUC of 0.652±0.034 on the held-out split. Next, we performed a greedy stepwise forward selection for risk functions. In each round, we selected the function with the largest average importance on the five temporal splits when added to the model. We carried out this procedure for different bin sizes for the discretization of continuous features and evaluated the PR-AUC on the validation split. The best model had a bin size of 200 and contained 80 1D risk functions. It showed a PR-AUC of 0.130±0.021 and a ROC-AUC of 0.681±0.026. We repeated the same procedure for 2D risk functions. Five 2D functions were added with a bin size of four and the model achieved a PR-AUC of 0.113±0.018 and ROC-AUC of 0.646±0.01. All 85 risk functions of the resulting EBM model are shown in the supplement.

We inspected the resulting EBM model in a team of two clinicians to identify problematic risk functions and remove them. A priori to the model inspection we determined four potential problems that we assigned to risk functions during inspection:

1. It encodes healthcare disparities that should not be reproduced (n=0)
2. It contains undesirable artifacts of the data generation process (n=8)
3. It contradicts medical knowledge (n=13)
4. It is not interpretable so that its effect cannot be clearly determined (n=17)

Model inspection took four hours, i.e. approximately three minutes per function. Not all risk functions with a problem were excluded, so we put the risk functions into three classes: included without problems (n=52), included with problems (n=15), and excluded with problems (n=18). Most functions were excluded due to a lack of interpretability (n=10) followed by undesirable artifacts (n=6) and contradiction of medical knowledge (n=6). Note that more than one problem could be assigned to each risk function. Five functions for partial thromboplastin time (PTT) were excluded due to artifacts (problem 2). With the aid of the feature histograms, we recognized a change in the PTT measurement procedure since 2019 invalidating the risk functions learned on the training set. Also, all 2D risk functions were labeled as not interpretable (problem 4) and were excluded from the model. Figure 2 shows two included 1D risk functions and three 1D and one 2D functions that were excluded due to different problems. The EBM model after model inspection contained 67 1D risk functions and achieved a PR-AUC of 0.119±0.020 and a ROC-AUC of 0.680±0.025 on the held-out data. Hence, inspection decreased the PR-AUC and increased the ROC-AUC compared to a model trained on all 1D risk functions.

**Figure 1.**
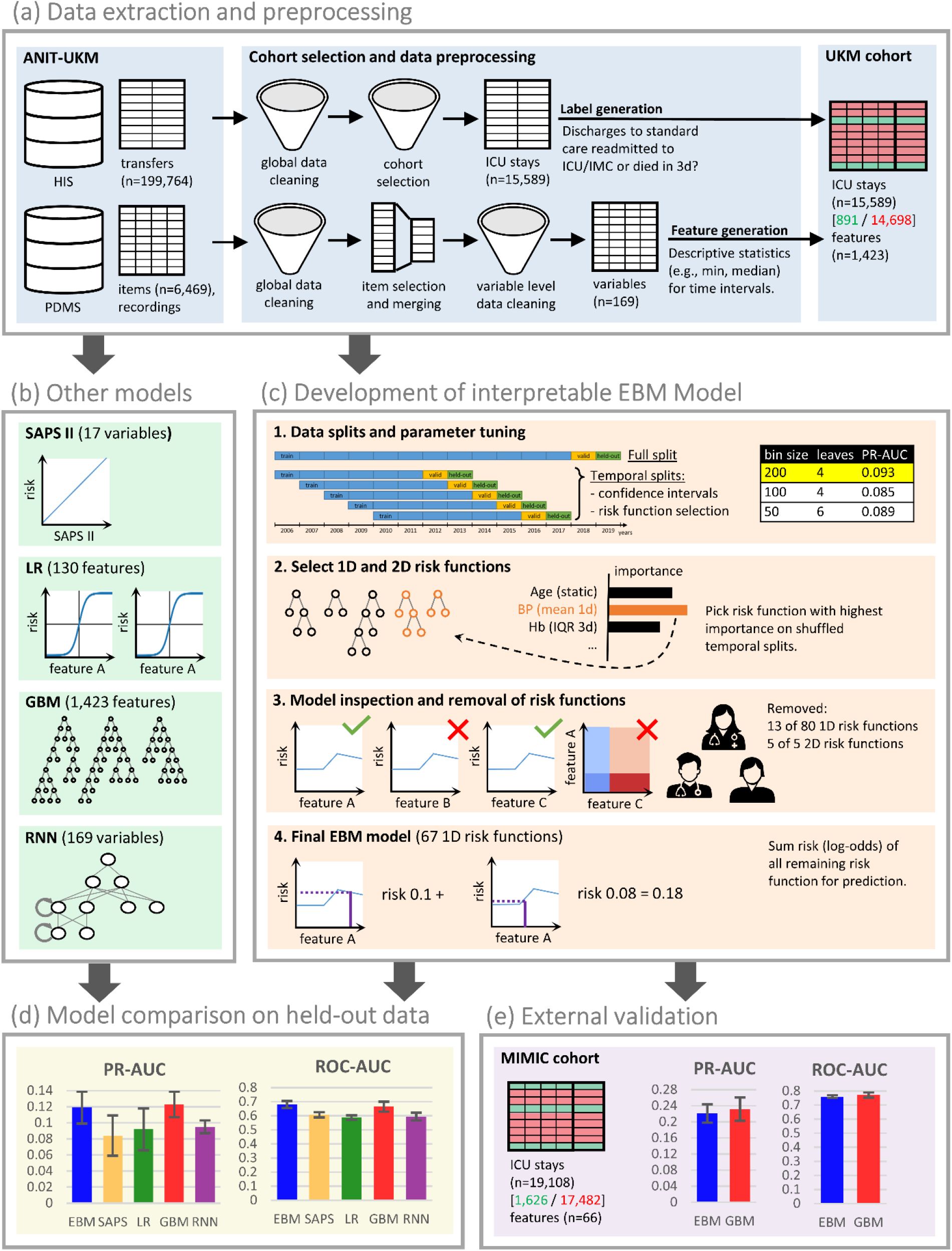
Flowchart of the study. (a) We created a local cohort for the development of ML models. ICU transfers were extracted from the hospital information system (HIS) and ICU data from the PDMS. Extensive preprocessing was applied to clean the data. We generated labels for 3-day ICU readmission and descriptive statistics as features. (b) Four ML models were developed for comparison. For LR we also performed feature selection. The RNN utilizes time series data directly so that no features were necessary. (c) Development of the EBM model comprised of four steps. We performed parameter tuning (also for the other models) and performed greedy risk function selection based on the importance determined on the temporal splits. In step three we inspected the model with a team of clinicians to identify and remove problematic risk functions. The remaining risk functions were used for predictions. (d) We evaluated all models for their area under the precision-recall curve (PR-AUC) and area under the receiver operating characteristic curve (ROC-AUC) on the held-out split (see Figure 5). (e) External validation for the EBM and GBM model was performed on the MIMIC-IV database. (d), (e)) Error bars were determined with the standard deviation on five temporal splits. EBM: explainable boosting machine, SAPS II: Simplified Acute Physiology Score II, LR: logistic regression, GBM: gradient boosting machine, RNN: recurrent neural networks with long short-term memory.

**Figure 2.**
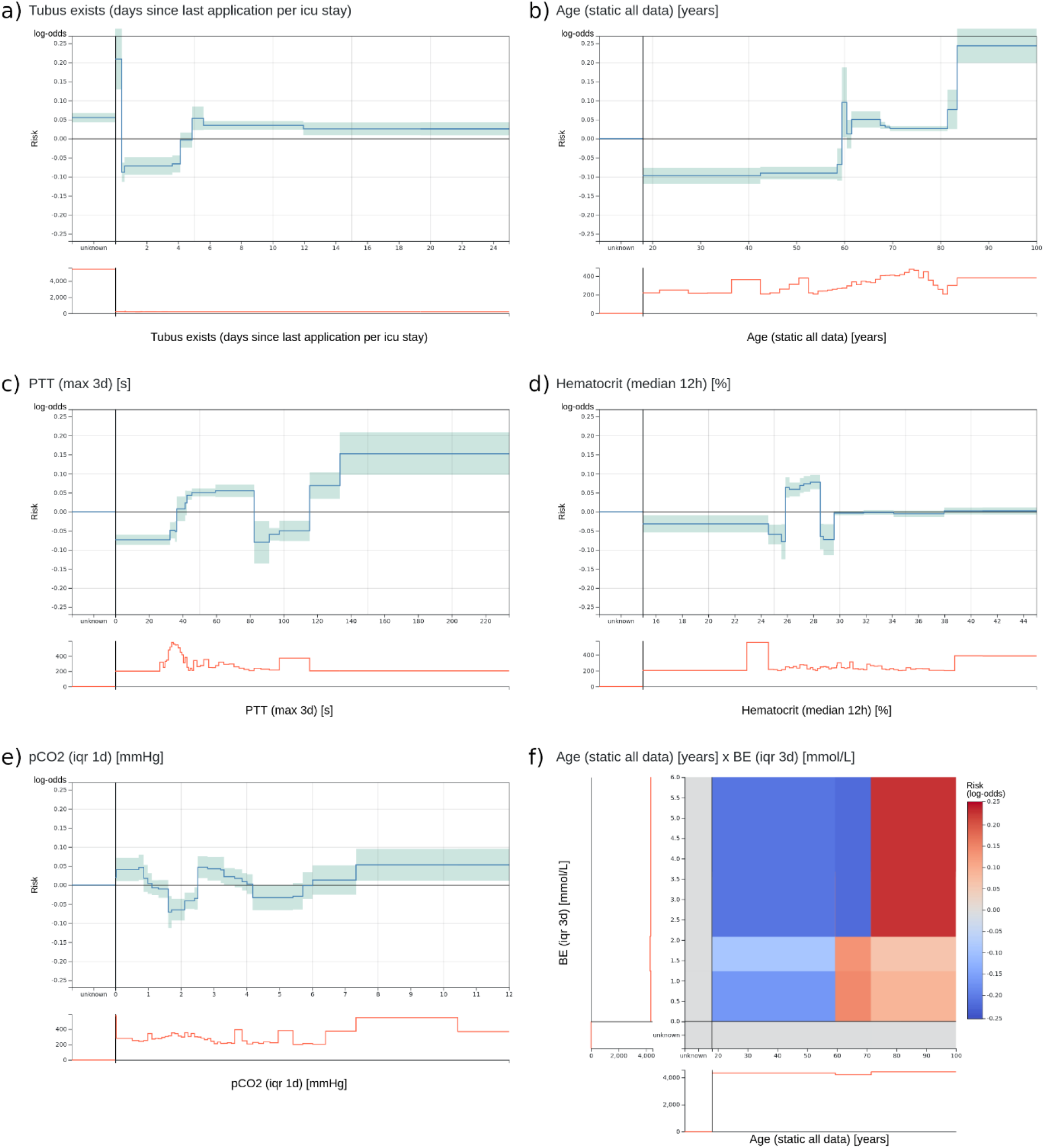
Two most important risk functions of the EBM model and four risk functions excluded during model inspection. (a), (b) Two most important risk functions included in the EBM model. (a) Contains the number of days since the last existence of an orotracheal tube. Patients that have an endotracheal immediately before discharge have a highly increased risk. Lower risk is assigned to values between 0.4 and 4.1 days which probably corresponds to patients that were extubated regularly. Also, patients with no endotracheal tube (unknown) receive an increased risk. (b) The risk function for age shows an increased risk for higher age values. There is a peak at 60 years with no obvious explanation. (c) A maximum PTT value over the last three days before discharge between 82.5 and 115.5 seconds gets a lower risk for 3-day ICU readmission. It was identified that this is an artifact of the previous procedure to determine the PTT for cardiac surgery patients. This will not generalize for future data. (d) For a median hematocrit between 24.875 and 28.525%, the model determined an elevated risk. For slightly lower and higher values the risk is negative. This is against common medical knowledge where a decreasing hematocrit value should be associated with increased risk. (e) The IQR of the pCO2 over the last day before discharge receives an increased risk for values between 0 and 0.863 and 2.513 and 3.313 mmHg. However, the interpretation of this behavior and determining its clinical implications was impossible. (f) The 2D risk function for age and the IQR of the BE over 3 days is interpretable. Patients over 71.5 years have a high risk for a high IQR of the BE. Patients between 59.5 and 71.5 have only a slightly increased risk for low IQR values and younger patients have a decreased risk across all BE values. However, interpreting this risk function to assess its clinical safety is very hard. The team concluded that it contained no clearly harmful relationships and included it in the model.

We collected qualitative feedback from the team during model inspection (see supplement). A major problem was to draw the line for risk function exclusion. Most functions fulfilled one of the above problems to some extent. The team agreed to only exclude a risk function when a problem was clearly present and there would be a considerable impact for patients, i.e. value ranges with many patients where affected. Still, many functions could be assigned to either category (comments 1-3). The team stated that it was difficult to consider the cohort reduced to a single independent risk function (comments 4-7). This is against clinical practice where several measurements of a patient are integrated. Also, only considering patients at discharge was difficult because usually the whole patient history is taken into account (comment 8). In addition, the team members tended to construct explanations for risk functions without clear evidence (comment 9). Moreover, values outside the usual value ranges and IQR and trend features were more difficult to understand (comments 10, 11). Especially, 2D function posed a problem since the combinations of features were uncommon in clinical practice. Even though it was possible to grasp the content of the risk function, it was very hard to infer its clinical implications leading to exclusion (comment 12). There was a tendency to rely more on the model to derive useful relationships when a risk function was less interpretable (comment 13). In addition to that, we collected general properties that hindered or supported interpretability which confirmed previous findings^31^.

### Performance of EBM model versus four other prediction models

After risk function selection and model inspection, the EBM model contained 67 1D risk functions. It achieved a PR-AUC of 0.119±0.020 and a ROC-AUC of 0.680±0.025 (see Figure 3). For recall values of 0.4, 0.5, 0.6, and 0.8 the precision was 0.130±0.032, 0.111±0.019, 0.105±0.013, and 0.082±0.005. Utilizing the SAPS II in the last 24 hours showed an inferior performance of 0.084±0.025 (PR-AUC) and 0.607±0.019 (ROC-AUC). Also, LR with 130 selected features and the RNN achieved a lower performance with a PR-AUC of 0.092±0.026 and 0.095±0.008 and a ROC-AUC of 0.587±0.016 and 0.594±0.027. Both were placed between EBM and SAPS II for PR-AUC and below SAPS II for ROC-AUC. The latter could be because we optimized for PR-AUC during parameter tuning and variable selection. The GBM trained on all 1,423 features performed similarly to the EBM with a PR-AUC of 0.123±0.016 and a ROC-AUC of 0.665±0.036.

**Figure 3.**
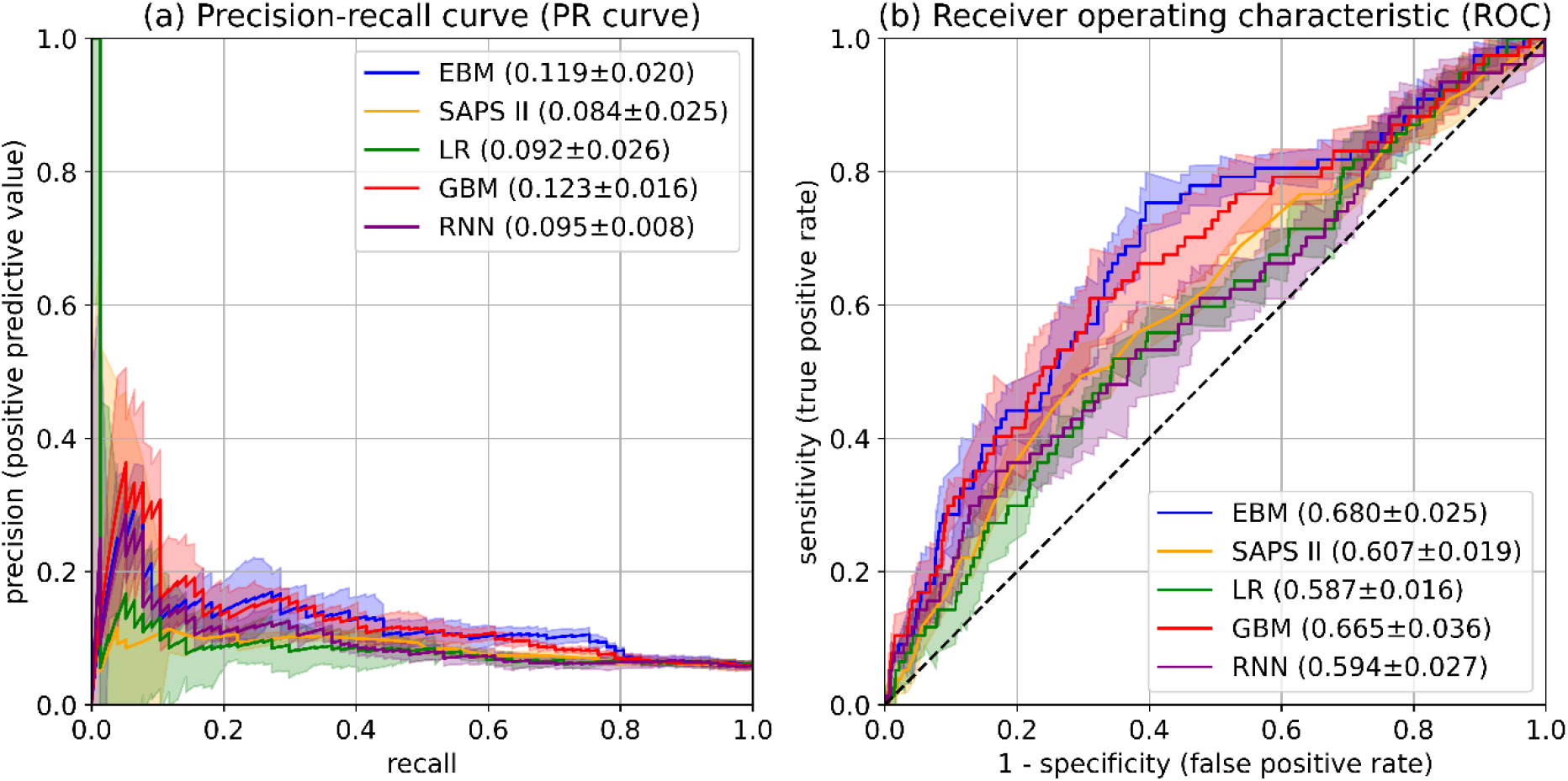
Performance evaluation on UKM cohort. (a) PR-AUC was considered the most relevant performance indicator due to the imbalanced label distribution. We optimized for PR-AUC during parameter tuning and selection procedures for all models. Differences between models are relatively small. EBMs and GBMs show the highest PR-AUC. (b) ROC-AUC was determined as an additional performance measure. Again, EBM and GBM models perform best. (a), (b) Confidence intervals were determined with the standard deviation on five temporal splits. Held-out data was only used for the final experiments.

### External Validation of EBM model on MIMIC-IV

We used the MIMIC-IV database^32,33^ for external validation. It contains 76,540 ICU stays of 53,150 patients admitted to Beth Israel Deaconess Medical Center (BIDMC) between 2008 and 2019. After applying a similar cohort selection and labeling procedure, we included 19,108 ICU stays of which 1,626 (8.5%) were labeled positively. The number of positive instances is considerably higher in the MIMIC-IV cohort. We resampled negative instances to obtain the positive rate of the UKM cohort for performance comparison. We extracted 41 variables used by the EBM model from MIMIC-IV. Only a single variable could not be generated because the relevant variable was not collected. Hence, the final EBM contained 66 1D risk functions. We also performed external validation with the GBM model since it performed best in the model comparison. However, we only used variables of the EBM model because extracting all variables from the MIMIC database was not feasible. Hence, 515 features of 41 variables were used as inputs. EBM and GBM performed very similarly with a PR-AUC of 0.221±0.023 and 0.232±0.029, and a ROC-AUC of 0.760±0.010 and 0.772±0.018 (see supplement).

## Discussion

This study showed that for 3-day ICU readmission prediction a transparent EBM model containing only 67 risk functions performed on par with state-of-the-art GBMs trained on 1,423 features and outperformed RNNs trained on time series data. Both GBMs and RNNs can be considered black box models due to their complexity. We demonstrated how stepwise risk function selection and model inspection through a team of doctors can be used to create an inherently interpretable EBM for a clinical use case. The final model achieved a PR-AUC of 0.119±0.020 and a ROC-AUC of 0.680±0.025. At a recall of 0.5, it reached a precision of 0.110±0.019. Hence, when 50% of all positive labels should be classified correctly every ninth prediction (11%) is a true positive. External validation on MIMIC-IV showed improved EBM results of 0.221±0.023 PR-AUC and 0.760±0.010 ROC-AUC and confirmed that they perform similar to GBMs. Our performance results are consistent with previous studies showing that EBMs outperform LR and are on par with random forests and boosting methods^12,30^. However, in contrast to existing work, adding 2D risk functions lead to lower performance on the held-out data. During risk function selection, we observed a slight decrease of the PR-AUC which could be considered as an accuracy-interpretability trade-off for EBMs. The overall predictive performance for 3-day ICU readmissions was relatively low. This is probably due to limitations regarding data quality, which is supported by the higher performance on MIMIC-IV. MIMIC-IV was created in several iterations and integrated the feedback of many researchers leading to higher data quality. Moreover, ICU readmission prediction is a hard task and only a few readmissions are preventable^34^. Still, we think that an EBM model for 3-day ICU readmission prediction can serve as a useful additional indicator for clinicians at ICU discharge. It can give predictions based on a local cohort, which might offer additional insights for decision-making.

Several studies on ICU readmission prediction exist^26,35–47^ and we identified two systematic reviews^48,49^. Most of them also used MIMIC^33^ for model development or validation, however, not the most recent version IV. Readmission intervals varied from 24 hours to anytime. We chose three days for our experiments since we considered it a good trade-off between including readmissions with relevant conditions and excluding stays with insufficient follow-up periods. Previous studies tested many different models and two^26,27^ mentioned the goal to develop interpretable models, but no validation through humans was performed. All studies reported ROC-AUC which varied from 0.64^47^ to 0.91^37^. Unfortunately, comparing the performance with existing work is impossible due to two reasons. First, we considered PR-AUC due to the label imbalance of ICU readmissions and optimized it in our experiments. However, none of the existing studies reported this performance measure. A single paper contained a precision-recall curve^42^, but no area under the curve. Second, we created a custom UKM cohort and we used MIMIC-IV for external validation. None of the identified studies used this data. If ROC-AUC is considered as a performance measure that we did not optimize for, our results are located at the lower spectrum of the reported models.

A goal of this study was to demonstrate the development of an inherently interpretable ML model and inspect it with clinicians to remove problematic risk functions. This approach showed mixed results. On the one hand, our collaboration with clinicians confirmed that they can easily grasp the concept of EBMs^31^ making them a useful transparent model candidate for healthcare applications^50^. Like LR, which is well known in the medical domain, feature contributions are summed to a total log-odds score. This modularity also allowed to focus on a single risk function at a time, which helped during the inspection. Confidence intervals and histograms over patient densities further helped to assess the relevance of function segments. For instance, it was possible to ignore fluctuations of risk functions in regions with few patients. In addition, our model development process enabled discussions with clinicians and encouraged a critical review of the model. Several aspects were raised for the first time such as the problem with PTT measurements. Hence, EBMs can help to include stakeholders into the development process to establish trust which ultimately could lead to higher adoption rates^13^. Moreover, we identified and removed 18 risk functions due to lack of interpretability, undesirable data artifacts, and contradiction of medical knowledge demonstrating the capability of EBMs to unveil and remove undesirable components. This would have been impossible with a black box ML model^10,12^. Lastly, model inspection led to a performance increase on the held-out data suggesting better generalization. However, we also observed several shortcomings regarding the interpretability of EBMs for 3-day ICU readmission prediction. During model inspection 33 of 85 were labeled as problematic and 17 thereof as not interpretable. Reducing a patient cohort to one or two features and considering a fixed time interval before discharge is against clinical practice where usually many variables are integrated over a long time horizon. So, it often proved hard to create an intuition about patient groups that would be affected. Also, for meaningful interpretation of EBMs, it is necessary to understand the model inputs^23,50^. Especially, variables and descriptive statistics that were less common in clinical practice hindered interpretability. It could be an option to let clinicians pick interpretable features a priori. In addition, shapes of risk functions sometimes showed fluctuating behavior^31^. We already increased the bin size to prevent those artifacts, but some still occurred in the final model. Another major issue was to draw the line between inclusion and exclusion of risk functions. Most functions showed some problematic behavior. So, we decided to exclude only function with a problem that affected a considerable part of the cohort. However, this decision rule is vague, and we expect low interrater reliability. We think it could be helpful to have a clear application scenario to determine more specific rules for exclusion. Moreover, we observed that it was harder to justify the exclusion of less interpretable functions and the team relied on the EBM algorithm to find relevant associations in the data^51,52^. We conclude that EBMs showed several useful properties for transparent model development. However, they are no panacea for ML in healthcare^53^.

This work has limitations. Even though ICU readmission prediction is a relevant medical problem, it can be difficult to turn predictions into actions when institutional factors such as missing ICU beds must be considered. No multicenter cohort was used for the development and validation of our prediction model, so the external validity of our results is low. Also, the data quality of the local cohort was limited and our experiments only focused on a single interpretable model. External validation on MIMIC-IV was only performed for two models and there was no in-depth analysis for the improved performance. Moreover, interpretability should be evaluated in the context of its end-task^14^. Ideally, this could be increased trust leading to higher adoption of the system or even improved patient outcome. We limited our analysis to prediction performance, identification of problematic risk functions, and qualitative feedback. Moreover, there was no rigorous set of rules for model inspections so that it would probably show low interrater reliability. Lastly, automatic risk function selection for EBMs might have removed important confounders.

Further experience with EBMs and other interpretable ML models in the healthcare setting is necessary. Different transparent models could yield an improved performance or prove more suitable regarding interpretability. Extending beyond ICU readmission prediction, it would be useful to obtain an evaluation of interpretable ML approaches for a wide variety of medical tasks as guidance for practitioners^14^. Also, translating an interpretable ML system into the clinical workflow and analyzing its effect on e.g. adoption rate or clinical outcome could yield valuable insights on their effectiveness. It is also important to determine the limitations of interpretable ML^53^ and explore possible alternatives that might be more suitable^54^. Moreover, future work could consider improvements of the EBM algorithm to increase its interpretability. Especially, further risk functions constraints such as smoothness and an improved risk function selection could be beneficial.

## Methods

### Study setting and preregistration

We conducted a retrospective cohort study based on ICU patients managed by ANIT-UKM. The department supervises four ICUs for surgical patients with critical conditions and perioperative patients with 40 beds and around 3,200 admissions per year (see supplement). Administrative data were obtained from the HIS of the UKM (ORBIS, Agfa HealthCare) and ICU data from the PDMS (Quantitative Sentinel, GE Healthcare) managed by the ANIT-UKM (see Figure 1). All analyses and computations were performed on a secure server within the network of the UKM. The ethical review board of the medical chamber Westfalen-Lippe approved this study (reference number: 2020-526-f-S). Preregistration for this work was registered online before carrying out the experiments^55^. Data processing was not included in the preregistration due to its dependence on the data. There were two deviations from the preregistration: a readmission interval of three instead of seven days was considered. We decided to require a follow-up period at the UKM of the same length as the readmission interval. Hence, an interval of seven days would have led to exclusion of many ICU stays. Also, we denoted that all experiments will be conducted on both datasets. However, this statement was formulated too broad and we used MIMIC-IV only for external validation. We provided the transparent reporting of a multivariable prediction model for individual prognosis or diagnosis (TRIPOD) checklist for prediction model development and validation^56^ in the supplement. All code for data preprocessing and our experiments is publicly available^57,58^.

### Cohort

We aimed to include all ICU stays managed by the ANIT-UKM that were discharged to standard care (hospital ward) within the same hospital and had a follow-up period of at least three days. The analysis was restricted to a single department to facilitate data acquisition and to obtain a homogeneous study population. Initially, all ICU and IMC transfers of all adult patients at the UKM between 2006 and 2019 were retrieved from the HIS. This data contained patient identifiers, case identifiers for each hospital stay, and start and end times of the hospitalization and ICU stays. We chose 2006 as a starting point since it was the first year that all included ICUs used the same PDMS and HIS for data collection.

The flow chart in Figure 4 illustrates the exclusion criteria for the cohort. First, correct formatting of the data was verified and, if possible, missing hospital discharge dates were derived from other transfer entries. 283 entries had to be removed due to ambiguous discharge dates, overlapping hospital stays, or overlapping transfers that could not be repaired. Next, transfers not managed by the ANIT-UKM (n=101,243) and IMC transfers (n=39,165) were excluded. A transfer entry can indicate admission to a different ward or a relocation within the same ward (e.g. for a room change). For the study, we aimed at merging consecutive transfers at included ICUs into a single ICU stay. Usually, transfers were stored in the HIS with no delay, so the discharge and admission times of two subsequent transfers were equivalent. We merged these transfers (n=26,246) in step four. However, there were also some artifacts in the data where short intervals occurred between two transfers even though no discharge occurred. To distinguish these artifacts from true readmissions, we manually inspected 147 consecutive transfers with a non-zero interval of at most twelve hours. We designed a stepwise algorithm based on the identified artifacts to decide if a discharge occurred or not. However, 19 transfer pairs remained that we inspected manually with the help of all available data in the PDMS including clinical notes. In steps five and six, we excluded stays that were irrelevant for the prediction since they ended with the death of the patient (n=2,327) or they were discharged to an external ICU or IMC unit (n=10,688). To distinguish consecutive transfers and readmissions to an external ICU, we used the same procedure as in step four. We identified 67 consecutive transfers with a non-zero interval of at most twelve hours and of those we inspected 29 manually. Next, we excluded all ICU stays which were discharged from the hospital within 72 hours to ensure that patients with worsening conditions are transferred to an observed ICU in the UKM (n=3,975). This also excluded stays that were transferred to an external facility or home, which might be unknown at discharge time from the ICU and, hence, introduces a selection bias in the cohort. However, we reckoned that ensuring a complete observation interval outweighed this effect. Lastly, we removed implausible cases that contained no age entry (n=63), had less than two hours of heart frequency recordings (n=137), or had no heart frequency recordings in the last six hours of the stay and no discharge information in the PDMS (n=63). The discharge times of all remaining stays (n=15,589) were set to the last heart rate recording giving a more precise time for a patient’s actual discharge.

**Figure 4.**
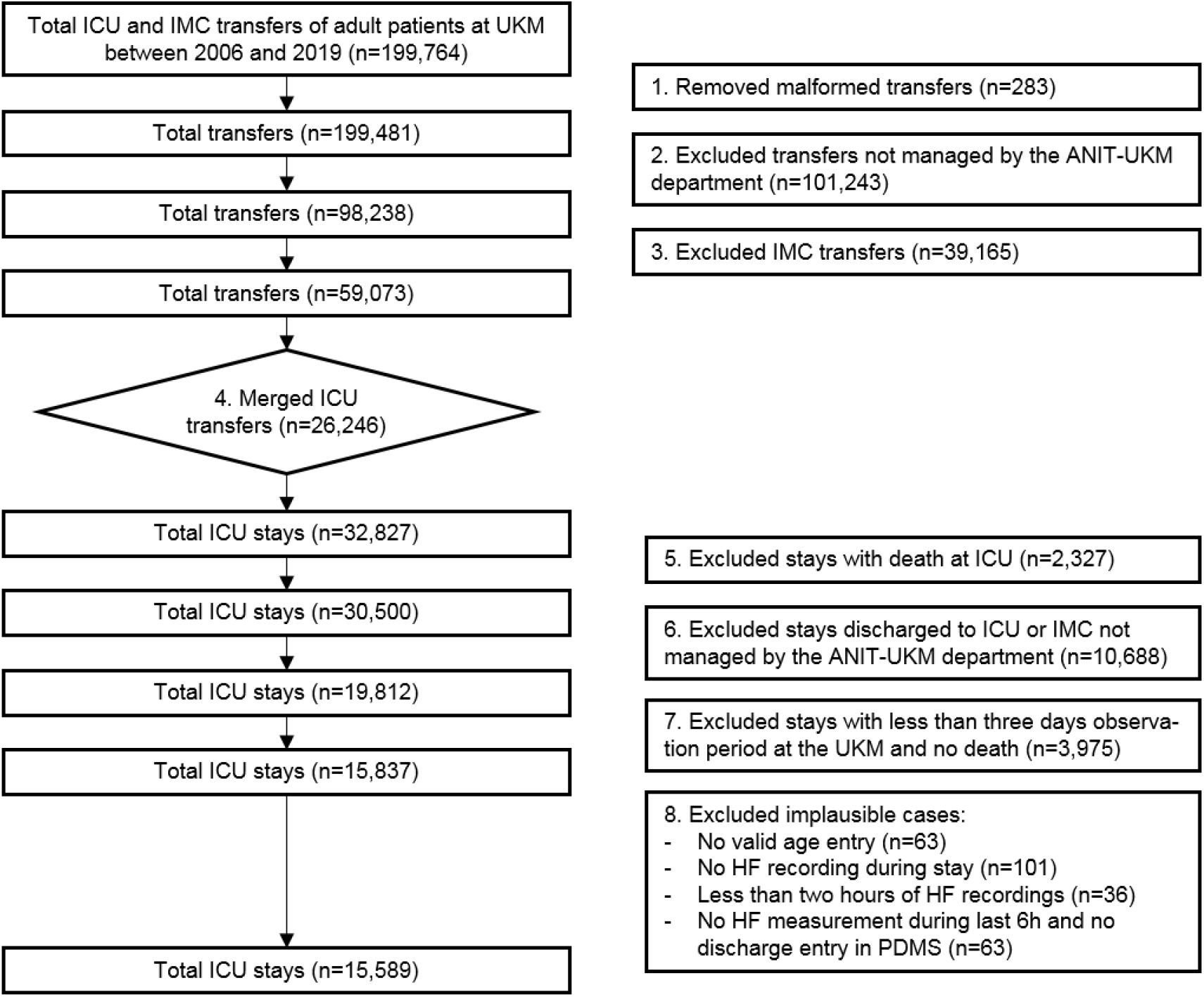
Cohort selection for UKM cohort. Transfers at ICU and IMC wards between 2006 and 2019 at the UKM served as initial data. We included four ICUs managed by the ANIT-UKM department. It was necessary to merge transfers including a manual procedure to obtain consecutive ICU stays. Death at the ICU and discharge to an external ICU or IMC were excluded. We required an observation period of at least three days to ensure readmission to an ICU within the UKM. Lastly, implausible cases were removed.

### Determining ICU Readmissions

ICU stays that were discharged to standard care (general ward) and were readmitted to any ICU (n=822) or IMC unit (n=31) or died within three days (n=38) in the UKM were labeled as true (n=891). All remaining stays received a negative label (n=14,698). All ICUs and IMC units were considered for readmission because patients with worsening conditions were not exclusively re-admitted to ICUs managed by the ANIT-UKM. Patient deaths were labeled as true to have a consistent label for negative outcomes. An interval of three days was used because we considered it reasonable that patients would be readmitted to an ICU due to a medical condition that may have already been present at discharge. Also, most patients had a three days observation after ICU discharge and previous studies also used it. Note that patients that were discharged to standard care and underwent a planned procedure with a subsequent re-admission to an ICU or IMC units within three days incorrectly received a positive label. However, we were unable to develop a reliable procedure to identify such cases. While this might lead to a bias in the data, we consider this a small effect.

To verify our cohort selection and labeling procedure we sampled 20 positive stays stratified across wards. We used all available clinical information including notes to determine if readmission occurred within three days. For one stay the available data did not suffice for a clear decision, so we removed it (step 1 in Figure 4). Since an automatic detection of this case was impossible, we did not change our data processing based on this finding. A figure in the supplement shows three example hospital stays to illustrate different situations and the according labels.

### Variables

We included all routinely collected variables that could be relevant to predict ICU readmissions and that had a sufficient data quality. A flow chart in the supplement illustrates the selection process. Initially, we included all variables defined in the PDMS (n=6,469). First, we removed all variables that were not collected during the study period (n=1,322). We excluded derived variables (n=1,029) computed by formulas in the PDMS since we only wanted to use the original data for the analysis. We also excluded text variables and clinical notes (n=777) since most of them had a very heterogeneous data quality during the study period and we considered them as too unreliable. Next, we scanned all variables and excluded clinically irrelevant ones (n=1,398) such as device-specific or billing variables. Together with a technical expert of the PDMS we discussed open questions and determined additional irrelevant variables (n=581). The remaining 1,362 variables were processed in consultation with a medical expert (a senior physician at the ANIT-UKM) having profound experience with the PDMS. They contained 802 non-medication entries and 560 medications, which were treated separately. For non-medication variables, we determined the coverage across the study period and generated descriptive statistics. Medical concepts collected across variables were considered as a single variable, e.g. blood pressure was collected across several variables and they were merged for the analysis. We used this information together with knowledge about the clinical relevance and the collection process to exclude 522 variables. This resulted in 280 variables of which 70 were included directly and 210 were further processed and merged into 50 variables yielding 120 included non-medication variables. For medications, we assigned World Health Organization Anatomical Therapeutic Chemical (ATC) codes to all entries. ATC codes have a hierarchical structure and we determined 44 clinical useful and relevant medication categories. 187 medication variables were not assigned to any category and were excluded. Additionally, we created five medication variables for therapeutic and prophylactic antithrombotic agents and equivalence dosages of cardiac stimulants, norepinephrine and dopamine, and glucocorticoids, which we considered clinically relevant. This led to 49 included medication variables. An overview of all variables is given in the supplement.

### Data Cleaning

We developed a data pipeline consisting of preprocessing, merging, filtering, and postprocessing. Treatment of duplicates with identical entries and numerical values was performed first (see Figure 1). Preprocessing methods were applied to the raw recordings and could be reused between different items. They also included datatype-specific routines for continuous and categorical variables. Merging was optional and usually needed a custom merging procedure to account for different data formats. Filtering allowed to enforce an interval or a set of allowed values. Lastly, postprocessing was applied analogously to preprocessing. The most important data cleaning procedures are summarized below.

#### Duplicates for non-medication and non-fluid variables

We used a similar approach as^15^ to treat duplicates. On a global level, we removed all recordings of a patient with identical timestamps and values. In case the values differed, we distinguished categorical and numerical variables. For categorical variables, we removed all recordings since we considered them as malformed. For numerical variables, we used the mean of all duplicates when the SD of the duplicates was <5% of the SD of the variable across all patients. Otherwise, we also removed the duplicates. After merging variables, new duplicates might occur in the recordings. For categorical variables, we handled duplicates in the custom merging procedures. For numerical variables, we used the median values.

#### Valid values for non-medications

For most variables, an interval or set of valid values was specified (see supplement). For continuous variables, lower and upper bounds in the PDMS served as a starting point but were narrowed in some cases. For categorical variables, lists for permissible values in the PDMS were used to construct sets of valid values. We checked all values outside these sets and detected several malformed recordings, which were due to manual data entries. Whenever a valid value could be determined for those, we mapped them accordingly. Value sets were sometimes reduced to more general categories when it seemed more appropriate.

#### Valid values for medications

The medication variables contained many artifacts which made it impractical to use dosages as variables. Instead, we only used indicators if a certain medication was administered. To this end, we removed zero and negative entries. We added three medication categories with dosages. We determined valid maximum values during the medical review of the variables and removed all entries above them.

#### Adjusting body temperature for measuring site

Based on the measuring site, body temperature measurements were adjusted for the core temperature. We applied an offset of 0.4°C for tympanic, 0.5°C for oral and, 0.6°C for axillary sites^59^. Groin and axillar show similar behavior, so an offset of 0.6°C was used for groin^60^. Since no evidence could be found for the nasal site, we used the same offset as for oral.

#### Adjusted blood pressure for measuring site

Non-invasive systolic, mean, and diastolic blood pressures were adjusted for measurement at arm or thigh. We used offsets of 7, 4, and 3 mmHg for arm and -5, 6, 11 mmHg for thigh^61^.

#### Missing values

We used value imputation only for some variables to incorporate the missingness of variables in the model. However, when only a few values were missing or missingness indicated a normal value, we imputed them. For gender, a single missing value could be derived from another source. For static variables patient class and responsible clinic, we used “ inpatient” and “ other”. Missing Glasgow Coma Score or Richmond Agitation-Sedation Scale indicate a normal value in the clinic, so we used them for imputation. For 29 time-series features with few missing values, we used the median value.

#### Computation of estimated glomerular filtration rate (eGFR)

The eGFR in the PDMS was based on different formulas. We used creatinine, gender, and age to calculate it as a new variable^62^.

### Features

We assigned each variable to one of eight feature classes that determined which features were generated (see supplement). For static variables, we used the last entry in the given time interval. For time series variables including flows and medications, we generated descriptive statistics for different time intervals, since EBMs cannot integrate them directly. We defined three types of time series features and assigned them according to the median sampling interval of a variable. For merged variables, we used the minimum sampling interval of all merged variables. All of them generated different descriptive statistics for three different time horizons before discharge. Note that for a given time horizon all data of a patient collected during this period at an included ICU were used. For continuous variables, these are minimum, maximum, interquartile range, median, and trend leading to 15 features. Flow and medications features shared the same time horizons as the time series feature for low sampling frequencies. For flows, the mean daily input/output is determined, which is extrapolated if the stay is too short. Medication dosages could not be used due to insufficient data quality. Instead, an indicator if any substance of a medications group was administered and the number of different substances was used. We did not use the whole stay as a time interval because only 10.18% of all included stays lasted longer than seven days and we reckoned that data closer to discharge is more relevant. So, only the seven days before discharge are considered for feature generation. Lastly, we generated two features for interventions indicating whether it was performed at all and for the time interval since its last performance. Five features could not be determined by these predefined types and were created manually. They are described in the supplement. In total 1,423 features were generated.

### Explainable Boosting Machines

EBMs are GAMs with additional shape functions for variable interactions. Standard GAMs^63^ predict a label *ŷ* that can be transformed by a link function *g* with the summation of features transformed by shape functions *f*_*i*_(*x*_*i*_) and a bias term *β*_0_ (see equation (1)). EBMs add shape functions for interactions of two features *f*_*i,j*_(*x*_*i*_, *x*_*j*_)^30^ and for dichotomous classification, the logit link function is used analogously to LR (see equation (2)).

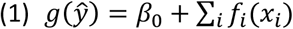

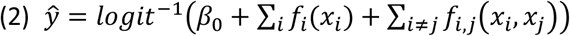

The shape functions of EBMs are also called 1D and 2D risk functions in this study. Different methods to estimate them exist^29,63^. EBMs use boosted decisions trees that allow versatile function shapes and showed optimal performance^29^. By visualizing learned risk functions EBMs can be inspected, and due to their modularity, inappropriate functions can be removed. Hence, they can be considered as decomposable transparent models^23,50^. Also, for a given input contributions of each risk function can be used as an explanation of a prediction. Even though EBMs are transparent models, their performance is comparable to Random Forests and Boosting^28,30^. A study applying them in two healthcare tasks highlighted their potential to identify and remove spurious correlations^12^. Moreover, an evaluation revealed that medical doctors can grasp the concept of EBMs and feel confident in working with them^31^ making them a suitable model for our experiments.

### Creation of EBM model with a limited number of risk functions

The goal was to train an EBM model with at most 80 risk functions. Together with clinicians, we defined this as the maximum amount to make interpretation feasible in a reasonable amount of time. First, we determined model parameters used during risk function selection. We did this in three steps: we performed parameter tuning on all features, then, we estimated the 80 most important risk functions and then performed another parameter tuning for these 80 risk functions. Parameter tuning was performed with a complete grid search over 490 parameter settings (see supplement). We tried to include relevant parameters and used prior experiments to narrow down the value ranges. For each setting, the EBM model was trained on all features of the full training set. We chose the parameters that led to the highest PR-AUC score on the full validation set. Next, we estimated the 80 most important risk functions based on a random 85% training and 15% validation split of the training and validation sets. We had to use a random split instead of the original validation set (all data from 2018) because a couple of variables were only collected for some years (see Figure 5). This led to a biased weight estimate when using training and validation data based on years. Importance was calculated with the mean absolute log-odds score assigned by a risk. Lastly, we performed another parameter tuning step with reduced sets of 80 risk functions to obtain the final parameters for risk function selection.

**Figure 5.**
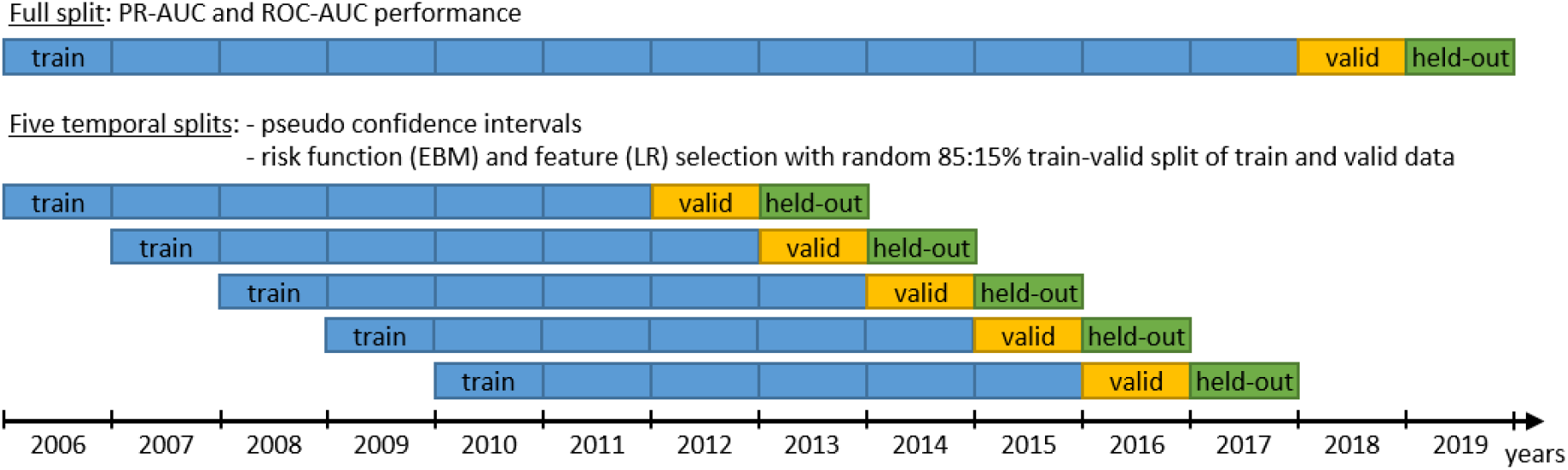
Data splits for the experiments. The full split utilizing all data was used to determine the performance in our experiments. We used data from 2018 and 2019 as validation (valid) and held-out sets to simulate predictions made on future data. Five additional temporal splits were used for confidence intervals and risk function (for EBM model) and feature (for LR model) selection^15^. We optimized the performance on the validation split in our experiments. Held-out data was only used for the final results reported in the manuscript.

To evaluate the effect of different bin sizes of the EBM model, we determined optimal parameters for each bin size and performed a separate risk function selection. We carried out a separate selection procedure for each bin size because prior experiments showed that the selected risk functions highly depend on the bin size.

Risk function selection was performed in a greedy stepwise forward selection process on the five temporal splits. In each round, we added the risk function or feature with the largest average importance. Importance was determined on a random 85% training and 15% validation split as explained above. In each round, we evaluated the model on the full split. We then chose the EBM bin size and selection of risk functions that yielded the best PR-AUC performance on the validation set. For EBM models we only considered 1D risk functions until this point. We added a second selection step for 2D risk functions on the features of the included 1D risk functions. This is coherent with the EBM training algorithm, which first trains 1D functions and then adds 2D functions for the residuals. We performed the same greedy stepwise forward selection process as for 1D risk functions and chose the set of 2D risk functions that led to the biggest PR-AUC improvement on the full validation split.

### Inspection of EBM model through a team of doctors

The goal of EBM model inspection was to identify risk functions that should not be part of the final prediction model. The value ranges of several risk functions were adapted to reduce the distortion due to outliers. We inspected the model in a team of three individuals: a senior physician working at the included ICUs, a senior physician responsible for the data infrastructure at the ANIT-UKM, and the developer of the EBM model with a computer science and medical background. We discussed and determined potential problems of the risk functions a priori to agree on a common set of exclusion criteria. For each risk function, we first described its shape and agreed on its content. Next, we determined if any of the identified problems were applied and denoted them in a protocol. Lastly, we decided if the problems justified the exclusion of a risk function. Hence, risk functions were assigned into three classes: included without problems, included with problems, and excluded. We collected the identified problems and classes for all risk functions (see supplement). In addition to that, we collected qualitative feedback EBM model inspection.

### Models for Performance Comparison

For the SAPS II model, we calculated the SAPS II for the last 24 hours of each stay and used the score as a prediction. SAPS II was developed and validated to predict mortality in the ICU based on the first 24 hours of a stay^64^. However, in the included ICUs it is calculated and manually validated every day. Hence, we think it is reasonable to consider the current SAPS II for our experiments. We included the SAPS II to evaluate the potential of a validated clinical score. Due to its simplicity, we expected a rather low performance for a weak baseline. Other ICU scores exist^65^. However, since the included ICUs only collected SAPS II variables consistently during the study period, we decided to use SAPS II in our experiments. Previous studies also evaluated its association with ICU readmissions^48,66^.

We aimed for an LR model with similar complexity and, hence, a comparable level of interpretability as the EBM model. To this end, we performed feature selection analogously to risk function selection for EBMs. Since LR cannot handle categorical data and unknown values, we added 106 dummy variables for 39 categorical variables and 856 unknown indicators which increased the number of features from 1,423 to 2,346. This corresponds to an increase of 65% so that we aimed for 130 instead of 80 features for the LR baseline model. Feature importance for LR was determined via mean absolute feature contribution on z-normalized data. GBMs served as a strong performance baseline trained on all features. We used the widely used XGBoost library with a parallelized implementation for our experiments^67^. We replaced 39 categorical variables with 106 dummy variables. An RNN was used as another strong baseline model that could incorporate raw time series data^68^. Hence, the variables were used directly as inputs without generating features. For time series variables (n=152), we limited the time interval to hospital stays or at most 21 days before discharge and resampled all data to one-hour intervals to reduce the input size. A fraction of 8.23% (n=1283) stays was longer than 21 days. We padded all input series to 21 days using unknown values, imputed variables at each timestamp, forward filled unknown values. The minimum value minus one was used as an unknown indicator. For static variables, we added dummy variables for categorical variables (n=15) and unknown indicators (n=3). Parameter tuning was performed for all models (see supplement). We chose the configuration with the highest PR-AUC on the validation data. Some parameter settings caused erroneous PR-AUC values due to few recall steps in the PR curve. We filtered these models by looking at the ROC-AUC to verify whether the mode learned any useful relationships. Particularly, for RNNs this effect was strong, and we used a cutoff of 0.55 ROC-AUC to filter valid runs of parameter tuning.

### External Validation on MIMIC-IV

We performed an external validation of the EBM and the GBM model on the MIMIC-IV database version 1.0 ^32,33^. MIMIC-IV consists of 76,540 stays of 53,150 patients admitted to an ICU at BIDMC between 2008-2019. It contains all data collected at the ICUs making it a good candidate for external validation. We used code from the shared code repository to load the data and generate medical concepts.

We tried to mimic the cohort selection as close as possible. MIMIC-IV already provided consecutive ICU stays as so-called concepts so that merging of transfer was not necessary. Especially, the manual procedure to classify gaps between ICU stays was irrelevant for MIMIC-IV. The flow chart in the supplement shows the cohort selection. First, 104 malformed ICU stays of 76 patients were excluded due to hospital discharges before admission, overlapping hospital stays, and a time of death before ICU admission. Second, stays that were not discharged from an ICU managed by the Department of Anesthesia, Critical Care, and Pain Medicine at BIDMC were excluded (n=43,154). The included ICUs were Trauma Surgical Intensive Care Unit, Surgical Intensive Care Unit, Cardiovascular Intensive Care Unit, and Neuroscience Intensive Care Unit. Next, we excluded all stays who died during the ICU stay (n=1,888). This included two stays that were transferred to an ICU after death which we also considered as death at ICU. Analogously to the original cohort, we required a length of stay at the hospital after the ICU of at least 72 hours (n=12,187). The ratio of the excluded cases was considerably higher than in the original cohort. Lastly, we removed cases that had no heart frequency entries for at least two hours (n=99).

We also labeled discharges from an included ICU to standard care as true when a patient was readmitted to any ICU or IMC units or died within three days. For the UKM cohort, we designed a special procedure and performed manual annotation for transfers of at most twelve hours. However, this was not feasible for the MIMIC-IV cohort because we lacked clinical knowledge about the data. Hence, we used a simplified procedure that required stays at a standard care unit or consecutive readmission to an ICU or IMC unit to last at least one hour to prevent artifacts. Also, we excluded the post-anesthesia care unit and unknown from the standard care units because they could indicate a planned surgery leading to readmission to an ICU or IMC unit. BIDMC had more IMC units so that fewer ICU stays were directly discharged to a standard care unit. Hence, we expected a lower fraction of positive labels. However, 1626 ICU stays were labeled as readmission or death. Of those, 1273 were readmitted to an ICU, 39 to an IMC unit, and 314 died within three days. We controlled 20 random positive labels stratified by ICUs to verify the labeling procedure.

We extracted 41 variables for the EBM model from MIMIC-IV. We also used the data collected during ICU stays. We searched through the item definitions and the provided code to identify relevant items. Analogously to the UKM cohort we defined allowed value ranges and applied median value imputation. Only the variable procalcitonin was not contained in MIMIC-IV. As a result, 66 features were generated for the EBM and 515 features for the GBM model. The same code was used for feature generation. Less effort was put into data cleaning compared to the UKM cohort. However, we expected that the MIMIC-IV database has better data quality data since it is publicly available, and the developers integrated the feedback of several researchers.

Variables were collected differently in the MIMIC cohort. For instance, blood loss was collected less frequently. Hence, we deemed it necessary to retrain the EBM model for this task. We used the same parameter setting determined on the UKM cohort. For a proper performance comparison, we resampled negative cases in the MIMIC-IV cohort to obtain the same ratio of positive cases. MIMIC-IV only provides intervals of three years for each ICU stay to protect the patient’s privacy. Hence, it was impossible to use the same temporal splits. Instead, we used data between 2008 until 2016 as training set and data from 2017 to 2019 as held-out split. For confidence intervals, we used five random 85:15% splits of the data between 2008 and 2016 for training and testing.

## Data Availability

Data of the UKM cohort used to develop the ML models cannot be published due to privacy concerns. To access MIMIC-IV it is necessary to complete an online course on data usage and a data use agreement must be signed. Code for data preprocessing and all experiments publicly available:
https://doi.org/10.5281/zenodo.5627167,
https://doi.org/10.5281/zenodo.5541443.

https://doi.org/10.5281/zenodo.5541443

## Data Availability

Data of the UKM cohort used to develop the ML models cannot be published due to privacy concerns. To access MIMIC-IV it is necessary to complete an online course on data usage and a data use agreement must be signed. Code for data preprocessing and all experiments publicly available^57,58^.

## Acknowledgments

Not applicable.

## Author contributions

S.H. designed the study, developed the code for all experiments and wrote the manuscript. S.H., T.V., and C.E. carried out the data preprocessing, cohort selection, and experiments. All authors provided critical feedback and helped shape the research and manuscript.

## Competing interests

The authors declare that they have no conflict of interest.

## Notes

### Competing Interest Statement

The authors have declared no competing interest.

### Funding Statement

Financial Support by German Research Foundation (Deutsche Forschungsgemeinschaft, DFG grant DU 352/11-2).

### Author Declarations

Ethical review board of the medical chamber Westfalen-Lippe gave ethical approval for this work (reference number: 2020-526-f-S).

