## Supplement for "Development and Validation of an Interpretable 3-day Intensive Care Unit Readmission Prediction Model Using Explainable Boosting Machines"

#### TRIPOD Checklist: Prediction Model Development and Validation

Will be added as a PDF for the final publication, because page numbers might differ. We ensured that all items are fulfilled.

#### Cohort

*Table 1. **Detailed information of included ICUs.** All ICUs are managed by the ANIT-UKM. The number of patients and transfers in rows six and seven corresponds to the input of cohort selection. The last two rows represent the UKN cohort. Note that they do not sum up to the total number of patients and ICU stays because one patient or stay can be associated with more than one ICU.*

| ICU | ICU 1 | ICU 2 | ICU 3 | ICU 4 |
|---|---|---|---|---|
| Full name | Intensivtherapie I | Intensivstation des Herzzentrums | Intensivtherapie II | Perioperative Anästhesiestation |
| Short name | 19A OST | 19B OST | ANAES INT 2 | ANAES PAS |
| Description | Surgical intensive care unit | Surgical intensive care unit | Surgical intensive care unit | Perioperative intensive care unit |
| Data in PDMS since | 2/1/2001 | 11/1/2005 | 5/1/2003 | 2/1/2001 |
| Number of beds[1] | 11 | 11 | 11 | 7 |
| Number of patients 2006-2019 | 7,389 | 10,372 | 6,231 | 17,061 |
| Number of transfers 2006-2019 | 9,422 | 13,988 | 9,368 | 26,296 |
| Number of included patients | 4,008 | 5,962 | 1,031 | 10,644 |
| Number of included ICU stays | 4,008 | 7,576 | 1,354 | 16,233 |

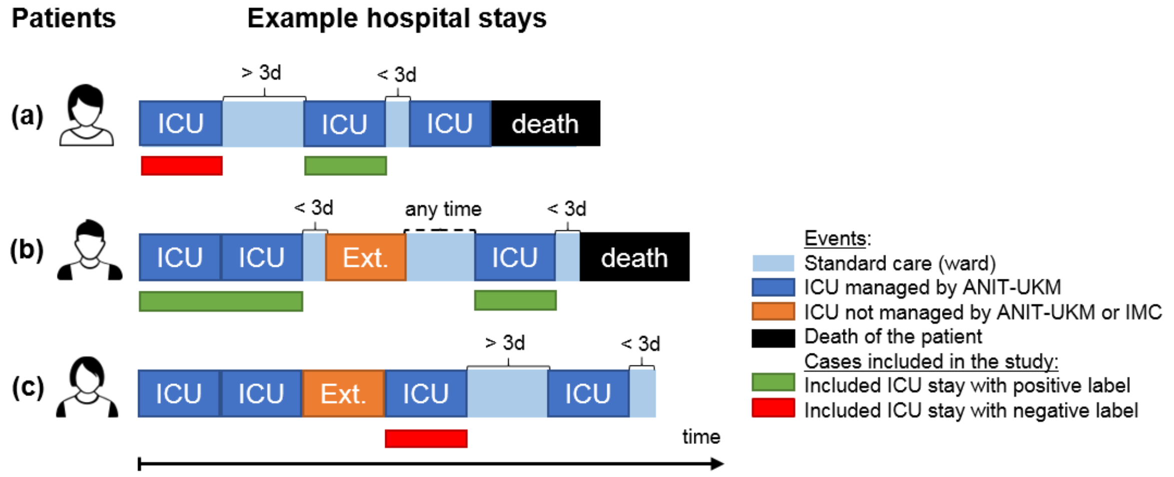


*Figure 1. **Three exemplary hospital stays with according ICU stays and labels.** The event sequence is represented with colored blocks. Included ICU stays have a green (positive label) or red (negative label) bar below them. Transfers at excluded ICUs or IMC wards (orange) or standard care (light blue) were never included in the study, so there are no bars below them. However, readmission to an excluded ICU or IMC ward after discharge from an included ICU was considered for labeling. (a) The first two ICU transfers are included as negative and positive instances, because readmission happened after three days and within three days, respectively. The third ICU transfer is excluded due to death at the ICU. (b) The first two ICU transfers are consecutively, hence they are merged and considered as a single stay. It gets a positive label because readmission to an excluded ICUs or IMC ward occurs within three days. The last ICU transfer is also labeled positively because the patient dies within three days. (c) The first two ICU transfers are not included because they are merged but directly followed by a transfer to an excluded ICUs or IMC ward. However, the subsequent transfer at an included ICU is included with a negative label. The last ICU transfer is excluded because the follow-up period within the hospital is less than three days.*

[1] These are estimated average values because the number of beds changed several times over the years.

### Variables

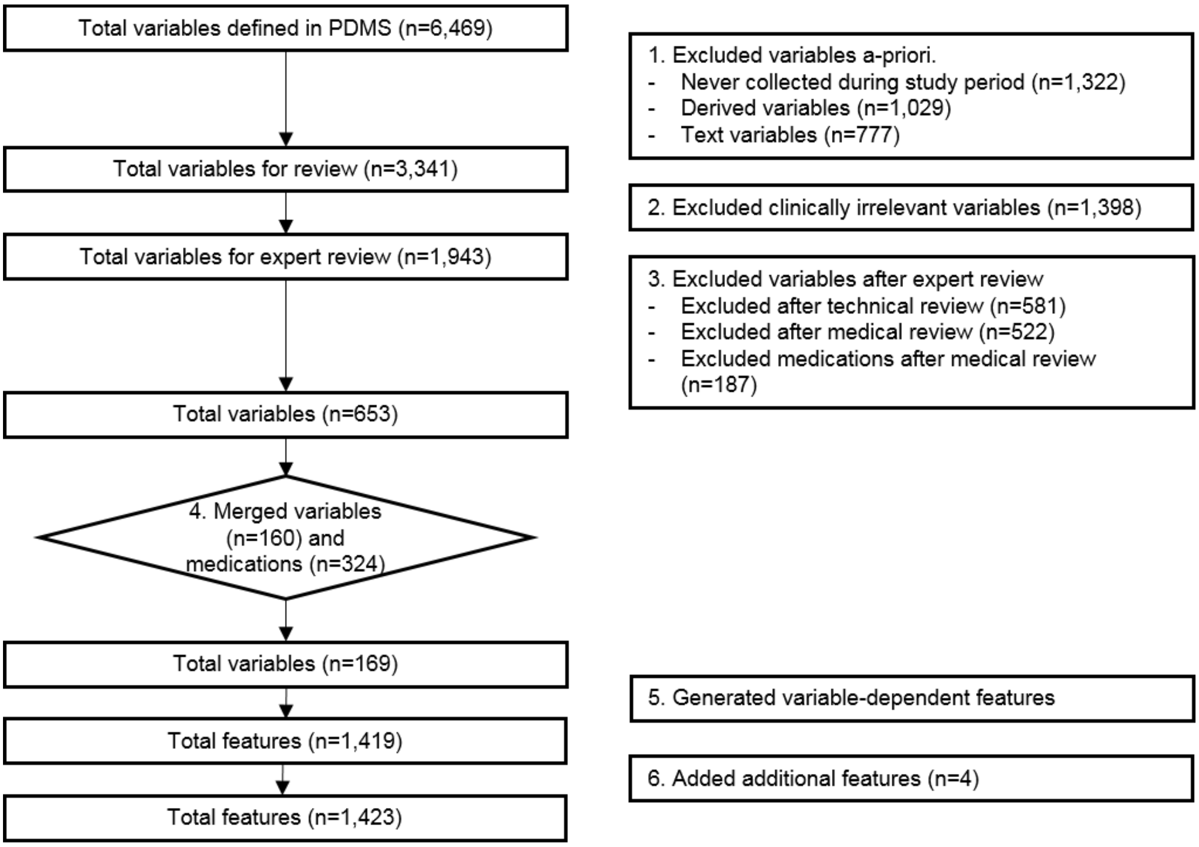


*Figure 2. **Variable selection and feature generation for UKM cohort**. First, 3,128 variables were excluded a-priori. Next, we reviewed the variables for their clinical relevance (n=1,398). During the technical and medical review, we excluded 581 and 709 further variables. Many variables collected the same data, so we merged 160 non-medication and 324 medication entries. The remaining 169 variables (Table 2) were assigned into feature classes (Table 3) from which we generated 1,419 features. Four additional static features (Table 4) were added afterwards.*

*Table 2. **Overview of included variables grouped by feature classes**.*

| Variable name | type | unit | Non-missing values in feature interval | median sampling interval | statistics during feat interval | Missing values in feature interval | Imputation value |
|---|---|---|---|---|---|---|---|
| **Feature class: static per patient** | | | | | | | |
| AIDS | categorical | | 3 | 2 d 09:37:00 | True (n=3) | 15584 | - |
| Age | continuous | years | 15639 | 5 d 07:09:30 | median: 66.00, min: 18.00, max: 112.00 | 0 | - |
| BMI | continuous | kg/m2 | 28261 | 3 d 00:00:00 | median: 27.17, min: 11.30, max: 73.28 | 482 | - |
| Gender | categorical | | 15405 | 03:03:00 | Male (n=10519), Female (n=4886) | 0 | - |
| Height | continuous | cm | 14709 | 02:47:00 | median: 173.00, min: 63.00, max: 210.00 | 448 | - |
| Hematologic malignancy | categorical | | 74 | 2 d 09:55:00 | True (n=74) | 15516 | - |
| Metastatic cancer | categorical | | 651 | 2 d 11:55:00 | True (n=651) | 14964 | - |
| Weight | continuous | kg | 29809 | 2 d 21:13:00 | median: 81.40, min: 30.00, max: 227.00 | 141 | - |
| **Feature class: static per hospital stay** | | | | | | | |
| Admission origin | categorical | | 17402 | 2 d 19:43:30 | operating room (n=14258),<br>general ward (n=1542),<br>other hospital (n=781),<br>emergency (n=306),<br>heart alarm (n=177),<br>ICU anesthesiology (n=133),<br>observation station surgery (n=101),<br>ICU non-anesthesiology (n=61),<br>intermediate care (n=43) | 377 | - |
| Admission reason | categorical | | 16197 | 2 d 11:54:00 | Cardiovascular disease (n=11093),<br>Neoplasm (n=1761),<br>Neurological disorder (n=1141),<br>Respiratory disorder (n=943),<br>Trauma or bleeding (n=572),<br>Sepsis or infection (n=300),<br>GI disorder (n=247),<br>Other (n=140) | 1132 | - |
| Hygienic precautions | categorical | | 525 | 2 d 22:25:30 | True (n=525) | 15205 | - |

| | | | | | | | |
|---|---|---|---|---|---|---|---|
| Patient class | categorical | | 15557 | 14:47:00 | Inpatient (n=14129),<br>Emergency (n=1428) | 13 | Inpatient |
| Responsible clinic | categorical | | 14570 | 1 d 10:52:30 | Cardiothoracic surgery (n=10788),<br>Thoracic surgery (n=873),<br>Orthopedic surgery (n=690),<br>Trauma surgery (n=519),<br>Neurosurgery (n=477),<br>Other (n=346),<br>General surgery (n=244),<br>Otorhinolaryngology (n=197),<br>Gynaecology (n=173),<br>Urology (n=156),<br>Oral and maxillofacial surgery (n=107) | 74 | Other |
| Type of admission | categorical | | 16938 | 2 d 10:09:00 | Scheduled surgical (n=12684),<br>Medical (n=2493),<br>Unscheduled surgical (n=1761) | 492 | - |
| **Feature class: static per ICU stay** | | | | | | | |
| Any heart arrhythmia occurred | categorical | | 258810 | 01:00:00 | True (n=258810) | 10013 | - |
| Has decubitus | categorical | | 8194 | 16:00:00 | True (n=8194) | 14505 | - |
| Shows aphasia or dysarthria | categorical | | 676 | 08:00:00 | True (n=676) | 15408 | - |
| **Feature class: time series high** | | | | | | | |
| Body core temperature | continuous | °C | 3281601 | 00:15:00 | median: 37.30, min: 25.00, max: 44.70 | 400 | 37.3 |
| Diastolic blood pressure | continuous | mmHg | 5028818 | 00:15:00 | median: 57.00, min: 20.00, max: 300.00 | 14 | 57 |
| Estimated respiratory rate | continuous | | 2567702 | 00:15:00 | median: 20.00, min: 6.00, max: 100.00 | 6062 | - |
| FiO2 | continuous | % | 1397347 | 00:15:00 | median: 40.00, min: 0.00, max: 100.00 | 644 | - |
| Heart rate | continuous | bpm | 5679878 | 00:15:00 | median: 83.00, min: 30.00, max: 300.00 | 0 | - |
| Mean blood pressure | continuous | mmHg | 5080902 | 00:15:00 | median: 75.00, min: 20.00, max: 300.00 | 15 | 75 |
| O2 saturation | continuous | % | 5448554 | 00:15:00 | median: 97.00, min: 0.00, max: 101.70 | 1 | 97.0 |
| PAP - PEEP | continuous | mmHg | 802165 | 00:15:00 | median: 13.00, min: -34.00, max: 66.00 | 5336 | - |
| Systolic blood pressure | continuous | mmHg | 5026861 | 00:15:00 | median: 114.00, min: 20.00, max: 300.00 | 14 | 114 |
| **Feature class: time series medium** | | | | | | | |
| Anion gap | continuous | mEq/L | 283297 | 02:40:00 | median: 5.00, min: -97.70, max: 97.60 | 1606 | - |
| BE | continuous | mmol/L | 450028 | 02:43:00 | median: 0.30, min: -24.90, max: 25.00 | 50 | 0.3 |
| Bicarbonate | continuous | mmol/L | 450284 | 02:43:00 | median: 24.50, min: 0.00, max: 74.40 | 50 | 24.5 |
| COHb | continuous | % | 524960 | 02:20:00 | median: 1.40, min: 0.00, max: 7.90 | 61 | 1.4 |
| Calcium | continuous | mmol/L | 453457 | 02:41:00 | median: 1.16, min: 0.00, max: 4.36 | 49 | 1.16 |
| Chloride | continuous | mmol/L | 448707 | 02:43:00 | median: 109.00, min: 0.00, max: 199.00 | 50 | 109.0 |
| Glucose | continuous | mg/dL | 566205 | 02:18:00 | median: 134.00, min: 0.00, max: 1196.00 | 37 | 134.0 |
| Hematocrit | continuous | % | 499803 | 02:33:00 | median: 29.80, min: 0.00, max: 69.40 | 39 | 29.8 |
| Hemoglobin | continuous | mmol/L | 587605 | 02:13:00 | median: 9.60, min: 0.00, max: 28.60 | 40 | 9.6 |
| Lactate | continuous | mmol/L | 533796 | 02:23:00 | median: 1.10, min: 0.00, max: 26.00 | 55 | 1.1 |
| MetHb | continuous | % | 524746 | 02:20:00 | median: 0.90, min: 0.00, max: 15.70 | 61 | 0.9 |
| O2Hb | continuous | % | 275139 | 02:41:00 | median: 95.30, min: 0.00, max: 100.00 | 1616 | - |
| Potassium | continuous | mmol/L | 450215 | 02:49:00 | median: 4.30, min: 0.00, max: 10.00 | 47 | 4.3 |
| RAS scale | continuous | | 1280104 | 01:00:00 | median: 0.00, min: -5.00, max: 5.00 | 4 | 0 |
| RHb | continuous | | 273565 | 02:41:00 | median: 2.10, min: 0.00, max: 87.60 | 1628 | - |
| Sodium | continuous | mmol/L | 448999 | 02:48:00 | median: 138.00, min: 0.00, max: 211.00 | 49 | 138.0 |
| pCO2 | continuous | mmHg | 444251 | 02:44:00 | median: 38.90, min: 0.00, max: 145.90 | 53 | 38.9 |
| pH | continuous | | 435866 | 02:46:00 | median: 7.42, min: 4.77, max: 7.98 | 68 | 7.417 |
| pO2 | continuous | mmHg | 443732 | 02:44:00 | median: 88.80, min: 0.00, max: 719.00 | 54 | 88.8 |
| paO2/FiO2 | continuous | mmHg/FiO2 | 382127 | 02:44:00 | median: 246.00, min: 15.00, max: 2877.62 | 1012 | 246.0 |
| **Feature class: time series low** | | | | | | | |
| Albumin | continuous | g/dL | 5695 | 23:45:00 | median: 2.80, min: 0.00, max: 5.30 | 13676 | - |
| Alkaline phosphatase | continuous | U/L | 10993 | 1 d 00:00:00 | median: 74.00, min: 0.00, max: 499.00 | 11714 | - |
| Antithrombin III | continuous | % | 3064 | 12:30:00 | median: 68.00, min: 30.00, max: 131.00 | 13593 | - |
| Bilirubin total | continuous | mg/dL | 65621 | 23:45:00 | median: 0.60, min: 0.00, max: 49.70 | 2573 | - |
| Blood Urea Nitrogen | continuous | mg/dL | 81272 | 23:45:00 | median: 21.00, min: 0.00, max: 463.00 | 771 | - |
| C-reactive protein | continuous | mg/dL | 76062 | 23:45:00 | median: 6.70, min: 0.00, max: 58.70 | 1224 | - |
| CK | continuous | U/L | 104111 | 23:00:00 | median: 302.00, min: 0.00, max: 10000.00 | 121 | 302.0 |
| CK-MB | continuous | U/L | 75249 | 13:45:00 | median: 28.00, min: 0.00, max: 4116.00 | 1726 | - |
| Cholinesterase | continuous | U/L | 5485 | 23:45:00 | median: 3699.00, min: 0.00, max: 14230.00 | 13405 | - |
| Creatinine | continuous | mg/dL | 94062 | 23:30:00 | median: 0.90, min: 0.00, max: 59.80 | 97 | 0.9 |
| Erythrocytes | continuous | millions/µL | 49093 | 23:45:00 | median: 3.20, min: 0.00, max: 71.00 | 6034 | - |
| Fibrinogene | continuous | mg/dL | 3642 | 10:00:00 | median: 261.00, min: 200.00, max: 350.00 | 13230 | - |
| GCS Eye | continuous | | 49324 | 1 d 00:00:00 | median: 4.00, min: 1.00, max: 4.00 | 2841 | 4 |
| GCS Motor | continuous | | 49325 | 1 d 00:00:00 | median: 6.00, min: 1.00, max: 6.00 | 2841 | 6 |
| GCS Verbal | continuous | | 49325 | 1 d 00:00:00 | median: 5.00, min: 1.00, max: 5.00 | 2841 | 5 |
| GCS score | continuous | | 49321 | 1 d 00:00:00 | median: 15.00, min: 3.00, max: 15.00 | 2841 | 15 |
| GOT (AST) | continuous | U/L | 53538 | 23:45:00 | median: 45.00, min: 0.00, max: 4992.00 | 3171 | - |
| GPT (ALT) | continuous | U/L | 94296 | 23:30:00 | median: 26.00, min: 0.00, max: 4986.00 | 197 | 26.0 |
| Gamma-GT | continuous | U/L | 69286 | 23:45:00 | median: 40.00, min: 0.00, max: 500.00 | 1864 | - |
| LDH | continuous | U/L | 30388 | 23:45:00 | median: 275.00, min: 0.00, max: 1000.00 | 7831 | - |
| Leucocytes | continuous | thousand/µL | 95074 | 23:30:00 | median: 11.49, min: 0.00, max: 99.41 | 117 | 11.49 |
| Lipase | continuous | U/L | 15405 | 1 d 00:00:00 | median: 25.00, min: 0.00, max: 498.00 | 10235 | - |
| MCH | continuous | pg | 32522 | 23:45:00 | median: 29.60, min: 0.00, max: 43.70 | 9424 | - |
| MCHC | continuous | g/dL | 32525 | 23:45:00 | median: 33.20, min: 0.00, max: 100.00 | 9424 | - |
| MCV | continuous | fL | 32527 | 23:45:00 | median: 88.80, min: 0.00, max: 140.50 | 9423 | - |
| Magnesium | continuous | mmol/L | 9155 | 1 d 00:00:00 | median: 0.86, min: 0.14, max: 33.81 | 12296 | - |

| | | | | | | | |
|---|---|---|---|---|---|---|---|
| Maximum decubitus stage | continuous | | 8194 | 16:00:00 | median: 2.00, min: 1.00, max: 4.00 | 14505 | - |
| Osmolality | continuous | mOsm/kg | 6069 | 1 d 00:00:00 | median: 297.00, min: 200.00, max: 350.00 | 13747 | - |
| PTT | continuous | s | 126767 | 13:15:00 | median: 42.00, min: 0.00, max: 236.00 | 84 | 42.0 |
| Phosphate | continuous | mg/dL | 73646 | 23:45:00 | median: 3.30, min: 0.00, max: 12.90 | 1357 | - |
| Procalcitonin | continuous | ng/mL | 28417 | 1 d 00:00:00 | median: 0.40, min: 0.00, max: 5.00 | 8982 | - |
| Protein | continuous | g/dL | 4751 | 1 d 00:00:00 | median: 5.30, min: 0.00, max: 8.70 | 14001 | - |
| Protein (urine) | continuous | mg | 5732 | 1 d 00:26:00 | median: 0.00, min: 0.00, max: 150.00 | 12920 | - |
| Prothrombin time (INR) | continuous | | 107160 | 14:22:00 | median: 1.19, min: 0.00, max: 90.00 | 1180 | - |
| Quick | continuous | % | 117782 | 14:15:00 | median: 75.00, min: 0.00, max: 140.00 | 88 | 75.0 |
| T3 free | continuous | ng/dL | 6352 | 6 d 23:30:00 | median: 1.90, min: 0.40, max: 6.70 | 11762 | - |
| T4 free | continuous | ng/dL | 6661 | 6 d 23:15:00 | median: 1.20, min: 0.20, max: 4.10 | 11612 | - |
| TSH | continuous | µU/mL | 8264 | 6 d 00:30:00 | median: 1.30, min: 0.02, max: 86.00 | 10650 | - |
| Thrombin time | continuous | s | 3318 | 23:45:00 | median: 20.00, min: 15.00, max: 50.00 | 13730 | - |
| Thrombocytes | continuous | thousand/µL | 104086 | 23:00:00 | median: 166.00, min: 0.00, max: 1000.00 | 87 | 166.0 |
| eGFR | continuous | L | 92259 | 23:45:00 | median: 77.61, min: 0.53, max: 306.55 | 108 | 77.60777 |
| **Feature class: flow** | | | | | | | |
| Blood volume in | continuous | mL | 28183 | 01:00:00 | median: 250.00, min: 0.00, max: 3000.00 | 10917 | - |
| Blood volume out | continuous | mL | 1130600 | 00:00:00 | median: 0.00, min: 0.00, max: 5000.00 | 1861 | - |
| Cardiac stimulants (epinephrine equivalence dosage) | continuous | | 292237 | 00:20:00 | median: 11.08, min: 0.00, max: 10556.40 | 11777 | - |
| Drainage volume out | continuous | mL | 34788 | 01:00:00 | median: 7.00, min: 0.00, max: 4000.00 | 14451 | - |
| Enteral nutrition volume in | continuous | mL | 596044 | 01:00:00 | median: 80.00, min: 0.00, max: 2550.00 | 357 | - |
| Feeding tube volume out | continuous | mL | 24857 | 06:55:00 | median: 50.00, min: 1.00, max: 2000.00 | 11390 | - |
| Glucocorticoids (cortison equivalence dosage) | continuous | | 44409 | 01:00:00 | median: 4.00, min: 0.03, max: 5500.00 | 12378 | - |
| Infusion volume in | continuous | mL | 6914612 | 00:00:00 | median: 1.00, min: 0.00, max: 100000.00 | 168 | - |
| Norepinephrine and Dopamine (norepinephrine equivalence dosage) | continuous | | 563212 | 00:20:00 | median: 79.20, min: 0.00, max: 12960.00 | 4792 | - |
| Plasma expander volume in | continuous | mL | 29669 | 01:00:00 | median: 500.00, min: 0.00, max: 1500.00 | 10502 | - |
| Stool volume out | continuous | mL | 71488 | 06:00:00 | median: 100.00, min: 1.00, max: 2000.00 | 8830 | - |
| Ultrafiltrate volume out | continuous | mL | 11942 | 06:00:00 | median: 500.00, min: 0.00, max: 5200.00 | 14984 | - |
| Urine volume out | continuous | mL | 972240 | 01:00:00 | median: 80.00, min: 1.00, max: 5000.00 | 116 | - |
| **Feature class: interventions** | | | | | | | |
| Antithrombotic agents prophylactic dosage | categorical | | 192060 | 01:00:00 | True (n=192060) | 3261 | - |
| Antithrombotic agents therapeutic dosage | categorical | | 869689 | 00:20:00 | True (n=869689) | 7775 | - |
| Arterial line exists | categorical | | 139666 | 08:00:00 | True (n=139666) | 1072 | - |
| Bladder catheter exists | categorical | | 138885 | 08:00:00 | True (n=138885) | 1259 | - |
| Bronchoscopy performed | categorical | | 1020 | 2 d 04:07:30 | True (n=1020) | 15050 | - |
| CCO exists | categorical | | 58782 | 00:15:00 | True (n=58782) | 15341 | - |
| Cardiopulmonary resuscitation performed | categorical | | 153 | 01:50:30 | True (n=153) | 15467 | - |
| Cardioversion performed | categorical | | 642 | 01:55:00 | True (n=642) | 15266 | - |
| Central venous catheter exists | categorical | | 133589 | 08:00:00 | True (n=133589) | 2713 | - |
| Central venous line exists | categorical | | 116510 | 08:00:00 | True (n=116510) | 2922 | - |
| Chest tube exists | categorical | | 431150 | 01:00:00 | True (n=431150) | 3549 | - |
| Defibrillation performed | categorical | | 143 | 00:20:00 | True (n=143) | 15536 | - |
| Dialysis or CVVH performed | categorical | | 11930 | 06:00:00 | True (n=11930) | 14984 | - |
| ECMO or RVAD | categorical | | 31631 | 01:00:00 | True (n=31631) | 15455 | - |
| Feeding system exists | categorical | | 73011 | 05:00:00 | True (n=73011) | 12533 | - |
| IABP | categorical | | 15390 | 01:00:00 | True (n=15390) | 15269 | - |
| ICP probe exists | categorical | | 242 | 08:00:00 | True (n=242) | 15576 | - |
| Is on automatic ventilation | categorical | | 692027 | 00:15:00 | True (n=692027 | 4196 | - |
| LVAD | categorical | | 119238 | 01:00:00 | True (n=18314) | 15191 | - |
| Shaldon catheter exists | categorical | | 18314 | 08:00:00 | True (n=93302) | 14506 | - |
| Thrombosis prophylaxis performed | categorical | | 93302 | 10:00:00 | True (n=93302) | 2018 | - |
| Tracheal cannula exists | categorical | | 17465 | 08:00:00 | True (n=17465) | 15127 | - |
| Tracheal secretion cleaned | categorical | | 80477 | 03:00:00 | True (n=80477) | 7098 | - |
| Tubus exists | categorical | | 24358 | 08:00:00 | True (n=24358) | 6635 | - |
| **Feature class: medication** | | | | | | | |
| ACE inhibitors plain and combinations | categorical | | 21311 | 12:00:00 | Enalapril (C09AA02) (n=16763),Ramipril (C09AA05) (n=3595),Acerbon (C09AA03) (n=393),Lisinopril Tbl. (C09AA03) (n=311),Captopril (C09AA01) (n=227),Delix plus (C09BA25) (n=22) | 11347 | - |
| Anesthestics | categorical | | 261094 | 00:20:00 | Propofol Perf. (N01AX10) (n=246642),Ketamin-S_Perfusor (N01AX03) (n=8088),Propofol (N01AX10) (n=2027),Remifentanil Perf. (N01AH06) (n=1990),Isofluran (N01AB06) (n=1744),Etomidat-Lipuro (N01AX07) (n=303),Ketamin-S (N01AX03) (n=180),Thiopental Perf. (N01AF03) (n=90),Thiopental (N01AF03) (n=22),Hydroxybuttersäure Perf. (N01AX11) (n=8) | 5051 | - |
| Angiotensin II receptor blockers | categorical | | 401 | 22:48:00 | Candesartan (C09CA06) (n=219),Valsartan (C09CA03) (n=92),Lorzaar Tbl. (C09CA01) (n=88),Irbesartan (C09CA04) (n=2) | 15433 | - |
| Antiandrenergic agents, centrally acting | categorical | | 215952 | 00:20:00 | Clonidin Perf. (C02AC01) (n=157259),Dexmedetomidin (C02AC) (n=50275),Catapresan (C02AC01) (n=6792),Moxonidin (C02AC05) (n=1626) | 8255 | - |
| Antiandrenergic agents, peripherally acting | categorical | | 44893 | 01:00:00 | Urapidil Perf. (C02CA06) (n=38927),Urapidil (C02CA06) (n=5966) | 12751 | - |
| Antiarrhythmics, class I and III | categorical | | 100470 | 00:21:00 | Amiodaron (Cordarex) Perf. (C01BD01) (n=86485),Amiodaron (Cordarex) Tbl (C01BD01) (n=9798),Amiodaron (Cordarex) (C01BD01) (n=3319),Ajmalin Perf. (C01BA05) (n=614),Flecainid (C01BC04) (n=174),Ajmalin (C01BA05) (n=50),Propafenon (C01BC03) (n=15),Propafenon Perf. (C01BC03) (n=15) | 13148 | - |
| Antibacterials for systemic use | categorical | | 104097 | 06:00:00 | Cefuroxim (J01DC02) (n=19797),Meropenem (J01DH02) (n=19269),Piperacillin/Tazobactam (J01CR05) (n=13523),Cephazolin (J01DB04) | 4824 | - |

| | | | | | | | |
|---|---|---|---|---|---|---|---|
| | | | | | (n=13097),Vancomycin (J01XA01) (n=6044),Flucloxacillin (J01CF05) (n=4030),Cefotaxim (J01DD01) (n=3036),Clindamycin (J01FF01) (n=2390),Ciprobay (J01MA02) (n=2195),Ampicillin (J01CA01) (n=2006),Ceftazidim (J01DD02) (n=1987),Gentamicin (J01GB03) (n=1651),Linezolid (J01XX08) (n=1451),Metronidazol (Clont) (J01XD01) (n=1337),Fosfomycin (J01XX01) (n=1263),Erythromycin (J01FA01) (n=1240),Penicillin G (J01CE01) (n=1235),Ampicillin/Sulbactam (J01CR01) (n=1001),Ceftriaxon (J01DD04) (n=901),Daptomycin (J01XX09) (n=798),Cotrim (J01EA01) (n=749),Tigecyclin (J01AA12) (n=670),Amoxicillin/Clavulansäure (J01CR02) (n=655),Teicoplanin (J01XA02) (n=471),Colistin (J01XB01) (n=378),Imipenem (J01DH51) (n=363),Moxifloxacin_Tbl (J01MA14) (n=319),Clarithromycin (J01FA09) (n=310),Moxifloxacin (J01MA14) (n=288),Baypen (J01CA10) (n=278),Combactam (J01CG01) (n=245),Metronidazol Tbl. (J01XD01) (n=220),Levofloxacin (J01MA12) (n=170),Ertapenem (J01DH03) (n=159),Tobramycin (J01GB01) (n=138),Amoxicillin (J01CA04) (n=130),Ciprofloxacin (J01MA02) (n=126),Cubicin (J01XX09) (n=117),Biklin (J01GB06) (n=33),Flucloxacillin p.o. (J01CF05) (n=11),Kepinol Tbl. (J01EE01) (n=10),Pipril (J01CA12) (n=4),Binotal (J01CA01) (n=1),Polymyxin B (J01XB02) (n=1) | | |
| Antidepressants | categorical | | 5572 | 1 d 00:00:00 | Escitalopram (N06AB10) (n=2335),Saroten (N06AA09) (n=1341),Cipramil Tbl. (N06AB04) (n=1117),Remergil (N06AX11) (n=779) | 14801 | - |
| Antiemetic preparation | categorical | | 1344 | 1 d 00:00:00 | Folsaeure (B03BB01) (n=478),Ferrosanol (B03AA01) (n=399),Vit.B12 (B03BA53) (n=214),Eisen-III (i.v.) (B03AC01) (n=85),Darbepoetin alfa (B03XA02) (n=80),Eisencarboxymaltose (B03AC01) (n=64),Erythropoetin (B03XA01) (n=20),Eisen-II-sulfat (B03AA07) (n=4) | 15206 | - |
| Antiepileptics | categorical | | 16286 | 01:00:00 | Pregabalin (N03AX16) (n=5290),Levetiracetam (N03AX14) (n=2978),Levetiracetam Perf. (N03AX14) (n=1969),Orfiril Perf. (N03AG01) (n=1804),Neurontin (N03AX12) (n=1578),Phenytoin Perf. (N03AB02) (n=698),Carbamazepin (N03AF01) (n=447),Orfiril (N03AG01) (n=365),Valproinsäure (N03AG01) (n=350),Lacosamid (N03AX18) (n=246),Valproinsäure Perf. (N03AG01) (n=158),Frisium (N03A) (n=123),Clonazepam Perf. (N03AE01) (n=104),Phenytoin (N03AB02) (n=104),Clonazepam (N03AE01) (n=71),Phenobarbital (N03AA02) (n=1) | 14480 | - |
| Antihemorrhagics | categorical | | 5449 | 01:00:00 | Cyclocapron (B02AA02) (n=3429),Konakion (B02BA02) (n=834),Haemocomplettan (B02BB01) (n=463),PPSB (B02BD01) (n=380),Konakion-Tropfen (B02BA02) (n=270),Trasylol (B02AB01) (n=24),Fibrogammin (B02BD07) (n=20),Novo7 (B02BD02) (n=16),Haemate (B02BD06) (n=13) | 14228 | - |
| Antihypertensiva | categorical | | 718002 | 00:20:00 | Clonidin Perf. (C02AC01) (n=157259),Glyceroltrinitrat Perf. (C01DA02) (n=123303),Furosemid Perf. (C03CA01) (n=65459),Dihydralazin (C02DB01) (n=63836),Dexmedetomidin (C02AC) (n=50275),Torasemid (C03CA04) (n=42864),Urapidil Perf. (C02CA06) (n=38927),Beloc-Zok (C07AB02) (n=26663),Nitroprussidnatrium Perf. (C02DD01) (n=26265),Amlodipin (C08CA01) (n=23563),Enalapril (C09AA02) (n=16763),Bisoprolol (C07AB07) (n=16671),Furosemid (C03CA01) (n=13719),Metoprolol (C07AB02) (n=10819),Catapresan (C02AC01) (n=6792),Urapidil (C02CA06) (n=5966),Spironolacton i.v. (C03DA01) (n=4623),Ramipril (C09AA05) (n=3595),Spironolacton Tbl. (C03DA01) (n=3540),ISDN (C01DA08) (n=2345),Querto (C07AG02) (n=2039),Xipamid (C03BA10) (n=1900),Moxonidin (C02AC05) (n=1626),Diltiazem Tbl. (C08DB01) (n=1389),Hydrochlorothiazid (C03AA03) (n=1383),Lonolox (C02DC01) (n=892),Verapamil (C08DA01) (n=594),Nebivolol (C07AB12) (n=579),Acerbon (C09AA03) (n=393),Alprostadil (C04AG01) (n=373),Verapamil Perf. (C08DA01) (n=370),Esmolol Perf. (C07AB09) (n=320),Lisinopril Tbl. (C09AA03) (n=311),Dociton (C07AA05) (n=297),Captopril (C09AA01) (n=227),Eplerenon (C03DA04) (n=224),Candesartan (C09CA06) (n=219),Trental Perf. (C04AD03) (n=167),Nimodipin Saft (C08CA06) (n=163),Lercanidipin (C08CA13) (n=151),Carmen (C08CA13) (n=133),Nimodipin Perf. (C08CA06) (n=126),Molsidomin (Corvaton) (C01DX12) (n=120),Nimodipin Tbl. (C08CA06) (n=101),Diltiazem Perf. (C08DB01) (n=97),Valsartan (C09CA03) (n=92),Lorzaar Tbl. (C09CA01) (n=88),Nitro-Spray (C01DA02) (n=86),Sotalex (C07AA07) (n=69),Molsidomin (C01DX12) (n=66),Diltiazem (C08DB01) (n=62),Nifedipin Tbl. (C08CA05) (n=47),Unat (C03CA04) (n=24),Delix plus (C09BA25) (n=22),Nifedipin Perf. (C08CA05) (n=3),Irbesartan (C09CA04) (n=2) | 2121 | - |
| Antimycotics for systemic use | categorical | | 4313 | 1 d 00:00:00 | Voriconazol Tbl. (J02AC03) (n=1266),Micafungin (J02AX05) (n=1137),Caspofungin (J02AX04) (n=606),Fluconazol (J02AC01) (n=555),Voriconazol (J02AC03) (n=372),Fluconazol Tbl. (J02AC01) (n=129),Posaconazol (J02AC04) (n=88),Anidulafungin (J02AX06) (n=80),AmBisome (J02AA01) (n=61),Flucytosin (J02AX01) (n=19) | 15221 | - |
| Antipsychotics (neuroleptics) | categorical | | 16896 | 07:24:00 | Haldol (N05AD01) (n=9988),Quetiapin (N05AH04) (n=2563),Risperidon (N05AX08) (n=2262),Melperon (N05AD03) (n=1541),Pipamperon-Saft (N05AD05) (n=398),Promethazin (N05A) (n=144) | 13649 | - |
| Antithrombotic agents excl. Platelet inhibitors and enzymes | categorical | | 1044845 | 00:20:00 | Heparin Perf. (B01AB01) (n=847558),Argatroban (B01AE03) (n=162454),Clexane (B01AB05) (n=29516),Heparin (B01AB01) (n=2770),Marcumar (B01AA04) (n=2052),Danaparoid-Na (B01AB09) (n=263),Fondaparinux (B01AX05) (n=220),Rivaroxaban (B01AF01) (n=6),Dabigatran (B01AE07) (n=4),Apixaban (B01AF02) (n=2) | 1572 | - |
| Antivirals for systemic use | categorical | | 4911 | 11:00:00 | Aciclovir(Zovirax) (J05AB01) (n=2305),Cymeven (J05AB06) (n=2117),Valcyte (J05AB14) (n=482),Foscarnet (J05AD01) (n=4),Entecavir (J05AF10) (n=3) | 15337 | - |
| Anxiolytics, hypnotics and sedatives | categorical | | 17372 | 01:00:00 | Midazolam Perf. (N05CD08) (n=8913),Lorazepam (N05BA06) (n=4924),Midazolam (N05CD08) (n=1530),Zolpidem (N05CF02) (n=825),Zopiclon (N05CF01) (n=408),Tranxilium (N05BA05) (n=304),Diazepam (N05BA01) (n=243),Rohypnol (N05CD03) (n=154),Bromazepam (N05BA08) (n=44),Temazepam (N05CD07) (n=25),Flurazepam (N05CD01) (n=1),Clomethiazol (N05CM02) (n=1) | 12600 | - |
| Arteriolar smooth muscle, agents acting on | categorical | | 90993 | 00:20:00 | Dihydralazin (C02DB01) (n=63836),Nitroprussidnatrium Perf. (C02DD01) (n=26265),Lonolox (C02DC01) (n=892) | 14015 | - |
| Beta blocking agents | categorical | | 57457 | 12:00:00 | Beloc-Zok (C07AB02) (n=26663),Bisoprolol (C07AB07) (n=16671),Metoprolol (C07AB02) (n=10819),Querto (C07AG02) (n=2039),Nebivolol (C07AB12) (n=579),Esmolol Perf. (C07AB09) (n=320),Dociton (C07AA05) (n=297),Sotalex (C07AA07) (n=69) | 5931 | - |
| Cardiac stimulants | categorical | | 312110 | 00:20:00 | Epinephrin Perf. (C01CA24) (n=145660),Dobutamin Perf. (C01CA07) (n=133546),Milrinon Perf. (C01CE01) (n=20282),Levosimendan (C01CX08) (n=12068),Epinephrin (C01CA24) (n=554) | 11776 | - |
| Digitalis glycosides | categorical | | 3757 | 1 d 00:00:00 | Digimerck (C01AA04) (n=1891),Lanicor (C01AA05) (n=1746),Lanitop (C01AA08) (n=78),Novodigal (C01AA02) (n=42) | 14670 | - |

| | | | | | | | |
|---|---|---|---|---|---|---|---|
| Diuretics | categorical | | 133736 | 01:00:00 | Furosemid Perf. (C03CA01) (n=65459),Torasemid (C03CA04) (n=42864),Furosemid (C03CA01) (n=13719),Spironolacton i.v. (C03DA01) (n=4623),Spironolacton Tbl. (C03DA01) (n=3540),Xipamid (C03BA10) (n=1900),Hydrochlorothiazid (C03AA03) (n=1383),Eplerenon (C03DA04) (n=224),Unat (C03CA04) (n=24) | 7785 | - |
| Drugs for constipation | categorical | | 110436 | 08:00:00 | Movicol (A06AD15) (n=53310),Laxoberal (A06AB08) (n=47723),Dulcolax (A06AB02) (n=7481),Practo-Klyss (A06AG20) (n=939),Bifiteral (A06AD11) (n=868),Prucaloprid (A06AX05) (n=88),Methylnaltrexon (A06AH01) (n=27) | 3780 | - |
| Drugs for functional gastrointestinal disorders | categorical | | 8482 | 08:00:00 | MCP (Gastrosil) (A03FA01) (n=3599),Sab simplex (A03AX13) (n=2331),Neostigmin (A03) (n=1961),Buscopan (A03BB01) (n=230),Lefax (A03AX13) (n=186),Domperidon (A03FA03) (n=168),Robinul (A03AB02) (n=7) | 13845 | - |
| Drugs for obstructive airway diseases | categorical | | 35537 | 06:00:00 | Sultanol (R03AC02) (n=10411),Berodual (R03AL01) (n=7918),Sultanol_Atrovent (R03AL02) (n=5187),Pulmicort DA (R03BA02) (n=2577),Formoterol (R03AC13) (n=1662),Reproterol Perf. (R03CC14) (n=1471),Budesonid (R03BA02) (n=1453),Orciprenalin Perf. (R03CB03) (n=1418),Berotec DA (R03AC04) (n=845),Tiotropiumbromid (R03BB04) (n=676),Bricanyl (R03CC03) (n=608),Theophyllin Perf. (R03DA04) (n=514),Symbicort (R03AK07) (n=349),Reproterol (R03CC14) (n=244),Theophyllin (R03DA04) (n=152),Flutide N forte DA (R03BA05) (n=41),Spiropent (R03CC13) (n=7),Orciprenalin (R03CB03) (n=4) | 13419 | - |
| Drugs for peptic ulcer and reflux without PPIs | categorical | | 13218 | 08:00:00 | Ranitidin (Zantic) (A02BA02) (n=11974),Ulcogant (A02BX02) (n=1214),Gastrozepin (A02BX03) (n=24),Cimetidin (A02BA01) (n=6) | 11554 | - |
| Drugs used in diabetes | categorical | | 996126 | 00:20:00 | Insulin Perf. (A10AB01) (n=988253),Insulin (A10AB01) (n=4988),Metformin (A10BA02) (n=2051),Insuman (A10AB01) (n=395),Sitagliptin (A10BH01) (n=198),Euglucon (A10BB01) (n=134),Insulin glargin (A10AE04) (n=78),Novorapid (A10AB05) (n=29) | 5420 | - |
| Glucocorticoids | categorical | | 44821 | 01:00:00 | Hydrocortison Perf. (H02AB09) (n=27905),Solu-Decortin H (H02AB06) (n=5470),Decortin (H02AB07) (n=4897),Prednisolon (H02AB06) (n=3145),Dexamethason (H02AB02) (n=1360),Methylprednisolon (H02AB04) (n=1323),Hydrocortison (H02AB09) (n=721) | 12376 | - |
| Immunglobulins | categorical | | 270 | 22:00:00 | Cytotect (J06BB09) (n=233),Pentaglobin (J06BA02) (n=16),Privigen (J06BA02) (n=14),Tetagam (J06BB02) (n=6),Varitect (J06BB03) (n=1) | 15467 | - |
| Immunsuppressants | categorical | | 79008 | 01:00:00 | Ciclosporintabletten (L04AD01) (n=54402),Prograf (L04AD02) (n=8785),Cellcept (L04AA06) (n=7023),Sandimmun Optoral (L04AD01) (n=3778),Ciclosporin (L04AD01) (n=3608),Azathioprin (L04AX01) (n=557),ATG (L04AA18) (n=475),Everolimus (L04AA18) (n=260),Rapamune (L04AA10) (n=119),Alemtuzumab (L04AA34) (n=1) | 15169 | - |
| Inhalative vasodilatators | categorical | | 72185 | 01:59:00 | Sildenafil (C02KX06) (n=27837),Ilomedin (C02KX08) (n=27741),NO (C02KX) (n=13711),Milrinon (C02KX) (n=1905),Bosentan (C02KX01) (n=957),Ilomedin-Perf. (C02KX08) (n=34) | 14512 | - |
| Lipid modifying agents | categorical | | 24342 | 1 d 00:00:00 | Zocor (C10AA01) (n=8390),Atorvastatin (C10AA05) (n=6255),Simvastatin (C10AA01) (n=5787),Pravastatin (C10AA03) (n=3628),Ezetimib (C10AX09) (n=282) | 8766 | - |
| Mineral supplements | categorical | | 692876 | 00:20:00 | Kalium Perf. (A12BA01) (n=617315),Calciumchlorid Perf. (A12AA07) (n=23516),Kalinor-Brause (A12BA02) (n=18355),NaCl 20proz. (A12CA01) (n=6925),Natriumphosphat (A12) (n=5486),Magnesiumaspartat (A12CC05) (n=5235),Magnesium p.o. (A12CC30) (n=4231),Calciumgluconat Perf. (A12AA03) (n=4184),Kalinor-Kps. (A12BA02) (n=2471),Magnesiumsulfat Perf. (A12CC02) (n=2328),Calciumgluconat (A12AA03) (n=744),Kalium (A12BA01) (n=678),Magnesiumaspartat Perf. (A12CC05) (n=404),Selenase (A12CE02) (n=400),Ideos (A12AX01) (n=260),Phosphat_Brausetbl (A12CX50) (n=183),Phosphat_Filmtbl (A12CX50) (n=89),NaCl 3proz (A12CA01) (n=49),NaCl Tablette (A12) (n=23) | 5286 | - |
| Minirin | categorical | | 275 | 12:00:00 | Minirin (H01BA02) (n=275) | 15375 | - |
| Muscle relexants | categorical | | 2562 | 06:00:00 | Cisatracurium (M03AC11) (n=1507),Esmeron (M03AC09) (n=531),Cisatracurium Perf. (M03AC11) (n=419),Baclofen (M03BX01) (n=71),Dantrolen (M03CA01) (n=26),Succinylcholin (M03AB01) (n=8) | 14418 | - |
| Non-opioid analgetics | categorical | | 59127 | 06:00:00 | Novaminsulfon-Tropfen (N02BB02) (n=18636),Novaminsulfon (Novalgin) (N02BB02) (n=14675),Perfalgan (N02BE01) (n=11694),Benuron (N02BE01) (n=8739),Paracetamol (N02BE01) (n=3337),Arcoxia (M01AH05) (n=795),Ibuprofen (M01AE01) (n=662),Voltaren (M01AB05) (n=326),Indometacin (M01AB01) (n=260),Prodafalgan (N02BE01) (n=3) | 5659 | - |
| Norepinephrine and Dopamine | categorical | | 581798 | 00:20:00 | Norepinephrin Perf. (C01CA03) (n=581700),Dopamin Perf. (C01CA04) (n=65),Norepinephrin (C01CA03) (n=33) | 4788 | - |
| Opioids | categorical | | 265173 | 01:00:00 | Piritramid (N02AC03) (n=107223),Sufentanil Perf. (N02AB07) (n=75601),Piritramid Perf. (N02AC03) (n=26651),Palladon (N02AA03) (n=14227),Palladon retard (N02AA03) (n=13765),Hydromorphon (N02AA03) (n=12176),Morphin (N02AA01) (n=8476),Targin (N02AA05) (n=4938),Oxygesic (N02AA05) (n=988),Sufentanil (N02AB07) (n=400),Durogesic-Pflaster (N02AB03) (n=280),Fentanyl (N02AB03) (n=148),Fentanyl Perf. (N02AB03) (n=65),Levo-Methadon (N02AC52) (n=57),Fentanyl TTS (N02AB03) (n=51),MST_Granulat (N02AA01) (n=48),Pethidin (N02AB02) (n=39),Buprenorphin (N02AE01) (n=30),Tramadol (N02AX02) (n=10) | 1754 | - |
| Platelet aggregation inhibitors excl. Heparin | categorical | | 38581 | 1 d 00:00:00 | ASS (B01AC06) (n=30526),Clopidogrel (B01AC04) (n=7156),Ticagrelor (B01AC24) (n=768),Prasugrel (B01AC22) (n=105),Aggrastat (B01AC17) (n=22),Aggrenox (B01AC36) (n=4) | 6609 | - |
| Proton pump inhibitor | categorical | | 49007 | 1 d 00:00:00 | Pantoprazol (A02BC02) (n=26597),Nexium (A02BC05) (n=13581),Esomeprazol (A02BC05) (n=8559),Omeprazol (A02BC01) (n=270) | 3204 | - |
| Selective calcium channel blockers with direct cardiac effects | categorical | | 2512 | 01:00:00 | Diltiazem Tbl. (C08DB01) (n=1389),Verapamil (C08DA01) (n=594),Verapamil Perf. (C08DA01) (n=370),Diltiazem Perf. (C08DB01) (n=97),Diltiazem (C08DB01) (n=62) | 15357 | - |
| Selective calcium channel blockers with mainly vascular effects | categorical | | 24287 | 04:00:00 | Amlodipin (C08CA01) (n=23563),Nimodipin Saft (C08CA06) (n=163),Lercanidipin (C08CA13) (n=151),Carmen (C08CA13) (n=133),Nimodipin Perf. (C08CA06) (n=126),Nimodipin Tbl. (C08CA06) (n=101),Nifedipin Tbl. (C08CA05) (n=47),Nifedipin Perf. (C08CA05) (n=3) | 11232 | - |
| Vasodilatators used in cardiac diseases | categorical | | 125920 | 00:20:00 | Glyceroltrinitrat Perf. (C01DA02) (n=123303),ISDN (C01DA08) (n=2345),Molsidomin (Corvaton) (C01DX12) (n=120),Nitro-Spray (C01DA02) (n=86),Molsidomin (C01DX12) (n=66) | 12798 | - |
| Vasopressin and analogues | categorical | | 22053 | 00:20:00 | Vasopressin Perf. (H01BA01) (n=19004),Vasopressin (Pitresin) (H01BA01) (n=2984),Glycylpressin (H01BA04) (n=65) | 15227 | - |

### Features

*Table 3. **Overview of Feature class.** Every variable was assigned to one feature class and the according features were generated. Timeseries variables were assigned to classes based on their median sampling frequency. For each time series variable, five statistical quantities were generated for three time intervals. Flow and medication Feature class use the same intervals as time series low. Medication features only use an indicator if a substance was given and the number of different substances because dosage information was of insufficient quality.*

| Feature class | Description | Generated features (total number) |
|---|---|---|
| static per patient | Variables are usually collected once per patient (e.g. height). | Last value (n=1) |
| static per hospital stay | Variables are usually collected once per hospital stay (e.g. admission type) | Last value per hospital stay (n=1) |
| static per ICU stay | Variables relevant for a single ICU stay (e.g. did heart arrhythmia occur) | Last value per ICU stay (n=1) |
| time series high | Variables are collected repeatedly with a median sampling interval <= 15 min (e.g. heart frequency). | For 4h, 12h, 24h intervals before ICU: median, IQR, min, max, trend (n=15) |
| time series medium | Timeseries with a median sampling interval <= 6 h (e.g. hemoglobin lab value). | For 12h, 24h, 3d intervals before ICU discharge: median, IQR, min, max, trend (n=15) |
| time series low | Timeseries with a median sampling interval > 6 h (e.g. GCS score). | For 1d, 3d, 7d intervals before ICU discharge: median, IQR, min, max, trend (n=15) |
| flow | Variables containing a continuous flow (e.g. urine) | For 1d, 3d, 7d intervals before ICU discharge: extrapolation of daily input/output (n=3) |
| medication | Medication variables were assigned to WHO ATC codes and grouped according to usage. | For 1d, 3d, 7d intervals before ICU: indicator if at least one substance of a group received and number of different substances from a group (n=6) |
| intervention | Indicator for intervention performed during the ICU stay. | A boolean indicator if an intervention was performed and the time interval between discharge and last time it was performed (n=2) |

*Table 4. **Features generated in addition to the standard feature classes.** There were no variables in the PDMS for these features, so they were generated based on administrative data in the HIS.*

| Variable name | Feature |
|---|---|
| HIS data | - An indicator if current stay a 3d-ICU readmission<br>- Length of ICU stay<br>- Length of hospital stay before ICU admission<br>- Name of ICU station |

### Parameter Tuning

#### *Explainable Boosting Machine*

Based on implementation in interpret.glassbox.ExplainableBoostingClassifier in interpret library with slight modifications enabling unknown values and exposing the argument min_samples_bin.

*Table 5. Parameter settings for parameter tuning of EBM.*

| Parameter | Values | Comment |
|---|---|---|
| learning_rate | $10^x$ for x from -12 to 1 | |
| min_samples_leaf | 2 | Not tuned because determined by min_samples_bin. |
| max_leaves | 2, 4, 6, 8, 10 | |
| interactions | 0 | |
| min_samples_bin | 100, 150, 200, 250, 300, 350, 400 | |
| binning | quantile | |
| max_rounds | 5000 | |
| outer_bags | 8 | The default value, which was a good trade-off between robustness and computation time. |
| random_state | 87 | Project-wide seed determined a-priori. |
| max_interaction_bins | 4, 6, 8, 10, 12, 14 | Considered 2D training for at most 20 iterations. |

Best parameters for all features:

learning_rate: 0.01, max_leaves: 8, min_samples_bin: 200

Best parameters for EBM with limited size (80 1D risk function, 5 2D risk functions):

learning_rate: 0.1, max_leaves: 4, min_samples_bin: 200, max_interaction_bins: 4

#### *Logistic Regression*

Based on implementation in sklearn.linear_model.LogisticRegression in scikit-learn library.

*Table 6. Parameter settings for parameter tuning of LR model.*

| Parameter | Values | Comment |
|---|---|---|
| C | $2^x$ for x from -20 to 3 | Taken from https://www.csie.ntu.edu.tw/~cjlin/papers/liblinear.pdf and refined by additional experiments. |
| penalty | l1, l2 | |
| tol | 0.1, 0.01, 0.001, 0.0001, 0.00001 | |
| solver | liblinear | |
| max_iter | 10, 100, 500 | |
| random_state | 87 | Project-wide seed determined a-priori. |

Best parameters for all features:

C: 0.125, penalty: ‘l1’, tol: 0.01, max_iter: 100 (same results for 500)

Best parameters for LR model with limited size (130 features):

C: 1, penalty: ‘l1’, tol: 0.1, max_iter: 100 (same results for 10, 500)

#### *Gradient Boosting Machine (XGBoost)*

Based on implementation in xgboost.sklearn.XGBModel in xgboost library.

*Table 7. Parameter settings for parameter tuning of GBM model.*

| Parameter | Values | Comment |
|---|---|---|
| learning_rate | x*0.02 for x from 1 to 10 | |
| min_child_weight | 1, 3, 5, 7, 9 | |
| max_depth | 1, 2, 3, 4, 5 | Already observed overfitting for 4-5 so used in as maximum. |
| objective | binary:logistic | |
| n_estimators | 100, 500 | |
| eval_metric | aucpr | |
| use_label_encoder | False | Hide deprecation warning. |
| seed | 87 | Project-wide seed determined a-priori. |

Best parameters for all features:

learning_rate: 0.02, min_child_weight: 7, max_depth: 3, n_estimators: 500

#### *Recurrent Neural Network with Long Short-Term Memory*

Based on implementation in tensorflow.keras.Model in tensorflow library.

*Table 8. Parameter settings for parameter tuning of RNN model.*

| Parameter | Values | Comment |
|---|---|---|
| lstm_neurons | 32, 64 | RNN layers for time series data. |
| dense_neurons | 64, 128 | Fully connected layers for RNN output and static variables. |
| dropout | 0, 0.1, 0.2, 0.3, 0.4, 0.5 | |
| recurrent_dropout | 0 | |
| learning_rate | 0.1, 0.01, 0.001, 0.0001, 0.00001 | |
| batch_size | 32 | |
| rnn_layers | 1 | The number of RNN layers. Preliminary experiments with more layers showed no benefit. |
| epochs | 1, 2, 3, 4, 5, 6, 7, 8, 9, 10 | |
| seed | 87 | Project-wide seed determined a-priori. |

Best parameters for all features:

lstm_neurons: 64, dense_neurons: 64, dropout: 0.4, learning_rate: 0.001, epochs: 3

### Risk function selection for EBM and feature selection for LR models

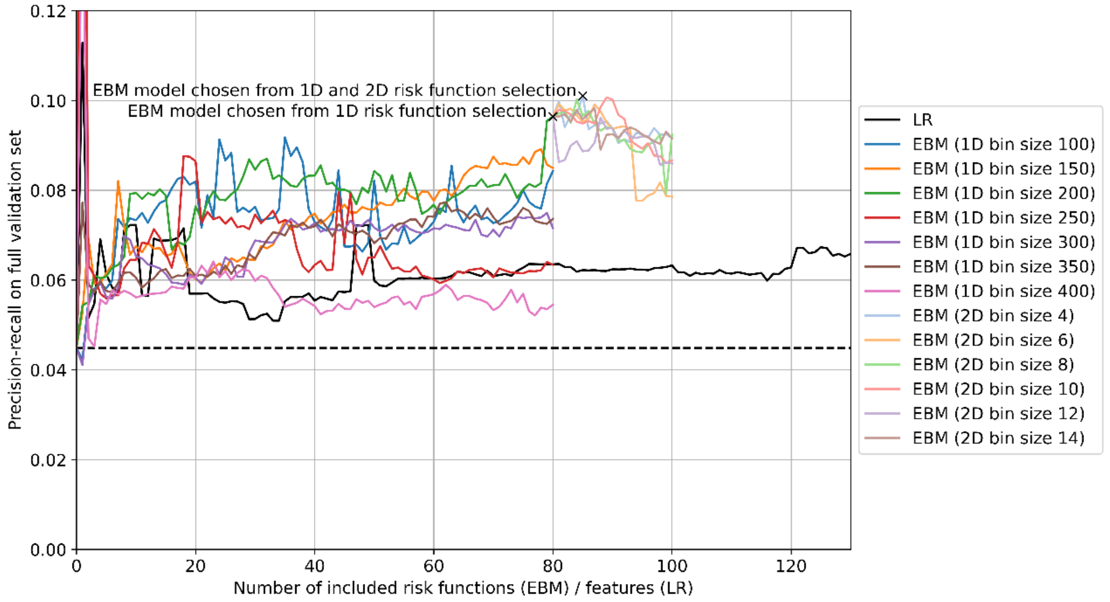


*Figure 3. **PR-AUC on full validation split during risk function (EBM) and feature (LR) selection**. For each step during the selection procedure, we evaluated the PR-AUC on the validation data of the full split. For EBMs we tried different bin sizes. The results are noisy. The best validation score is reached for a bin size of 200 and 80 risk functions. We repeated this procedure for 2D risk functions with a different bin size for 2D risk functions. The best result was achieved with a bin size of four and five risk functions. We selected 130 features for logistic regression since adding dummy variables for categorical features and unknown indicators increased the input from 1,423 to 2,346. So, the logistic regression also incorporates the same ratio of the overall input data.*

### Risk functions of final EBM model

**1.** Age (static all data) [years] x BE (iqr 3d) [mmol/L]

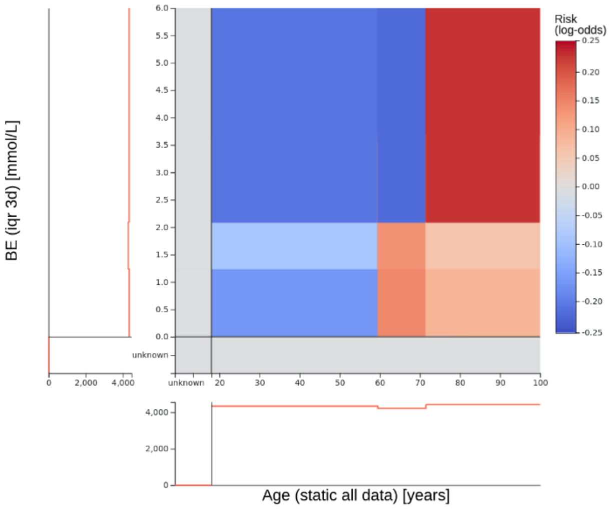


Relative importance: 4.20%

Applicable exclusion criteria: 4

Notes: -

Decision: 3

**2.** Drugs for constipation (unique 1d) x Leucocytes (median 1d) [thousand/µL]

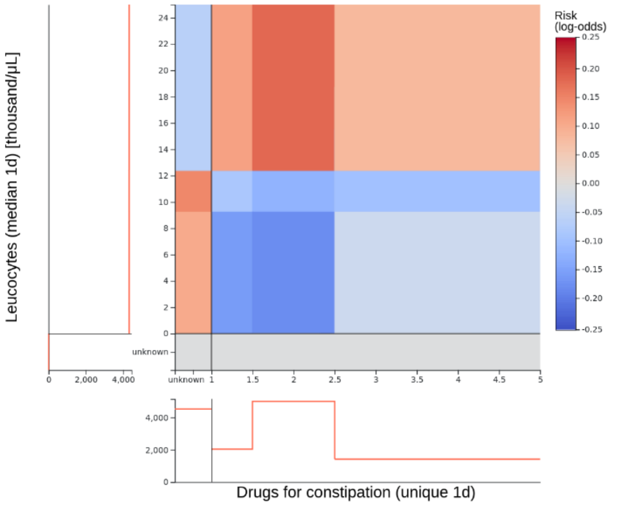


Relative importance: 3.52%

Applicable exclusion criteria: 4

Notes: -

Decision: 3

**3.** Blood volume out (extrapolate 7d) [mL] x Procalcitonin (max 7d) [ng/mL]

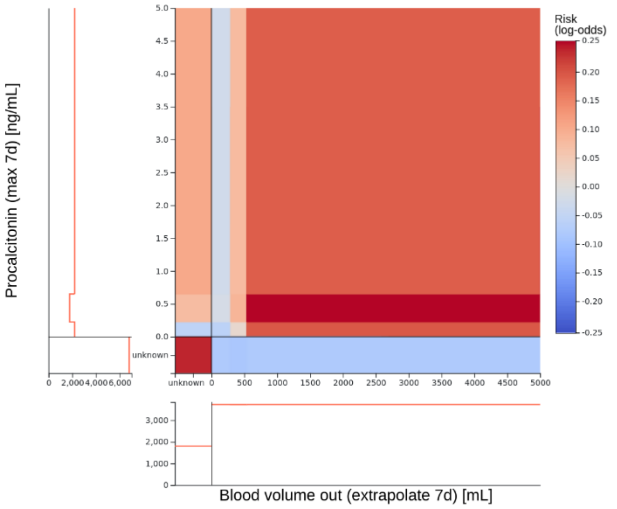


Relative importance: 2.57%

Applicable exclusion criteria: 4

Notes: -

Decision: 3

**4.** Hematocrit (max 3d) [%] x Blood volume out (extrapolate 3d) [mL]

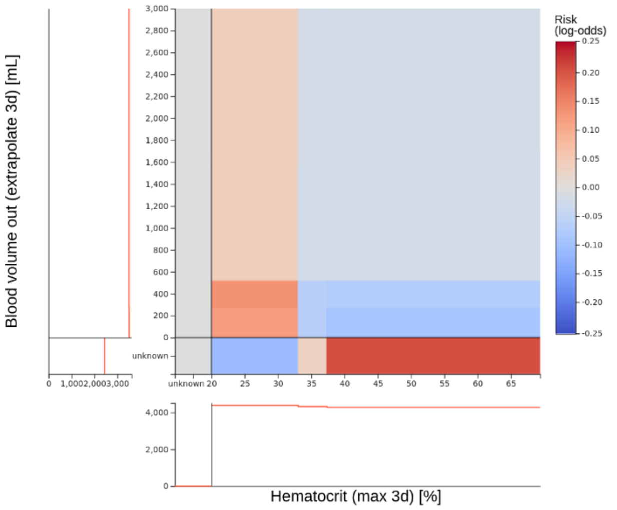


Relative importance: 2.19%

Applicable exclusion criteria: 4

Notes: -

Decision: 3

**5.** Leucocytes (median 1d) [thousand/µL] x Blood volume out (extrapolate 3d) [mL]

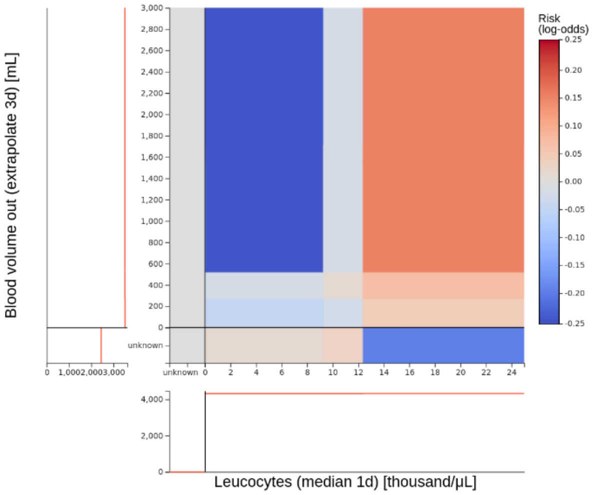


Relative importance: 1.87%

Applicable exclusion criteria: 4

Notes: -

Decision: 3

**6.** Tubus exists (days since last application per icu stay)

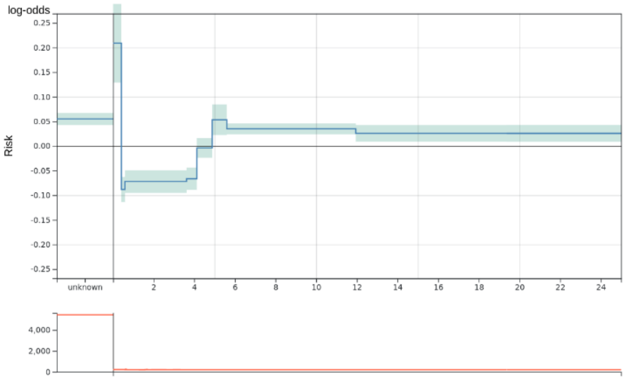


Tubus exists (days since last application per icu stay)

Relative importance: 1.71%

Applicable exclusion criteria: -

Notes:

- Decreased risk between 0.416 and 4.130 might be due to surgical patients that are extubated as planned

Decision: 1

**7.** Age (static all data) [years]

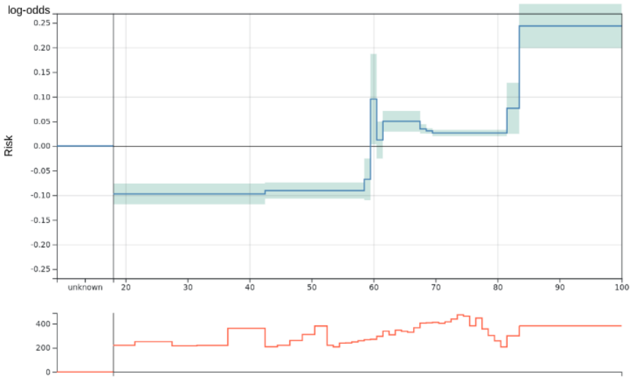


Age (static all data) [years]

Relative importance: 1.70%

Applicable exclusion criteria: 3

Notes:

- Effect of peak at 60 considered as negligible

Decision: 2

**8.** Antithrombotic agents prophylactic dosage (days since last appl. per icu stay)

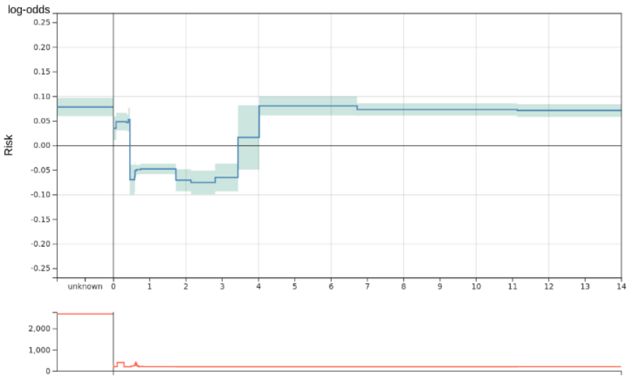


Antithrombotic agents prophylactic dosage (days since last application per icu stay)

Relative importance: 1.65%

Applicable exclusion criteria: -

Notes:

- Difficult to determine patient cohorts responsible for different interval
- Information about therapeutic dosage would be helpful, but not included in the model

Decision: 1

**9.** PTT (max 1d) [s]

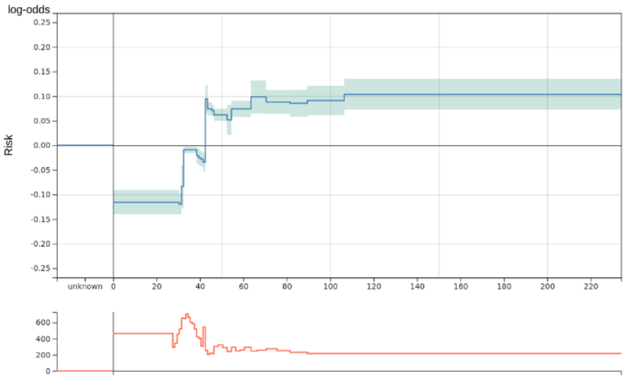


PTT (max 1d) [s]

Relative importance: 1.63%

Applicable exclusion criteria: 2

Notes:

- Practice of measuring PTT changed in 2019, which cannot be corrected easily.

Decision: 3

**10.** O2 saturation (min 12h) [%]

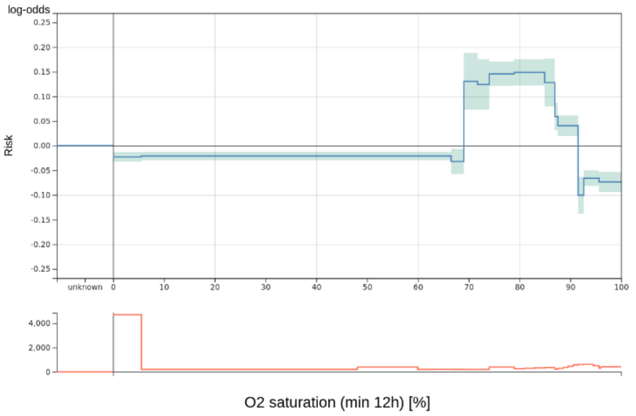


Relative importance: 1.58%

Applicable exclusion criteria: -

Notes: -

Decision: 1

**11.** Blood volume out (extrapolate 7d) [mL]

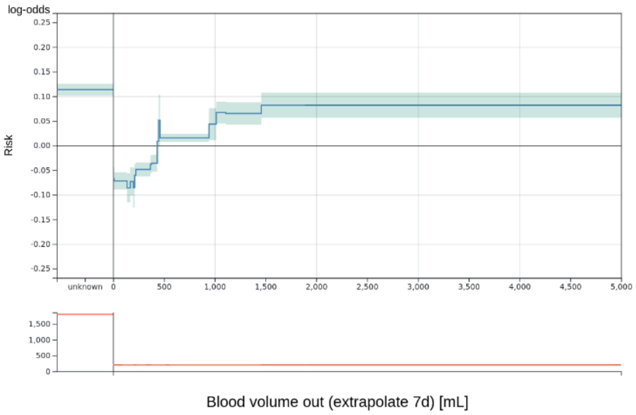


Relative importance: 1.52%

Applicable exclusion criteria: -

Notes:

- Effect of peak at 450 ml considered as negligible

Decision: 1

**12.** Gamma-GT (median 7d) [U/L]

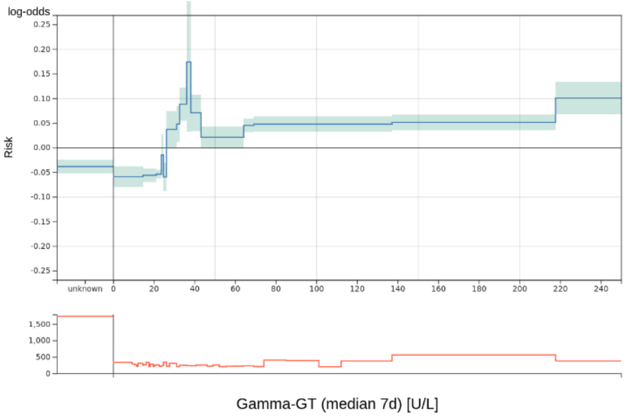


Relative importance: 1.46%

Applicable exclusion criteria: -

Notes:

- Higher risk between 26.25 and 43.25 probably due to medical patients (i.e. no surgery)

Decision: 1

**13.** Chloride (trend per day 3d) [mmol/L]

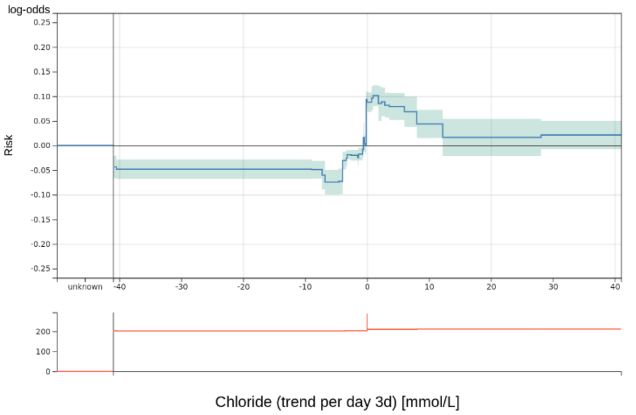


Relative importance: 1.40%

Applicable exclusion criteria: -

Notes:

- Mixed cohort of hyper- and hypochloremia making it hard to determine a general trend
- More hyperchloremia patients, so that negative trend better

Decision: 1

**14.** Heart rate (min 4h) [bpm]

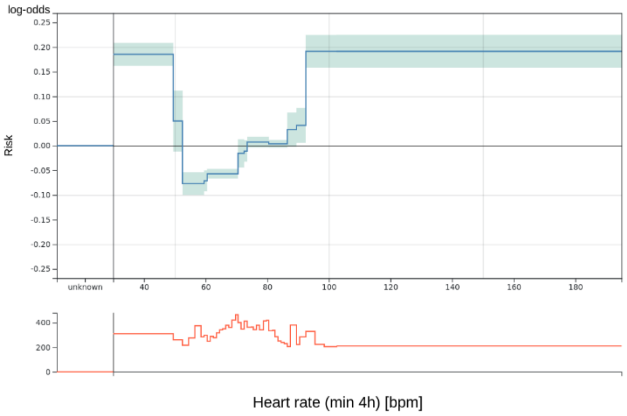


Relative importance: 1.39%

Applicable exclusion criteria: -

Notes: -

Decision: 1

**15.** PTT (max 3d) [s]

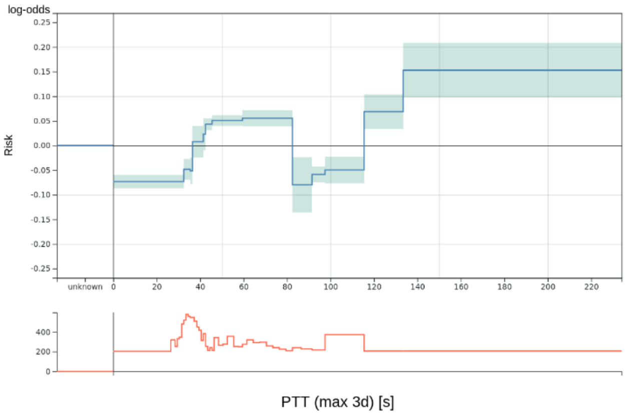


Relative importance: 1.37%

Applicable exclusion criteria: 2

Notes:

- Practice of measuring PTT changed in 2019, which cannot be corrected easily

Decision: 3

**16.** Chloride (min 1d) [mmol/L]

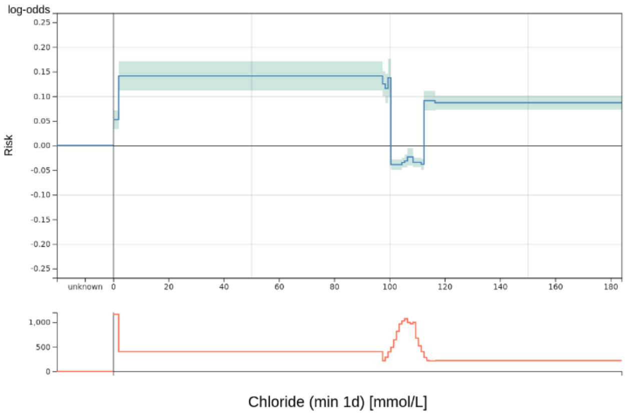


Relative importance: 1.37%

Applicable exclusion criteria: -

Notes: -

Decision: 1

**17.** Hemoglobin (max 3d) [mmol/L]

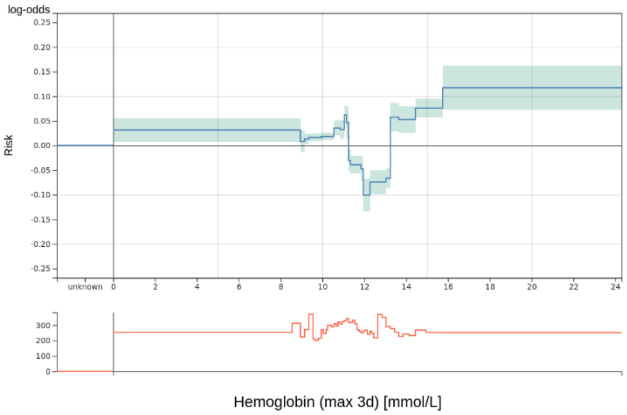


Relative importance: 1.30%

Applicable exclusion criteria: -

Notes: -

Decision: 1

**18.** Length of stay before ICU [days]

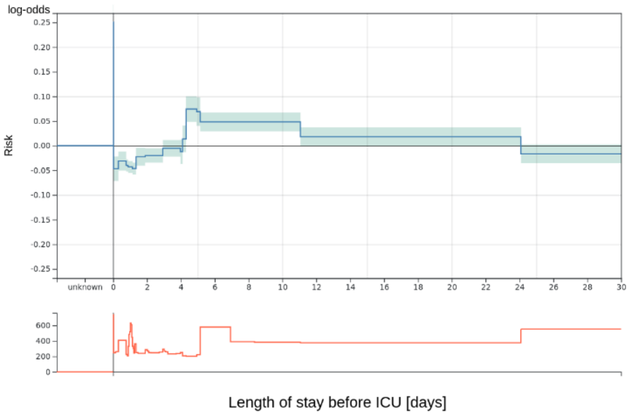


Relative importance: 1.28%

Applicable exclusion criteria: -

Notes:

- Difficult to interpret for patients with repeated ICU stays where this quantity is high.

Decision: 1

**19.** Hematocrit (max 3d) [%]

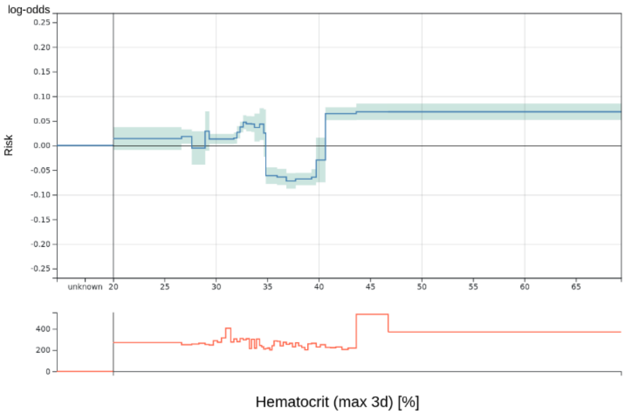


Relative importance: 1.26%

Applicable exclusion criteria: -

Notes: -

Decision: 1

**20.** Calcium (trend per day 3d) [mmol/L]

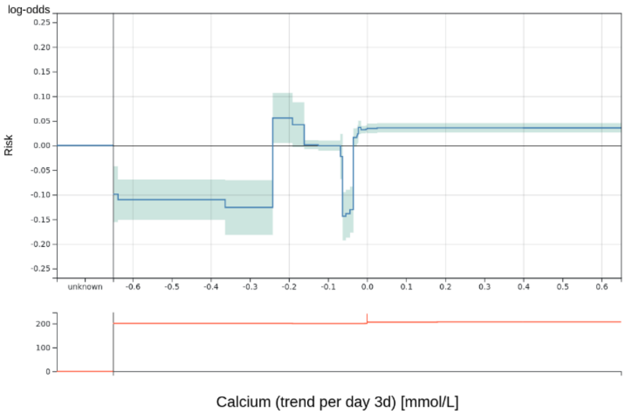


Relative importance: 1.26%

Applicable exclusion criteria: 3, 4

Notes: -

Decision: 3

**21.** eGFR (trend per day 7d) [L]

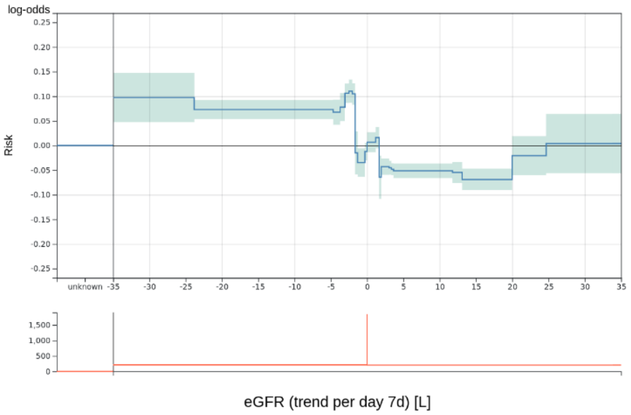


Relative importance: 1.24%

Applicable exclusion criteria: -

Notes: -

Decision: 1

**22.** RAS scale (max 3d)

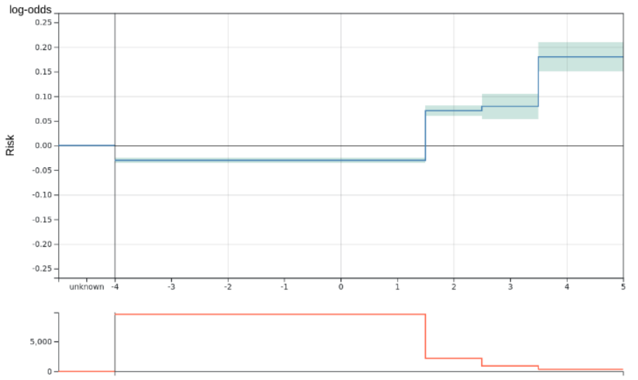


RAS scale (max 3d)

Relative importance: 1.24%

Applicable exclusion criteria: -

Notes: -

Decision: 1

**23.** Urine volume out (extrapolate 1d) [mL]

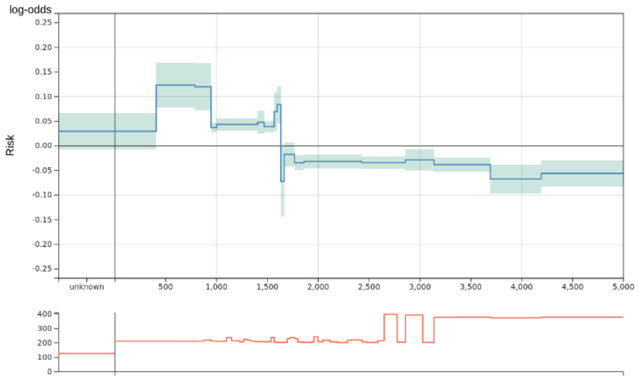


Urine volume out (extrapolate 1d) [mL]

Relative importance: 1.24%

Applicable exclusion criteria: -

Notes:

- Peaks at 1600 and 1650 considered as irrelevant

Decision: 1

**24.** Thrombocytes (trend per day 7d) [thousand/μL]

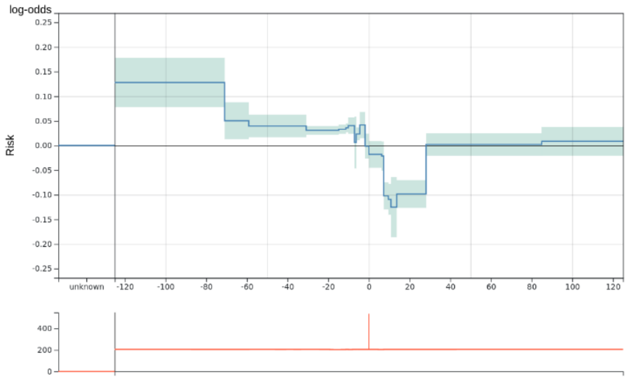


Thrombocytes (trend per day 7d) [thousand/μL]

Relative importance: 1.24%

Applicable exclusion criteria: -

Notes: -

Decision: 1

**25.** Blood volume out (extrapolate 3d) [mL]

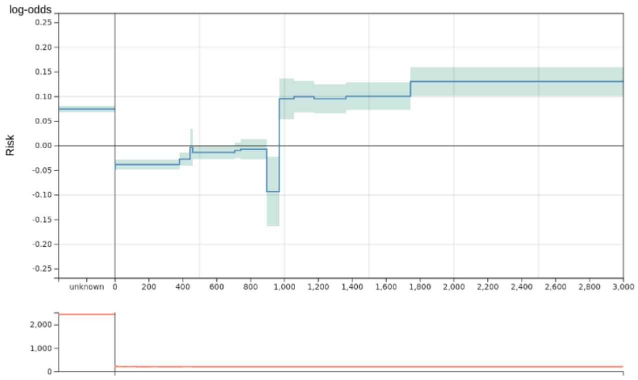


Blood volume out (extrapolate 3d) [mL]

Relative importance: 1.23%

Applicable exclusion criteria: 3

Notes:

- No medical explanation for drop at 900.

Decision: 2

**26.** paO2/FiO2 (median 1d) [mmHg/FiO2]

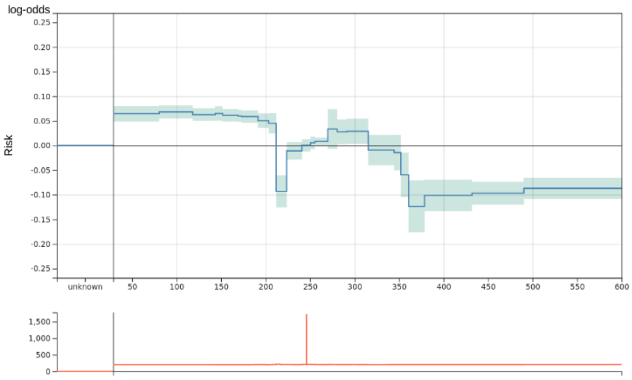


paO2/FiO2 (median 1d) [mmHg/FiO2]

Relative importance: 1.21%

Applicable exclusion criteria: 2

Notes:

- Drop at 215 because of venous blood gas analyses.

Decision: 2

**27.** pH (trend per day 3d)

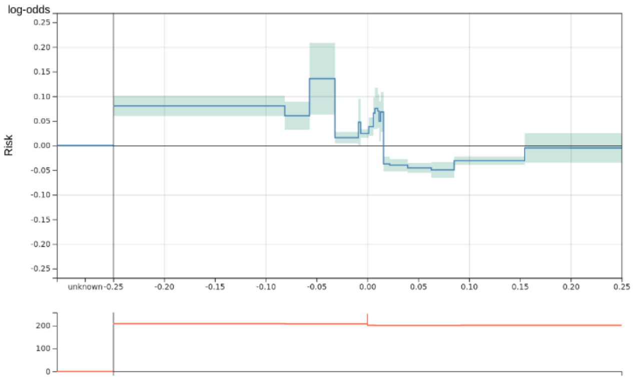


pH (trend per day 3d)

Relative importance: 1.21%

Applicable exclusion criteria: -

Notes: -

Decision: 1

**28.** Phosphate (min 7d) [mg/dL]

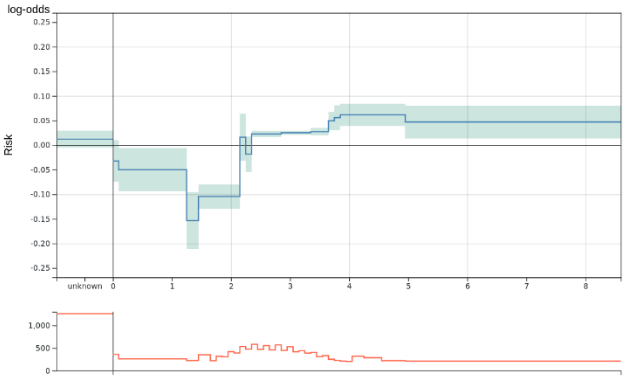


Phosphate (min 7d) [mg/dL]

Relative importance: 1.20%

Applicable exclusion criteria: 3

Notes:

- Increasing risk for values larger than 2.35 might be due to cohort of ICU discharges.

Decision: 2

**29.** pH (median 1d)

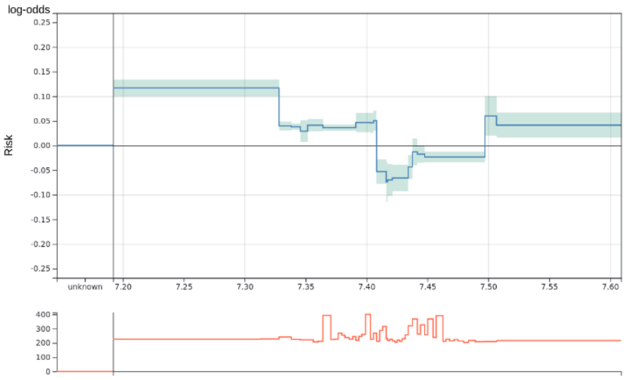


pH (median 1d)

Relative importance: 1.20%

Applicable exclusion criteria: -

Notes: -

Decision: 1

**30.** Body core temperature (min 1d) [°C]

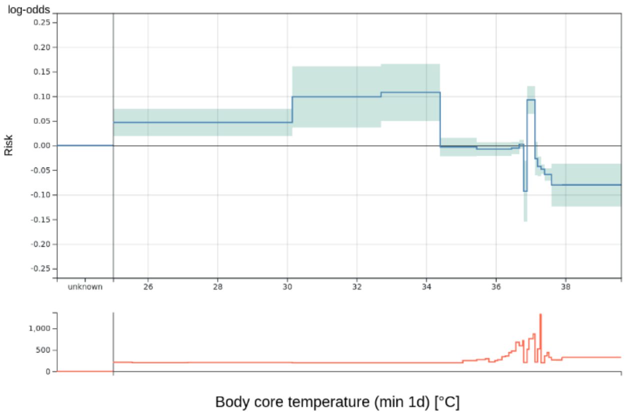


Relative importance: 1.18%

Applicable exclusion criteria: 3

Notes:

- Strong fluctuation for similar value ranges.
- Maximum bin goes from 37.9 to 39.6 which should behave differently

Decision: 3

**31.** CK (min 7d) [U/L]

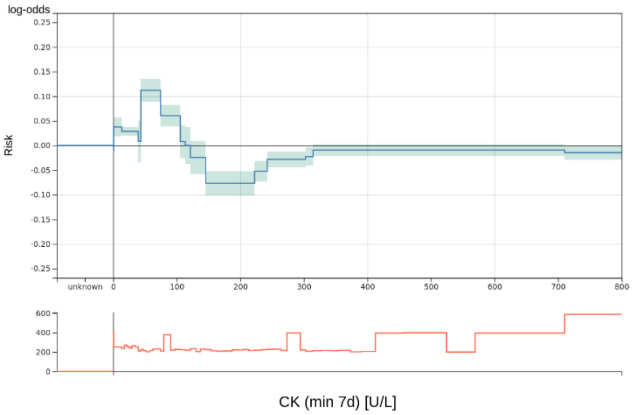


Relative importance: 1.15%

Applicable exclusion criteria: -

Notes:

- Severity of a surgery probably a confounder for this variable.

Decision: 1

**32.** RAS scale (trend per day 12h)

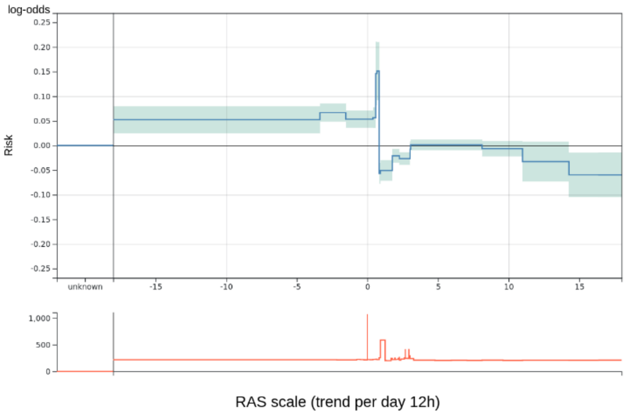


Relative importance: 1.13%

Applicable exclusion criteria: 2, 4

Notes:

- Trend of categorical variable not meaningful.

Decision: 3

**33.** Potassium (median 1d) [mmol/L]

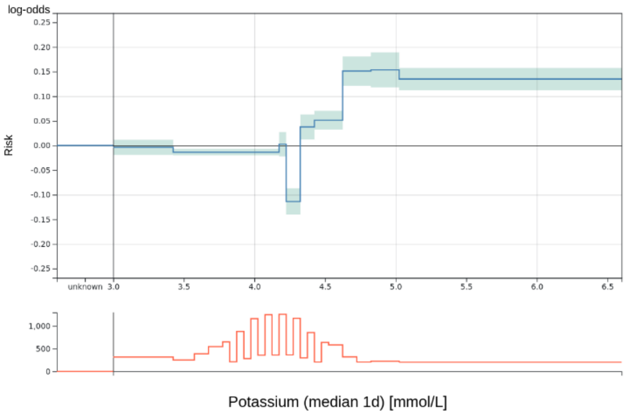


Relative importance: 1.13%

Applicable exclusion criteria: -

Notes:

- Drop at 4.25 probably due to cardiac surgery patients that have this as a target value.

Decision: 1

**34.** GCS score (min 3d)

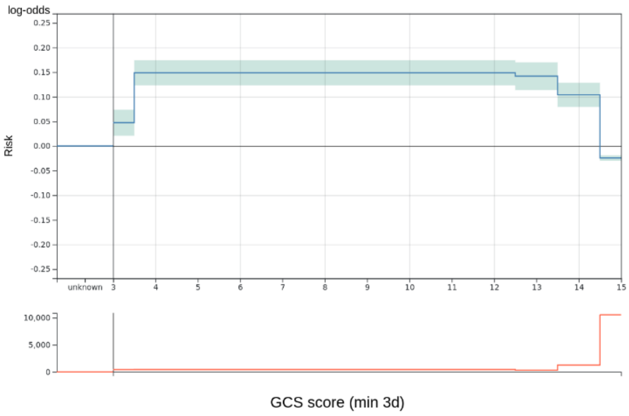


Relative importance: 1.11%

Applicable exclusion criteria: -

Notes:

- Lower risk for GCS 3 probably due to sedated patients after surgery.

Decision: 1

**35.** Body core temperature (median 1d) [°C]

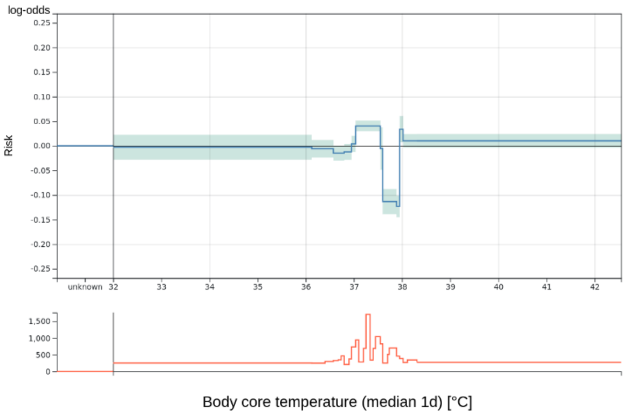


Relative importance: 1.10%

Applicable exclusion criteria: -

Notes: -

Decision: 1

**36.** BE (iqr 3d) [mmol/L]

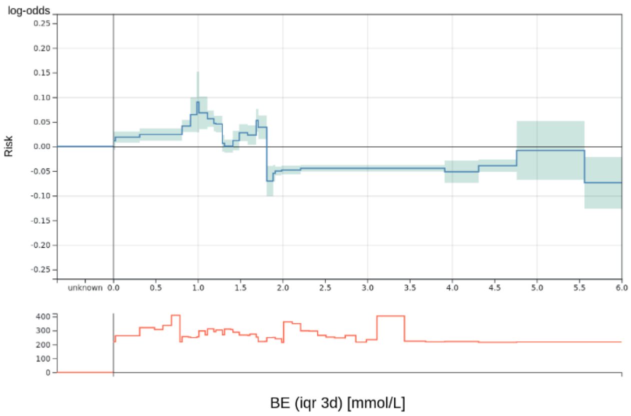


Relative importance: 1.10%

Applicable exclusion criteria: 3, 4

Notes:

- Low risk for large values against medical practice.

Decision: 3

**37.** Blood Urea Nitrogen (min 3d) [mg/dL]

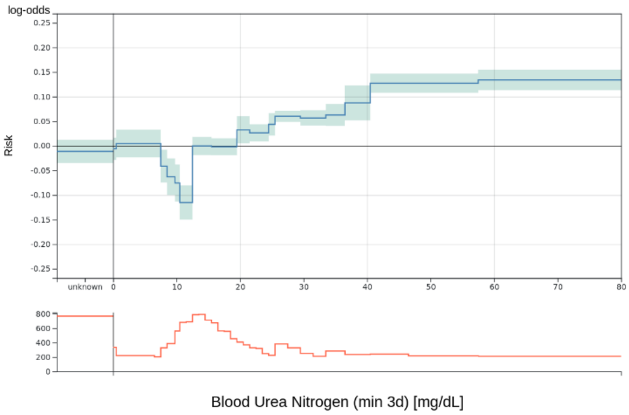


Relative importance: 1.10%

Applicable exclusion criteria: -

Notes: -

Decision: 1

**38.** paO2/FiO2 (trend per day 3d) [mmHg/FiO2]

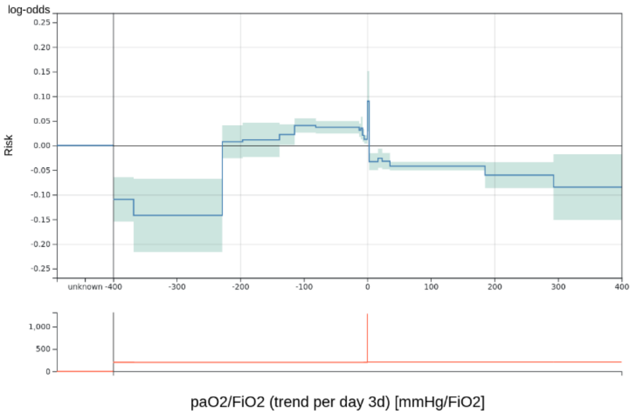


Relative importance: 1.09%

Applicable exclusion criteria: 4

Notes: -

Decision: 2

**39.** Drugs for constipation (unique 1d)

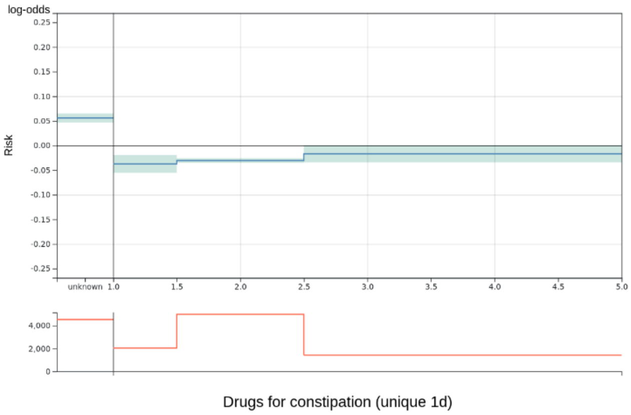


Relative importance: 1.09%

Applicable exclusion criteria: -

Notes: -

Decision: 1

**40.** Urine volume out (extrapolate 7d) [mL]

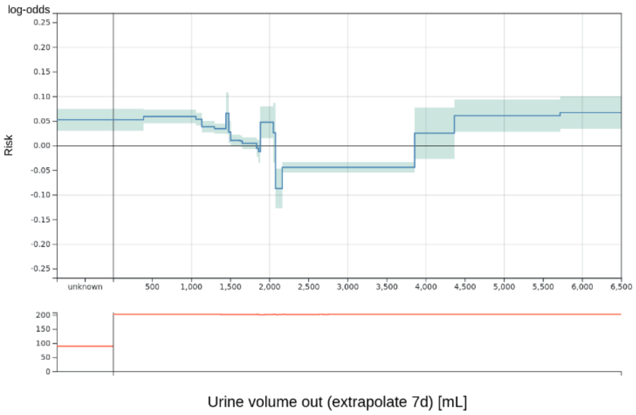


Relative importance: 1.09%

Applicable exclusion criteria: -

Notes: -

Decision: 1

**41.** PTT (min 7d) [s]

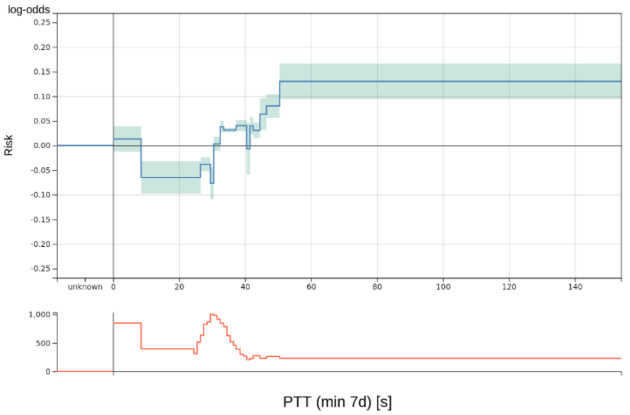


Relative importance: 1.07%

Applicable exclusion criteria: 2

Notes:

- Practice of measuring PTT changed in 2019, which cannot be corrected easily.

Decision: 3

**42.** Diastolic blood pressure (median 1d) [mmHg]

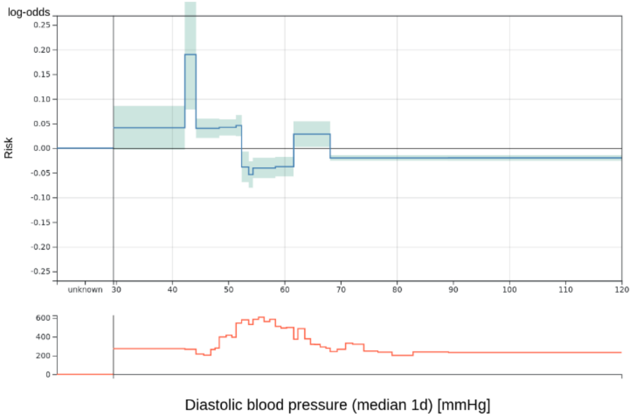


Relative importance: 1.06%

Applicable exclusion criteria: 3

Notes:

- Peak at 43 considered problematic.

Decision: 2

**43.** pO2 (min 12h) [mmHg]

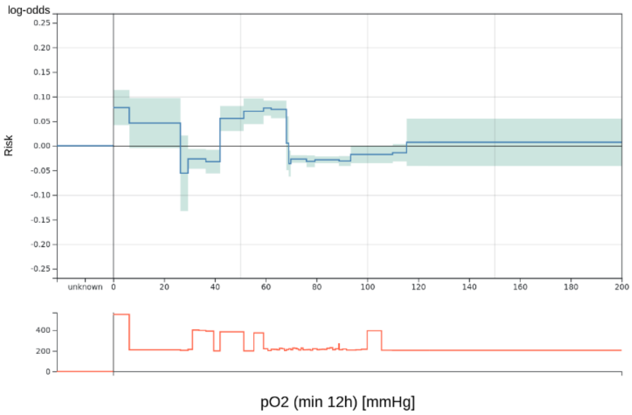


Relative importance: 1.06%

Applicable exclusion criteria: 2

Notes:

- Drop at 26.45 to 42.05 because of venous blood gas analyses.

Decision: 2

**44.** CK-MB (max 3d) [U/L]

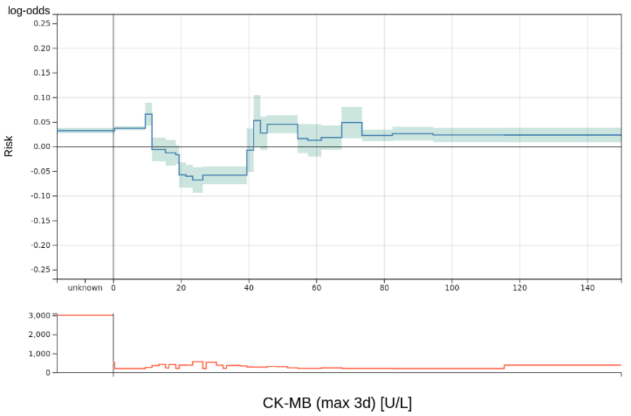


Relative importance: 1.05%

Applicable exclusion criteria: -

Notes: -

Decision: 1

**45.** RAS scale (max 1d)

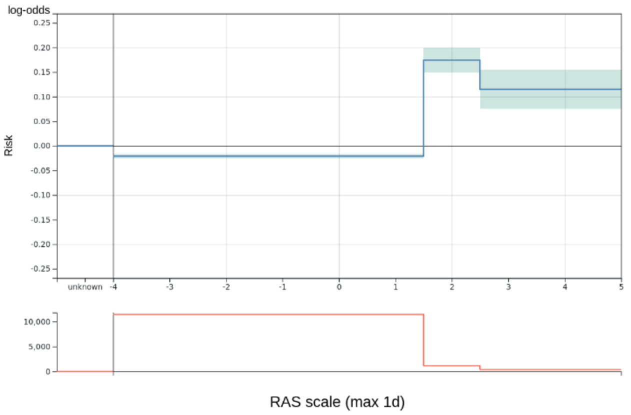


Relative importance: 1.05%

Applicable exclusion criteria: -

Notes: -

Decision: 1

**46.** PTT (min 3d) [s]

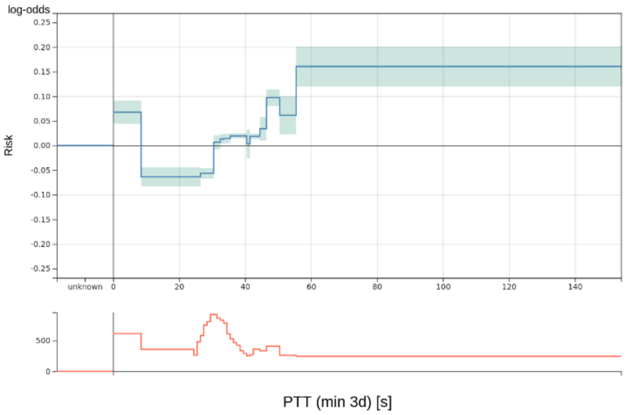


Relative importance: 1.05%

Applicable exclusion criteria: 2

Notes:

- Practice of measuring PTT changed in 2019, which cannot be corrected easily.

Decision: 3

**47.** Systolic blood pressure (iqr 12h) [mmHg]

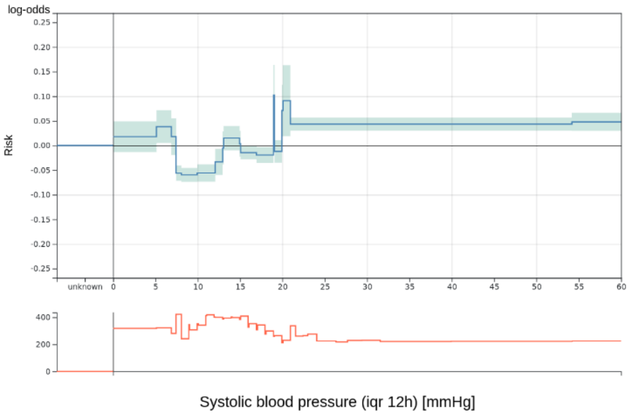


Relative importance: 1.05%

Applicable exclusion criteria: 4

Notes: -

Decision: 2

**48.** paO2/FiO2 (median 3d) [mmHg/FiO2]

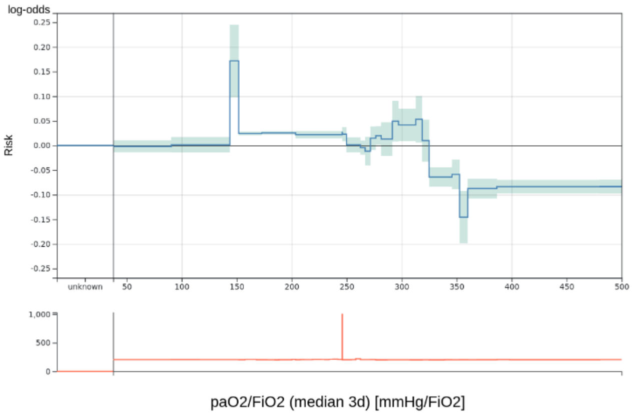


Relative importance: 1.04%

Applicable exclusion criteria: -

Notes: -

Decision: 1

**49.** CK (median 7d) [U/L]

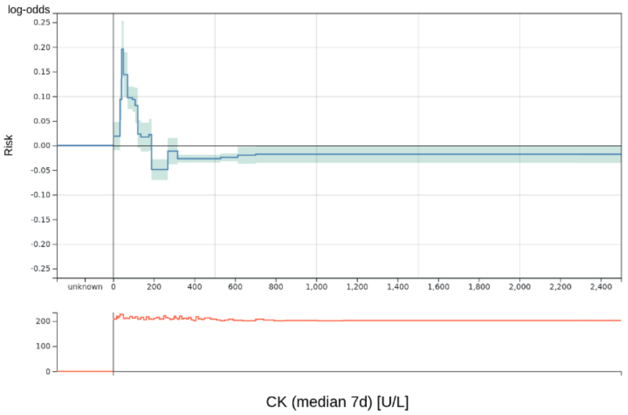


Relative importance: 1.04%

Applicable exclusion criteria: 3

Notes:

- Peak at 45 probably due to non-surgery patients (confounder). High values for example after heart surgery.
- Low risk for high values against medical knowledge.

Decision: 3

**50.** Lactate (max 3d) [mmol/L]

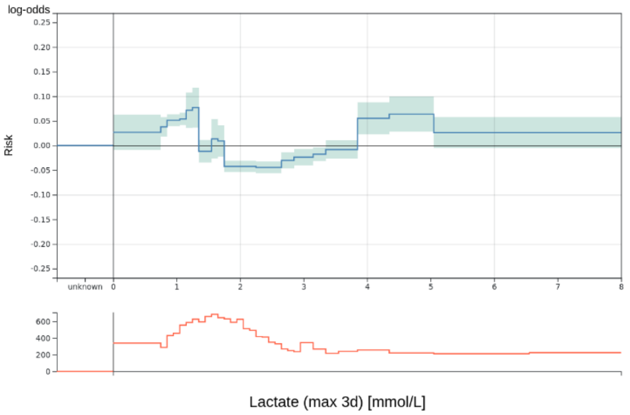


Relative importance: 1.04%

Applicable exclusion criteria: -

Notes:

- Drop only at 1.8 and not earlier probably because higher lactate values are monitored more closely.

Decision: 1

**51.** CK-MB (median 3d) [U/L]

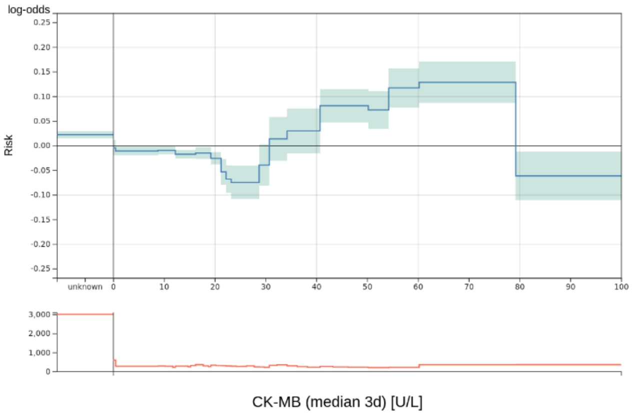


Relative importance: 1.04%

Applicable exclusion criteria: -

Notes: -

Decision: 1

**52.** Lactate (min 12h) [mmol/L]

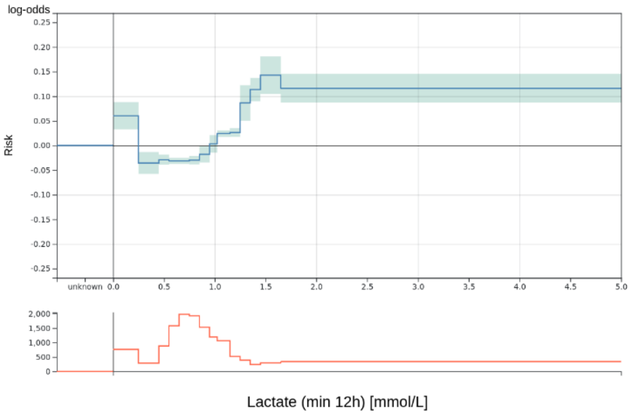


Relative importance: 1.00%

Applicable exclusion criteria: -

Notes: -

Decision: 1

**53.** Phosphate (max 1d) [mg/dL]

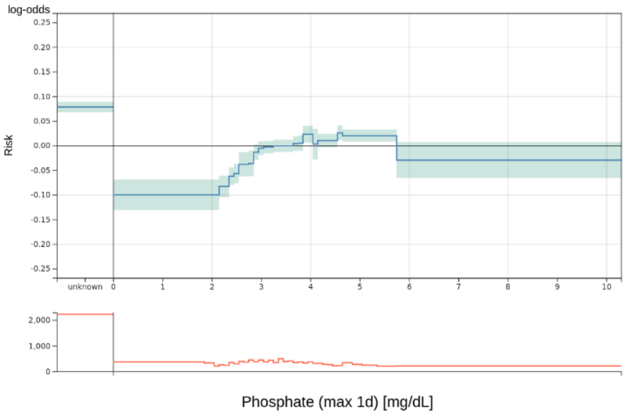


Relative importance: 1.00%

Applicable exclusion criteria: -

Notes: -

Decision: 1

**54.** PTT (max 7d) [s]

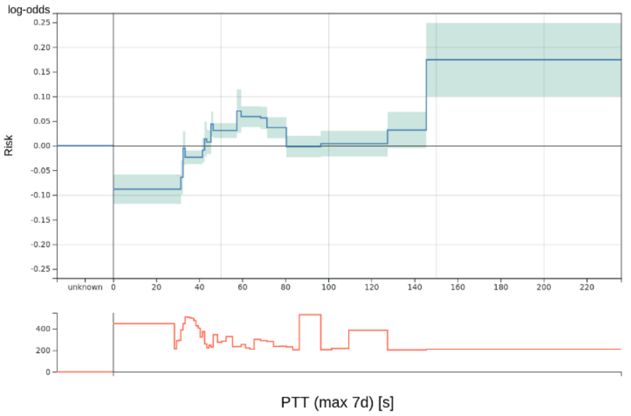


Relative importance: 0.98%

Applicable exclusion criteria: 2

Notes:

- Practice of measuring PTT changed in 2019, which cannot be corrected easily.

Decision: 3

**55.** pCO2 (median 1d) [mmHg]

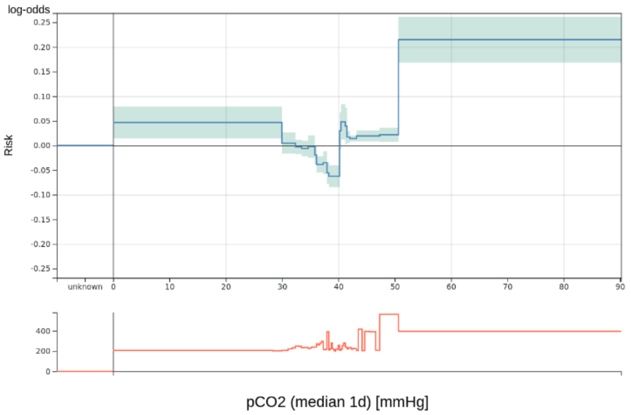


Relative importance: 0.98%

Applicable exclusion criteria: -

Notes: -

Decision: 1

**56.** BE (trend per day 3d) [mmol/L]

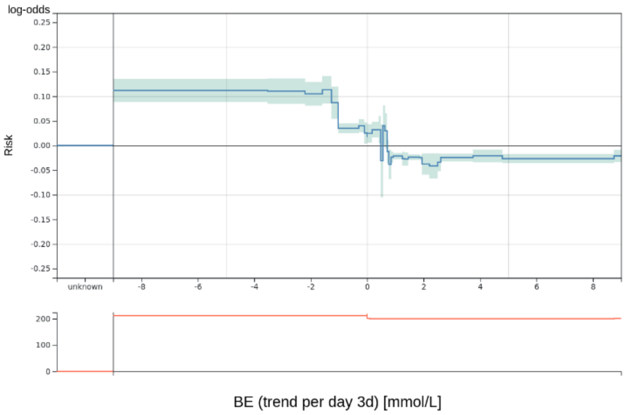


Relative importance: 0.97%

Applicable exclusion criteria: -

Notes: -

Decision: 1

**57.** Glucose (median 3d) [mg/dL]

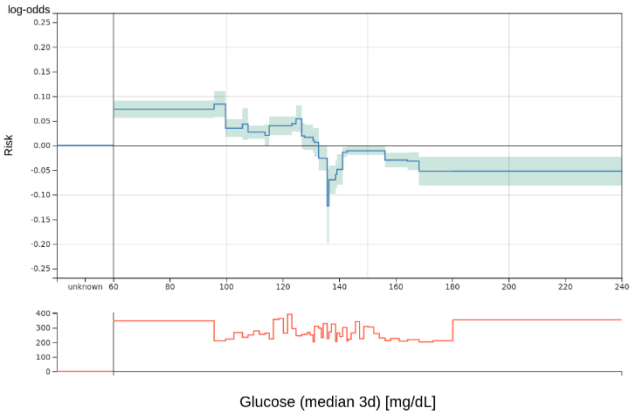


Relative importance: 0.97%

Applicable exclusion criteria: -

Notes: -

Decision: 1

**58.** BE (min 12h) [mmol/L]

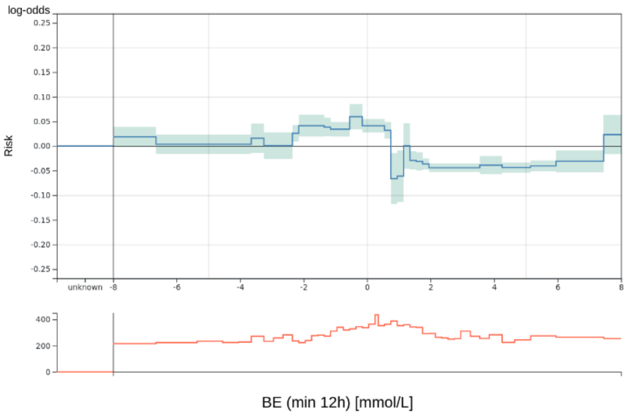


Relative importance: 0.96%

Applicable exclusion criteria: -

Notes: -

Decision: 1

**59.** MetHb (min 12h) [%]

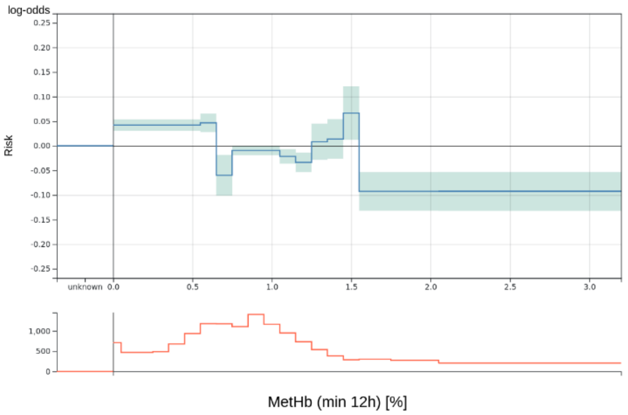


Relative importance: 0.96%

Applicable exclusion criteria: 4

Notes:

- Seldomly used in clinical practice.

Decision: 2

**60.** Is on automatic ventilation (days since last application per icu stay)

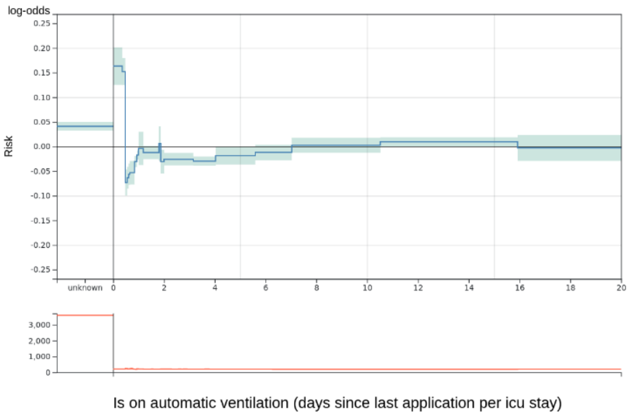


Relative importance: 0.95%

Applicable exclusion criteria: -

Notes:

- Drop at 0.5 similarly to tubus features probably because of regularly treated surgical patients.

Decision: 1

**61.** Body core temperature (min 4h) [°C]

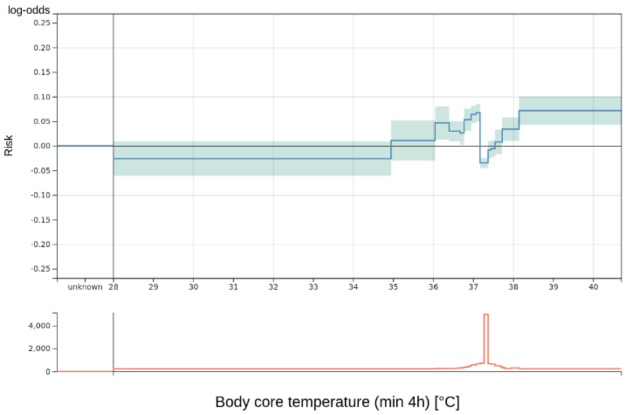


Relative importance: 0.95%

Applicable exclusion criteria: 3, 4

Notes:

- Values in normal range receive very different risk values.

Decision: 3

**62.** pCO2 (iqr 1d) [mmHg]

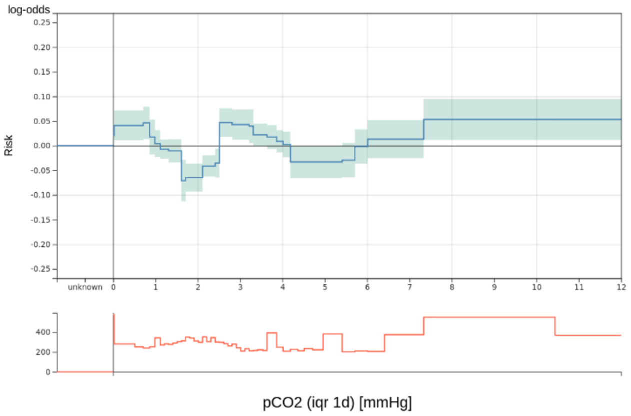


Relative importance: 0.95%

Applicable exclusion criteria: 4

Notes: -

Decision: 2

**63.** Sodium (median 3d) [mmol/L]

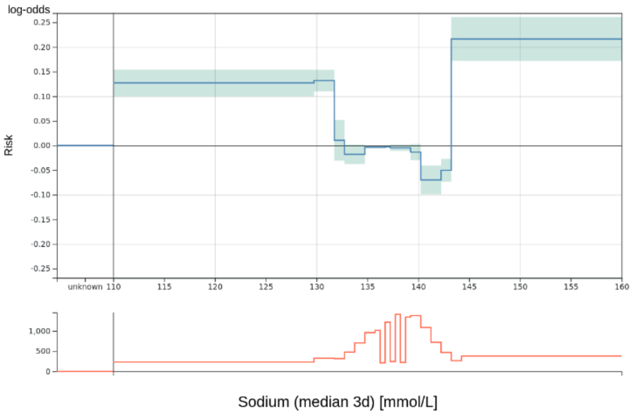


Relative importance: 0.93%

Applicable exclusion criteria: -

Notes: -

Decision: 1

**64.** Leucocytes (median 1d) [thousand/µL]

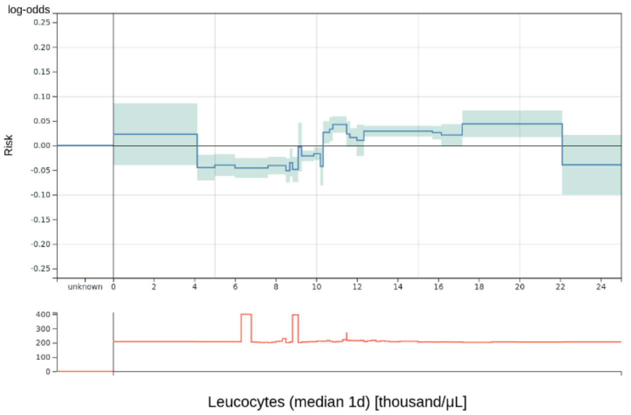


Relative importance: 0.92%

Applicable exclusion criteria: -

Notes: -

Decision: 1

**65.** Sodium (trend per day 3d) [mmol/L]

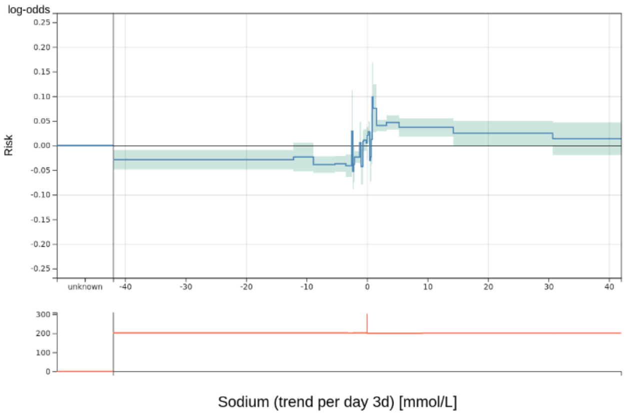


Relative importance: 0.92%

Applicable exclusion criteria: 4

Notes: -

Decision: 2

**66.** Procalcitonin (max 7d) [ng/mL]

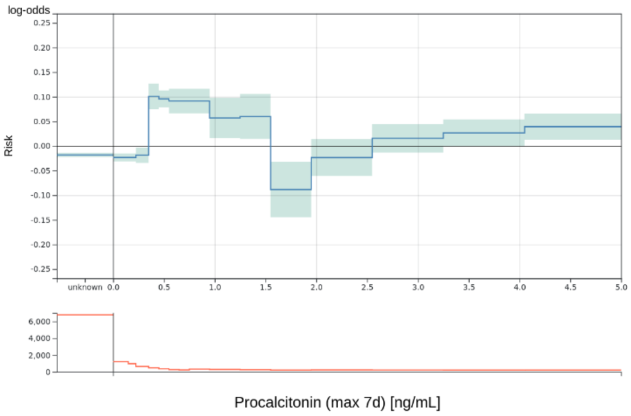


Relative importance: 0.91%

Applicable exclusion criteria: -

Notes:

- Lower risk for higher values probably because monitored more closely.

Decision: 1

**67.** BE (median 12h) [mmol/L]

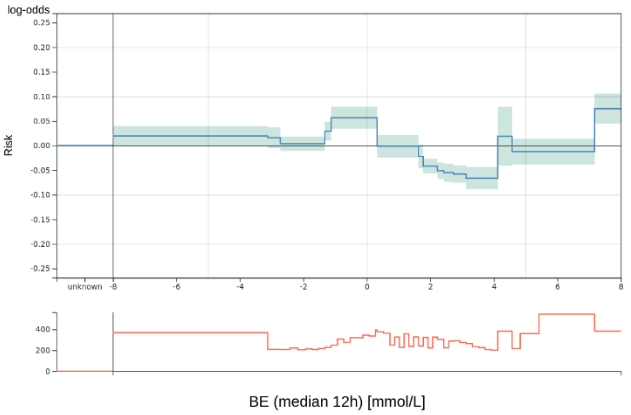


Relative importance: 0.91%

Applicable exclusion criteria: -

Notes: -

Decision: 1

**68.** Mean blood pressure (median 4h) [mmHg]

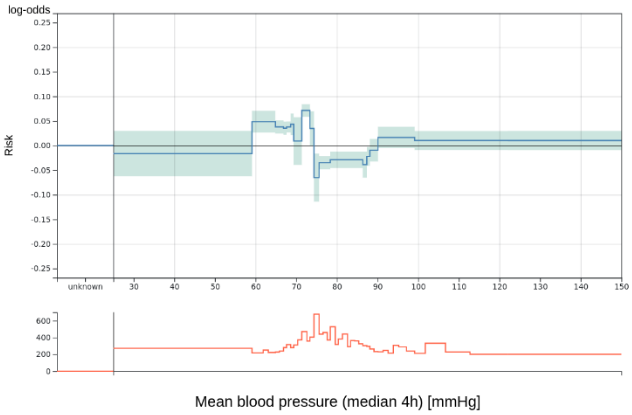


Relative importance: 0.87%

Applicable exclusion criteria: -

Notes: -

Decision: 1

**69.** Leucocytes (trend per day 3d) [thousand/μL]

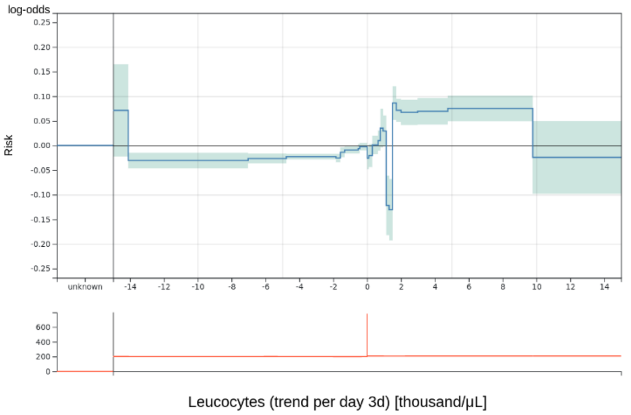


Relative importance: 0.84%

Applicable exclusion criteria: 3

Notes:

- No medical explanation for the drop at 1.2.

Decision: 3

**70.** pH (median 3d)

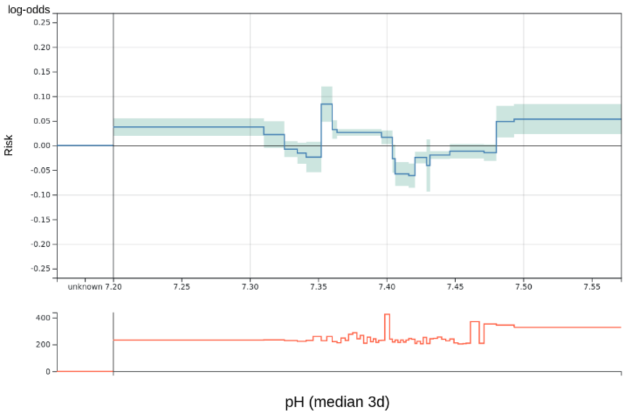


Relative importance: 0.84%

Applicable exclusion criteria: -

Notes: -

Decision: 1

**71.** Bilirubin total (max 7d) [mg/dL]

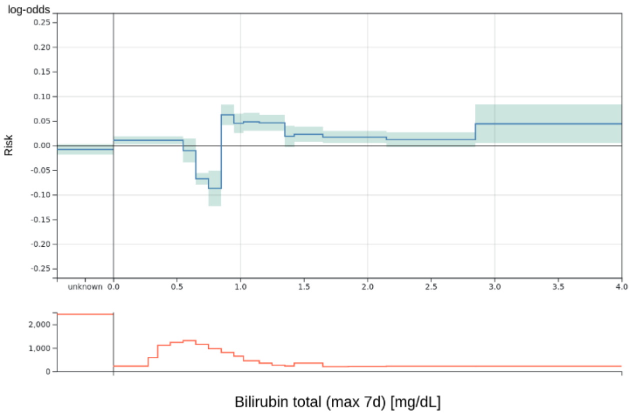


Relative importance: 0.84%

Applicable exclusion criteria: -

Notes: -

Decision: 1

**72.** pO2 (iqr 12h) [mmHg]

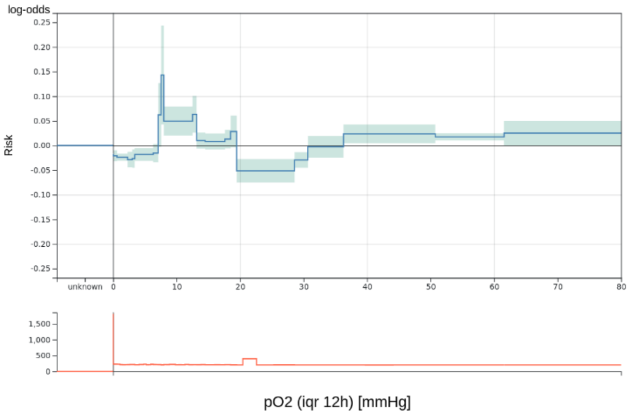


Relative importance: 0.84%

Applicable exclusion criteria: 3

Notes: -

Decision: 2

**73.** BE (iqr 1d) [mmol/L]

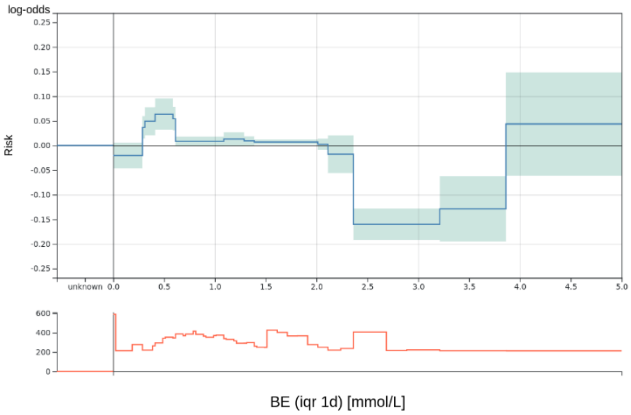


Relative importance: 0.83%

Applicable exclusion criteria: -

Notes: -

Decision: 1

**74.** Body core temperature (trend per day 1d) [°C]

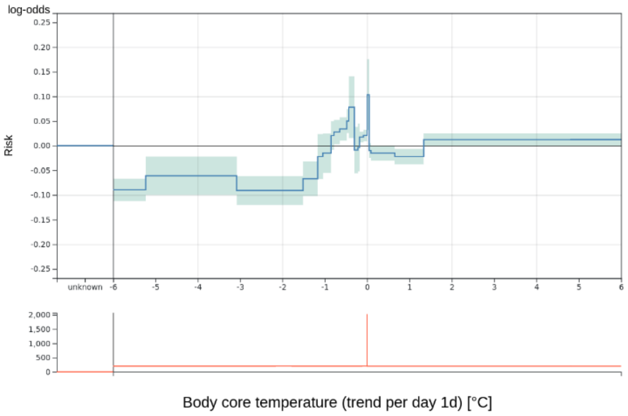


Relative importance: 0.83%

Applicable exclusion criteria: 3, 4

Notes:

- Even though difficult to interpret, no clear evidence to remove it.

Decision: 2

**75.** C-reactive protein (max 3d) [mg/dL]

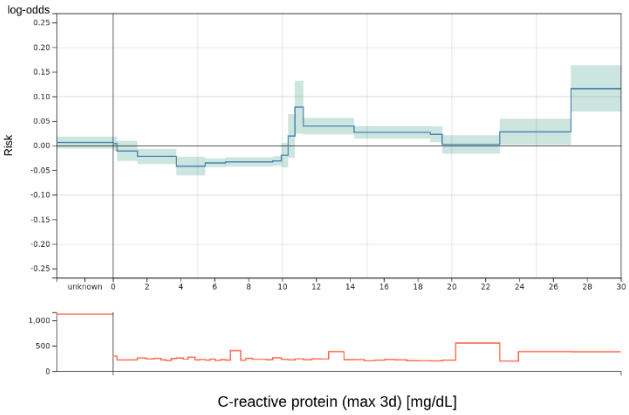


Relative importance: 0.83%

Applicable exclusion criteria: -

Notes: -

Decision: 1

**76.** Heart rate (min 1d) [bpm]

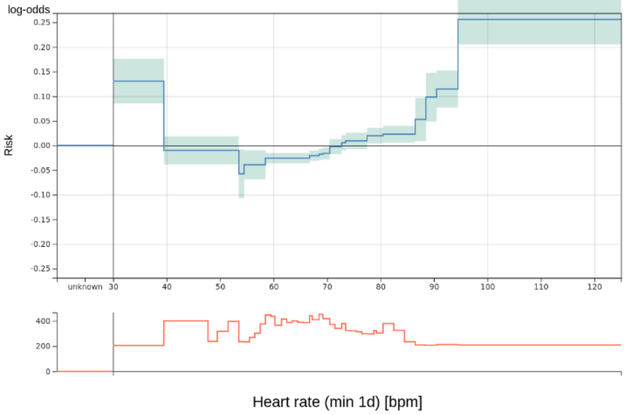


Relative importance: 0.82%

Applicable exclusion criteria: -

Notes: -

Decision: 1

**77.** Hematocrit (median 12h) [%]

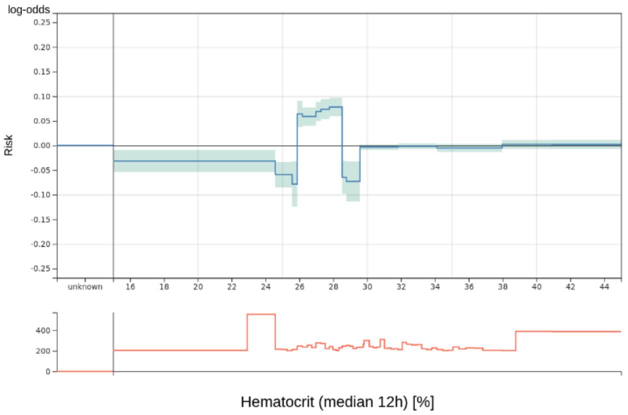


Relative importance: 0.80%

Applicable exclusion criteria: 3

Notes:

- Drops at 25 and 28 against medical knowledge but effect considered small.

Decision: 2

**78.** pCO2 (min 3d) [mmHg]

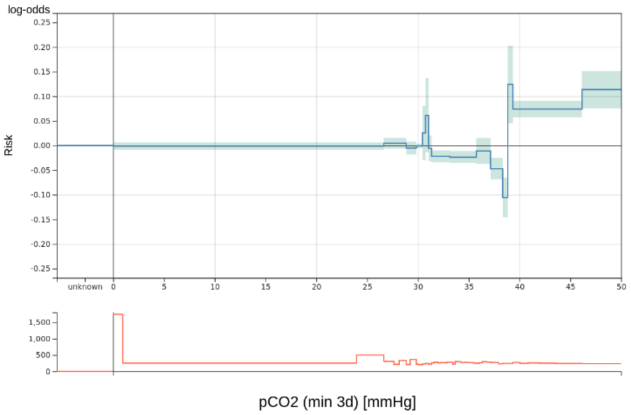


Relative importance: 0.76%

Applicable exclusion criteria: -

Notes: -

Decision: 1

**79.** Mean blood pressure (median 12h) [mmHg]

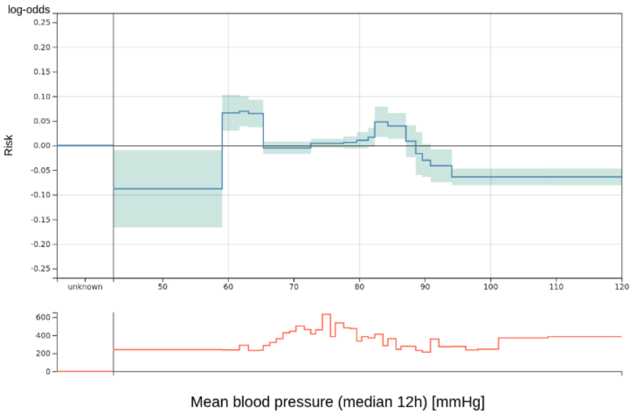


Relative importance: 0.72%

Applicable exclusion criteria: -

Notes: -

Decision: 1

**80.** Calcium (max 1d) [mmol/L]

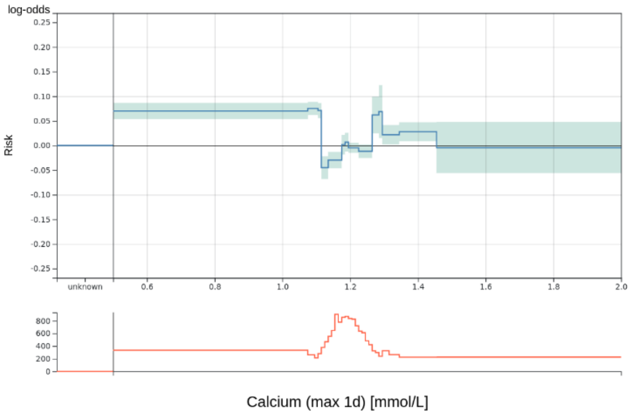


Relative importance: 0.69%

Applicable exclusion criteria: -

Notes: -

Decision: 1

**81.** Estimated respiratory rate (median 1d)

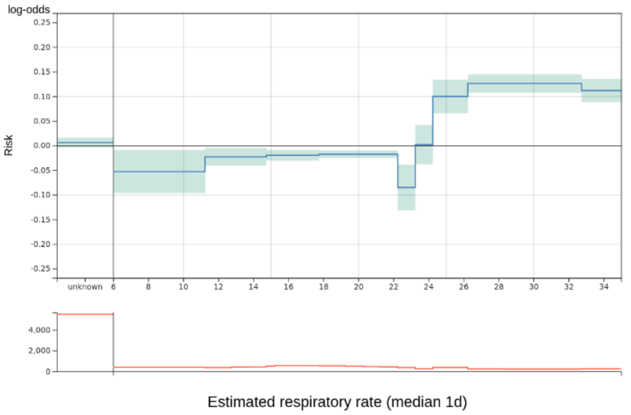


Relative importance: 0.68%

Applicable exclusion criteria: -

Notes: -

Decision: 1

**82.** pH (iqr 1d)

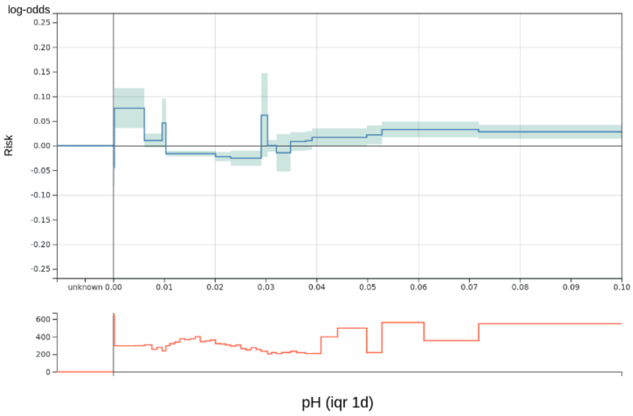


Relative importance: 0.67%

Applicable exclusion criteria: -

Notes: -

Decision: 1

**83.** Leucocytes (iqr 3d) [thousand/μL]

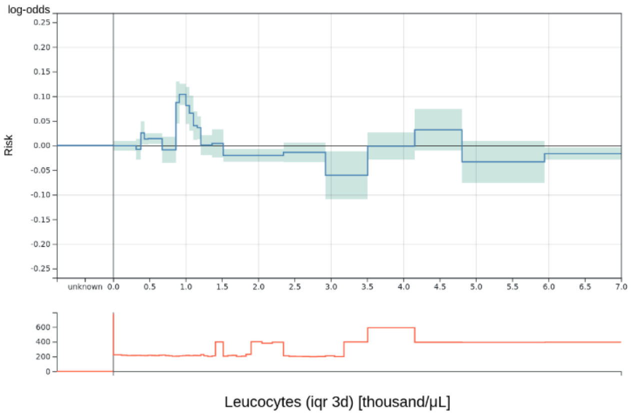


Relative importance: 0.63%

Applicable exclusion criteria: 4

Notes: -

Decision: 2

**84.** Heart rate (iqr 4h) [bpm]

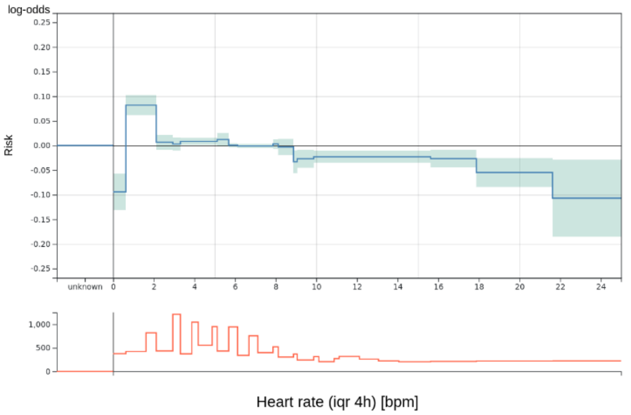


Relative importance: 0.60%

Applicable exclusion criteria: -

Notes: -

Decision: 1

**85.** RHb (median 12h)

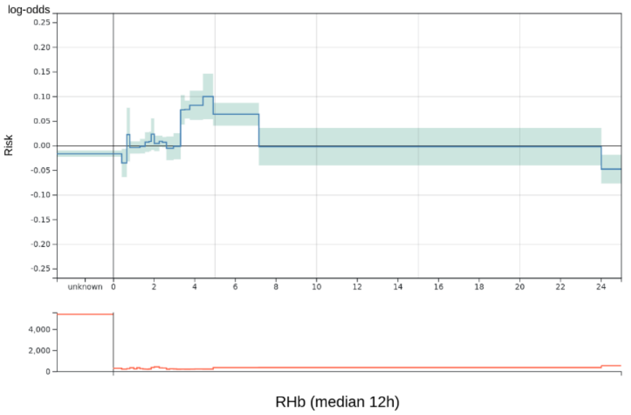


Relative importance: 0.60%

Applicable exclusion criteria: 4

Notes:

- Calculated variable without medical relevance that is only used as quality measure in clinical practice, so cannot determine its effect.

Decision: 3

### Comments during EBM model inspection

General comments

1. Most risk functions exhibited one of the listed problems to some extent. However, often the effect should not be large. Hence, we agreed to only exclude risk functions when a problem was clearly fulfilled and there is a considerable impact for patients.
2. Small peaks and drops of the risk function were often ignored. Especially, when they occurred in a sparse region where probably only a few patients would be affected (e.g. 7, 23).
3. Outlier values often received inexplicable risk values, however, since only a few patients would be affected or since they are caused by erroneous data they were often ignored (e.g. 42).
4. It is unusual in clinical practice to consider only isolated variables or pairs, so this information was sometimes not sufficient for a reasonable decision (e.g. 66).
5. It was difficult to interpret a feature over the complete cohort because sub-cohorts behaved or were treated very differently (e.g. 13). For instance, features over long time intervals contain patients with very short and long stays.
6. Feature values did depend on the measuring frequency which could differ between patients and was unknown during risk function inspection (e.g. 62).
7. The severity of surgery probably acted as a confounder for some variables. However, adding surgery as an explicit variable did not change the behavior, so there was probably a more complicated relationship (e.g. in 31 increase of CK as a sign of the severity of the surgery).
8. It was difficult to consider the situation at discharge because many variables were also used routinely during the ICU stay. Also, different clinical paths could lead to the same situation at discharge and it was hard to incorporate them all.
9. The team tended to construct an explanation for each risk function even though it is very hard to find evidence for different explanations (e.g. 8, 17).
10. It was hard to interpret risk for values outside the usual value ranges and variables that were seldomly used in clinical practice (e.g. 8, 15, 59).
11. There was limited intuition for IQR and trend features and it was very hard to mentally switch between the different quantities during inspection (e.g. 36, 62). Also, small values for trend features were hard to interpret (e.g. 20).
12. Even though the content of the 2D risk function could be understood it was very hard to determine its clinical relevance. Often variables were combined that in practice are not interpreted together and have no obvious connection (1-5).
13. When a risk function was less interpretable there was a tendency to rely more on the model to find useful relationships and the risk function was included.

Factors hindering interpretability

14. Fluctuating risk functions that rapidly switch between positive and negative values made it hard to interpret them and to determine their effect (e.g. 20, 30, 47, 56).
15. It was difficult to interpret the number of cases in a bin because bin sizes differed (e.g. in 6, 12 histograms suggested even data distribution).
16. Bins were sometimes too large and included abnormal and normal values, so the team was unable to distinguish them (e.g. 30).
17. Minimum and maximum features were more susceptible to outlier values (e.g. 30).

Factors supporting interpretability

18. It was helpful to see the data distribution as a sanity check to verify the item distribution and see if it was collected as expected.

### External validation

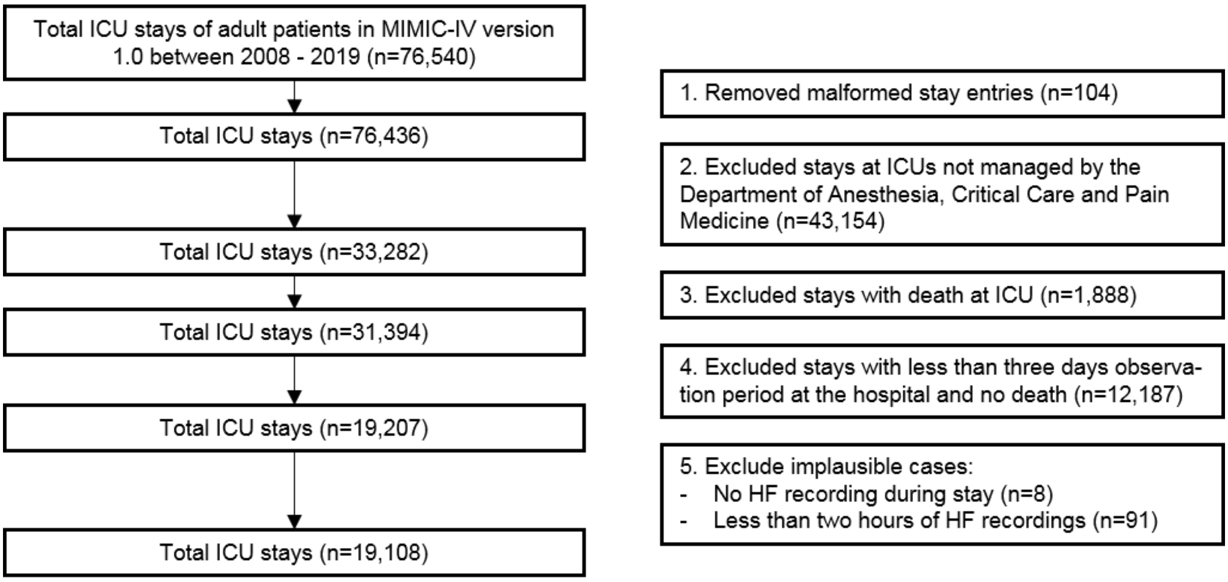


*Figure 4. **Cohort selection for the MIMIC cohort**. We tried to apply the same selection procedure as for the UKM cohort. Merging was not necessary since MIMIC-IV already contained consecutive ICU stays.*

*Table 9. **Overview of the MIMIC cohort**.*

| **Characteristic** | **All ICU stays** | **No 3-day readmission or death after ICU discharge** | **3-day readmissions or death after ICU discharge** |
|---|---|---|---|
| Number of stays | 19,108 (100.0%) | 17,482 (91.5%) | 1,626 (8.5%) |
| Number of patients | 17,499 (100.0%) | 16,591 (94.8%) | 1,525 (8.7%) |
| Mean age (SD) | 65.16 (15.86) | 64.81 (15.86) | 68.97 (15.34) |
| Gender female | 7,681 (100.0%) | 6,971 (90.8%) | 710 (9.2%) |
| Gender male | 11,427 (100.0%) | 10511 (92.0%) | 916 (8.0%) |
| Mean length of ICU stay (SD) | 3.52 days (4.69 days) | 3.47 days (4.63 days) | 4.05 days (5.26 days) |
| ICU at discharge | Cardiac Vascular Intensive Care Unit (n=6,891)<br>Surgical Intensive Care Unit (n=6,397)<br>Trauma SICU (n=5,291)<br>Neuro Surgical Intensive Care Unit (n=529) | Cardiac Vascular Intensive Care Unit (n=6,523)<br>Surgical Intensive Care Unit (n=5,778)<br>Trauma SICU (n=4,744)<br>Neuro Surgical Intensive Care Unit (n=437) | Cardiac Vascular Intensive Care Unit (n=368)<br>Surgical Intensive Care Unit (n=619)<br>Trauma SICU (n=547)<br>Neuro Surgical Intensive Care Unit (n=92) |

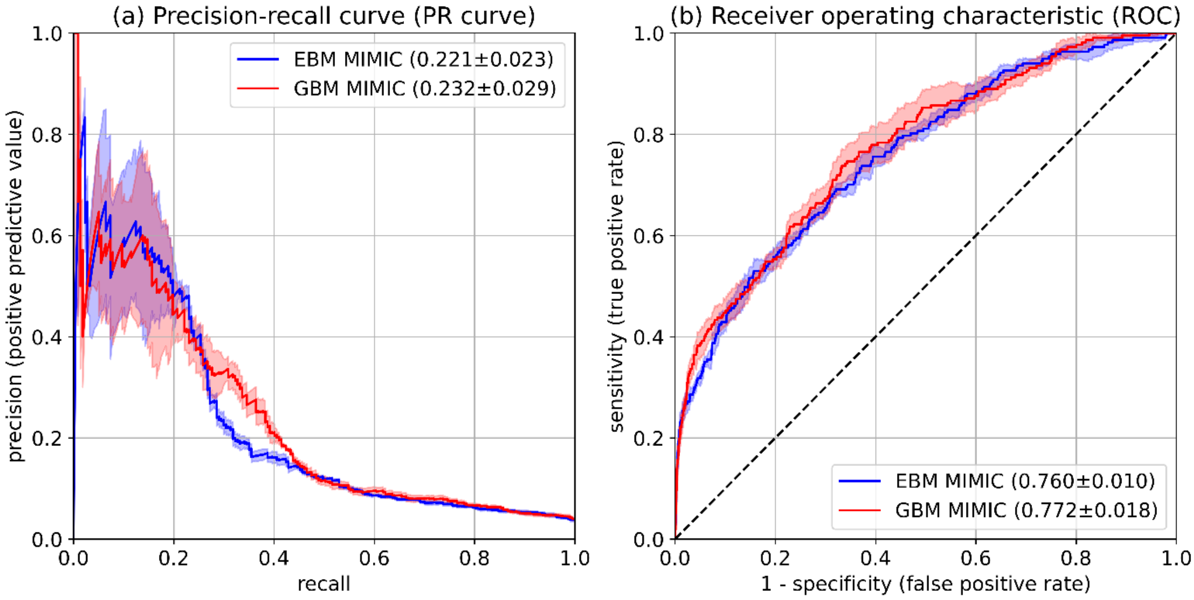


*Figure 5. **External validation of EBM and GBM model on the MIMIC cohort**. (a), (b) The same performance indicators were determined. Both models showed again similar results. Surprisingly, the performance of the UKM cohort was much better. We attribute this to the better data quality of the MIMIC-IV database and differences in the collection process.*
